# Machine Learning-Based Prediction of Cell-type Resolved Brain eQTLs Enhances Discovery of Variants Explaining Alzheimer’s Disease Heritability

**DOI:** 10.64898/2025.12.03.25341562

**Authors:** Chirag M Lakhani, Giacomo Cavalca, Anjing Liu, Rohan Nidumbur, Ru Feng, Towfique Raj, Philip De Jager, The Alzheimer’s Disease Functional Genomics Consortium, Gao Wang, David A. Knowles

**Affiliations:** New York Genome Center, New York, NY, USA; University of Bologna, Bologna, Italy; Center for Statistical Genetics, The Gertrude H. Sergievsky Center, Columbia University, New York, NY, USA; University of Maryland, College Park, College Park, MD, USA; Neuroscience, Icahn School of Medicine, Mount Sinai, New York, NY, USA; Department of Neurology, Columbia University, New York, NY, USA; Department of Biostatistics, Columbia University, New York, NY, USA; Department of Computer Science, Columbia University, New York, NY, USA; Department of Systems Biology, Columbia University, New York, NY, USA; Data Science Institute, Columbia University, New York, NY, USA

## Abstract

The majority of causal genome-wide association studies (GWAS) variants for Alzheimer’s disease (AD) are believed to reside in noncoding regions of the genome, where they likely affect gene regulation, particularly in microglia. Although expression Quantitative Trait Loci (eQTL) studies offer valuable insights into gene regulation, they tend to identify variants in the promoter regions of genes under weaker selection. In contrast, GWAS variants are often found in enhancer regions linked to genes under stronger selection. To address this discrepancy, we developed predictive models, called single-cell Enhanced Expression Modifier Scores (scEEMS), to identify cell type-specific eQTLs using 4,839 genomic features, including deep learning-based scores that predict the effects of variants on various molecular phenotypes.

These models were trained on fine-mapped single-cell eQTLs from six cell types and exhibited strong performance, with an average cross-validation area under the precision-recall curve (AUPRC) of 0.67. Notably, for microglia, the predicted eQTLs explained 15.3% of AD GWAS heritability, with a 140.6-fold enrichment of heritability (p-value 5.15 x 10^⁻6^), compared to just 5% of heritability explained by fine-mapped eQTLs. Incorporating scEEMS predictions as priors in eQTL fine-mapping refined credible sets and resulted in a net gain of 107 eGenes in microglia and 271 eGenes in astrocytes, reflecting improved statistical power that both identified new associations and filtered false positives. We used these models to link variants to cell type-specific genes and then applied the eMAGMA framework to nominate cell type-specific AD risk genes based on GWAS data. Our eMAGMA approach using predicted eQTLs identified 215 cell type-gene pairs (111 unique genes), substantially more than the 76 pairs (55 unique genes) found using fine-mapped eQTLs. Among these, we identified 43 microglia-specific, 18 astrocyte-specific, and 15 oligodendrocyte-specific AD risk genes. Of the 215 pairs, 62 replicated in at least one non-European population (African-American, Hispanic, or East Asian), compared to only 29 from fine-mapped eQTLs. Importantly, 18 of the replicated pairs represent novel discoveries not found by standard MAGMA or fine-mapped eMAGMA analyses. Five cell type-gene pairs replicated across European and at least two non-European populations: *BIN1* (microglia), *PICALM* (microglia), *ABCA7* (astrocyte and oligodendrocyte), and *ARHGAP45* (microglia).

## Introduction

A central post-GWAS challenge is elucidating the functional consequences of disease-associated noncoding variants. It is believed that many of these variants modulate gene regulation (e.g., transcription, splicing or stability) of genes, which then has downstream effects on disease risk. Large consortia such as GTEx^1^ have generated population-scale functional genomics data, uncovering expression quantitative trait loci (eQTL) that modulate baseline gene expression in bulk tissue. However, for a typical trait only 11% of disease heritability is mediated through such bulk eQTLs^2^. This discrepancy has been referred to as the ‘missing regulation^3^’ problem, where disease-associated variants are strongly enriched in regulatory regions of the genome but are often not explained by any bulk tissue eQTLs. Recent theoretical and empirical analysis has shown that there are distributional differences in the types of variants and genes implicated in eQTL studies compared to disease GWAS^4^. In particular, eQTLs are more likely to be found in promoter regions, whereas disease-associated GWAS variants are more likely to be found in distal enhancer regions. Genes with eQTLs tend to be under less selective constraint compared to disease-associated GWAS genes. Bulk-tissue eQTLs may not capture enough of the cell or context specificity needed to find a disease-associated eQTL, especially for rarer cell types^5^.

Both single-cell eQTL^6,7^ and context-specific eQTL studies^8^ have uncovered additional disease-associated eQTLs beyond bulk studies. However, a fundamental challenge remains: the substantial disparity in sample sizes between eQTL and GWAS studies limits colocalization analyses. For example, our single-cell eQTL dataset contains 788 samples^9,10^, whereas a recent Alzheimer’s disease GWAS^11^ has a sample size of 788,989 individuals—a thousand-fold difference. This disparity makes it particularly difficult to directly validate low MAF variant eQTLs that may colocalize with GWAS risk variants, as many variants reach genome-wide significance in well-powered GWAS but lack statistical power for detection in eQTL cohorts, especially for rare cell types like microglia.

Another approach to elucidating the functional consequences of disease associated variants is to leverage intermediate molecular QTLs such as chromatin accessibility QTLs (caQTLs).

Recent studies have shown that caQTLs mediate a larger percentage of disease heritability compared to eQTLs^12,13^, showing the promise of other molecular QTLs for elucidating the function of disease variants. However, it is challenging to generate population-scale data for all possible molecular phenotypes in all possible cellular contexts. Genomic deep learning models^14–18^, which predict epigenomic profiles as a function of genomic sequence, can help overcome this limitation by providing cell and assay-specific variant prediction scores for any genetic variant. These models are typically trained either on a single genomic assay or in a multi-task fashion using thousands of functional genomics assays from the Roadmap^19^ or ENCODE^20^ consortia.

Building upon the Expression Modifier Score^21^ (EMS), we propose single-cell Enhanced Expression Modifier Scores (scEEMS) for the prediction of causal single-cell eQTLs as a function of distance, ABC scores^22,23^, cell type and non cell type-specific features, deep learning based variant effect prediction (DL-VEP) scores, and gene features^24^. We developed a framework for training such models in the context of challenges arising in single-cell eQTL mapping, such as reduced power for rare cell types. We use the gradient boosting model CatBoost^25^ to predict causal eQTL status as a function of 4,839 features for 6 brain cell types profiled by single-nucleus post-mortem brain RNA-seq from the ROSMAP^9,10,26^ cohort. Single cell eQTL summary statistics and subsequent fine-mapping results were generated as part of the FunGen-xQTL resource^27,28^ within the Alzheimer’s Disease Sequencing Project^29^. We use scEEMS to nominate putative causal cell type eQTLs, especially in enhancer regions, which are likely to be more functionally relevant. We used SHapley Additive exPlanations (SHAP)^30,31^ model interpretation to calculate Shapley values for all high probability predicted eQTLs (**Online Methods**). On average, the largest contributing set of features were the Enformer^15^ DL-VEP scores. We also compared the contribution of fine-mapped versus predicted eQTLs to a recent large-scale AD GWAS meta-analysis^11^, referred to as Bellenguez AD GWAS hereafter. While fine-mapped eQTLs had higher enrichment of heritability in the GWAS, the predicted eQTLs explained more trait heritability while still maintaining high enrichment of heritability. We also integrated scEEMS predictions as informative priors during fine-mapping, which improved variant prioritization and resulted in a net gain of 107 eGenes in microglia and 271 in astrocytes by enhancing detection of true associations while filtering false positives. Finally, we utilized scEEMS to link variants to cell type-specific genes and then used the eMAGMA^32^ framework to nominate cell type-specific AD risk genes based on cross-ancestry AD GWAS data. The scEEMS approach nominated 215 cell type-gene pairs compared to only 75 cell type-gene pairs using fine-mapped eQTLs. Among the 215 cell type-gene pairs, there were 62 which replicated in a non-European population^33^. Finally, among the 62 replicated cell type-gene pairs there were 18 cell type-gene pairs which were uniquely scEEMS eMAGMA genes, showing the ability of scEEMS to nominate putative AD risk genes not found through fine-mapped eQTL or standard MAGMA approaches.

## Results

### Overview of scEEMS Training

scEEMS is a cell type-specific CatBoost binary classifier trained to predict whether a variant is a causal cell type-specific eQTL as a function of 4,839 variant, gene and variant-gene pair features. The fine-mapped eQTLs for this analysis were provided by the FunGen-xQTL data resource which harmonized three single-cell brain atlases (CUIMC1^10^, MIT^9^, and CUIMC2^34^) from 788 individuals in the ROSMAP cohort (**Supplementary Table 1 and Online Methods**).

Based on previous analysis^35^, the cells were mapped to 6 brain cell types: astrocytes (Ast), excitatory neurons (Exc), inhibitory neurons (Inh), microglia (Mic), oligodendrocytes (Oli), and oligodendrocyte precursor cells (OPC). Following data processing and normalization, genes were classified based on whether they met the mean log₂CPM ≥ 2.0 (counts per million) filtering criterion after combining all datasets (**Online Methods**). Genes meeting this threshold, designated as *MEGA* genes, were prioritized for a mega-analysis using the combined cohort to maximize statistical power for eQTL discovery and fine-mapping. Genes not meeting this threshold were analyzed in their original constituent cohorts where they had sufficient expression, designated as *Other* genes. For both gene categories, we performed fine-mapping for all cis-variants (+/- 1 MB of gene TSS) directly on the genotype and expression data. For *MEGA* genes, we applied SuSiE^36^ to the merged genotype matrices and pseudobulk expression counts from the combined cohort. For *Other* genes, we applied SuSiE to the subset genotype matrix and expression data from their constituent cohort. Prior to fine-mapping, expression data were residualized for batch effects, age, sex, and postmortem interval. This approach determined causal eQTL credible sets (CS) for each gene across all 6 cell types (**Online Methods and Figure 1a**).

**Figure 1:**
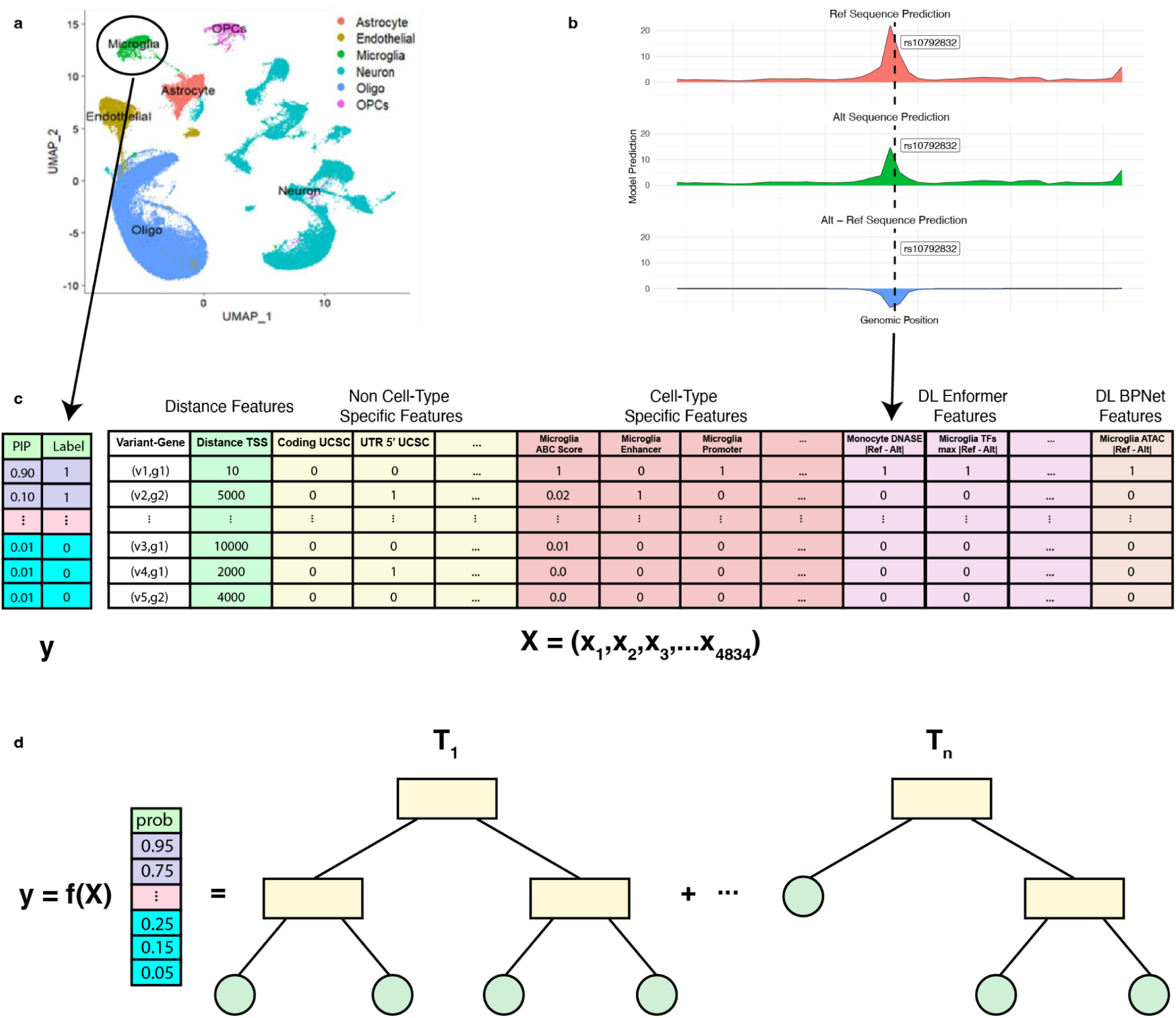
scEEMS framework for predicting single-cell eQTLs a) UMAP plot representing neuronal cell types which have single-cell eQTL data. b) Enformer predictions of *PU.1* TF binding for reference and alternate sequences as well as the difference in predictions. c) Feature matrix showing 4,839 features per variant-gene pair, including distance to TSS (log-transformed), ABC scores, non cell type-specific features, cell type-specific features, DL-VEP scores, and a gene feature. d) CatBoost model architecture for predicting eQTL probabilities as a function of the 4,839 features.

In contrast to the EMS training procedure where positive labels are variants with high (> 0.9) posterior inclusion probability (PIP), we designed scEEMS to leverage variants with weaker signals in the model training. This is motivated by three factors: 1) rarer cell types which may be important for the trait of interest (e.g. microglia for AD^37^) will have weaker eQTL signal, 2) at comparable sample size sc-eQTLs are typically less powered than bulk due to higher technical noise and sparsity in single-cell measurements^38^, resulting in fewer variants to train on, and 3) variants in enhancers will typically have weaker eQTL signal compared to variants in promoters yet they may be more disease relevant^4^. We reasoned that for rarer cell types, the increase of variants for model training could boost model performance. For our model training, we balance the need to be inclusive of weaker eQTLs while not drowning out the true signals. For each cell type, the positive class includes two types of variants: 1) all PIP > 0.05 variants that are in a CS where there is a variant with maximum PIP > 0.1 (variant) and 2) any PIP > 0.5 variants regardless of whether they are in a CS. For each positive variant, we selected 10 variants in the same gene with PIP < 0.01 matched on variant type (SNP, insertion, deletion) for the negative class (**Online Methods**). For each cell type we created two training datasets, one from fine-mapped variants from the *MEGA* genes and another combining fine-mapped variants from both the *MEGA* and *Other* genes (*MEGA+Other*). We used sample weighting in order to account for the class imbalance and upweight higher PIP positive variants (**Online Methods and Supplementary Methods**). This weighting scheme ensures that the total weight for both the positive vs negative variants is the same, and positive variants with higher PIPs are given larger weights. To accurately assess model performance, the test set used a more stringent criteria: positive variants with PIP > 0.90 and 10 matched negative variants (PIP < 0.01) in the same gene (**Online Methods**). We also restricted the test set data to only include variants from *MEGA* genes in order to have a fixed test dataset for all subsequent evaluations.

We developed a pipeline to annotate all variants within FunGen-xQTL resource using a variety of resources. The annotations fall into six broad categories: 1) distance to transcription start site (TSS), 2) activity-by-contact (ABC) scores for 4 brain cell types (astrocyte, microglia, neuron, and oligodendrocytes)^23^, 3) non cell type-specific annotations, e.g. gnomAD^39^ minor allele frequency, 4) cell type-specific annotations, 5) DL-VEP scores from Enformer^15^, BPNet^18^ and ChromBPNet^17^, and 6) gene features, e.g. the GeneBayes constraint score^24^ (**Online Methods, Figure 1c**). For the Enformer annotations, we calculated DL-VEP scores for all non-CAGE assays (**Figure 1b**), to remove the circularity of finding variants that disrupt gene expression using a score meant to measure the disruption of gene expression. We also reasoned this would aid in model interpretation, investigating the local molecular features driving changes in gene expression. Enformer is trained on publicly available functional genomics data derived from large consortia such as Roadmap Epigenomics^19^ and the ENCODE^20^ project, which do not include data for AD-relevant cell types such as microglia. We therefore trained cell-specific deep learning models (BPNet^18^ and ChromBPNet^17^) predicting H3K27ac, H3K4me1, and ATAC-seq for 4 brain cell types^37^. Following the approach we used in Lakhani et al.^40^, we calculated DL-VEP scores for all variants within 1KB of a peak region for each assay (**Online Methods**).

For each cell type, we used a leave one chromosome out (LOCO) cross-validation approach to model training. Holding out each chromosome in turn, we trained a sample-weighted CatBoost model on variants in the training data from all other chromosomes (**Figure 1d and Supplementary Methods**). We evaluated model performance on the test data from the heldout chromosome. We trained models using data from just *MEGA* or *MEGA+Other*.

Unlike typical machine learning datasets, our training data has “noisy labels” since even state-of-the-art fine-mapping tools like SuSiE that take linkage disequilibrium (LD) into account can pick the wrong SNP due to challenges such as particularly complex LD, missing variants, genotype imputation errors, or low statistical power. Such errors are most common between SNPs in high LD, which tend to be proximal in the genome. At the same time, many annotations (e.g. distance to TSS, ABC scores) are highly autocorrelated, meaning that proximal SNPs will have similar values. By contrast DL-VEP do not have substantial autocorrelation^15^, in principle making them ideal for distinguishing the causal variant out of a set of variants in high LD. However, the model may overly weigh features whose autocorrelation matches the LD-induced correlation in the labels. To account for this possibility, we trained a model where we upweighted all DL-VEP features — which are more dependent on local sequence context than LD correlation and therefore more robust to confounding between causal variants and their proxies — to encourage the model to prioritize these highly informative features (**Supplementary Methods**).

### Upweighting deep learning features better localizes AD polygenic signal

The models with and without DL-VEP upweighting had similar test AUPRC (**Supplementary Figures 1-6**). However, we tested whether the DL-VEP upweighted or DL-VEP unweighted model had better discriminative abilities in an external bulk cell-sorted microglia eQTL dataset called MIGA^41^. The MIGA dataset contained both genotyping array and bulk cell-sorted microglia RNA-seq measurements from 4 brain regions for 100 individuals. We created two increasingly stringent evaluation datasets by selecting fine-mapped eQTL variants that replicated across multiple brain regions: 1) variants with PIP > 0.25 in at least two brain regions (n=46), and 2) variants with PIP > 0.50 in at least two brain regions (n=9) (**Supplementary Figure 7a-b**). At the PIP > 0.25 threshold we found 41.3% of the variants had a predicted probability > 0.25 using the DL-VEP upweighted model compared to only 36.9% of variants using the DL-VEP unweighted model (**Supplementary Figure 7a**). At the more stringent threshold (PIP > 0.50) we found 66.6% of variants had predicted probability > 0.25 using the DL-VEP upweighted model compared to only 55.5% of variants using the DL-VEP unweighted model (**Supplementary Figure 7b**). Also, the DL-VEP unweighted model assigned more putative causal eQTLs (based on MIGA fine-mapping) a predicted probability near zero, incorrectly classifying them as non-causal (i.e., producing more false negatives). We conclude the DL-VEP upweighted model better identifies true causal eQTLs by reducing false negative predictions, and therefore used this model for all subsequent analyses.

We also investigated how well both the microglia DL-VEP upweighted and DL-VEP unweighted models explain heritability and enrichment of heritability for AD GWAS. For each model, we constructed binary functional annotations where each variant received a value of 1 if the variant was a predicted eQTL with probability >= p for any gene, and 0 otherwise. We generated 20 annotations per model using probability thresholds ranging from p = 0.80 to p = 0.99 in increments of 0.01. We used stratified LD score regression^42^ (SLDSC) to calculate the proportion of heritability (𝑝_*h*^2^_) and enrichment of heritability explained (𝑒_*h*^2^_) by each annotation (**Online Methods**). Throughout our subsequent analyses, we use this binary annotation framework with varying probability or PIP thresholds to assess heritability contributions.

When comparing the Pareto frontiers of 𝑝_*h*^2^_ versus 𝑒_*h*^2^_ the DL-VEP upweighted model dominated the DL-VEP unweighted model (**Supplementary Figure 8**). Specifically, for every 𝑝_*h*^2^_

achieved by the DL-VEP unweighted model at a given probability threshold, the DL-VEP upweighted model achieved a similar 𝑝_*h*^2^_ (possibly at a different threshold) but with higher enrichment of heritability, thus better localizing the polygenic signal from the AD GWAS. Given that the DL-VEP upweighted model better identifies true eQTLs in the MIGA dataset and achieves superior polygenic localization in the AD GWAS, we use this model for all subsequent analyses and refer to it simply as scEEMS.

### scEEMS outperforms Distance to TSS in predicting cell type eQTLs

As expected, the number of variant-gene pairs available for model training was heavily dependent on the cell type abundance in the underlying single cell datasets. This results in large variability in training data size, the smallest being for microglia (3810 positive variants from 801 genes) and the largest for excitatory neurons (32,090 positive variants from 5,843 genes) (**Figure 2a**). Correspondingly, the number of variants and genes in the test set was also smallest in microglia (173 positive variants from 157 genes) and largest in excitatory neurons (1990 positive variants from 1691 genes) (**Figure 2b**). To assess performance we obtained the predicted probability for each test variant under the model where its chromosome was held out.

**Figure 2:**
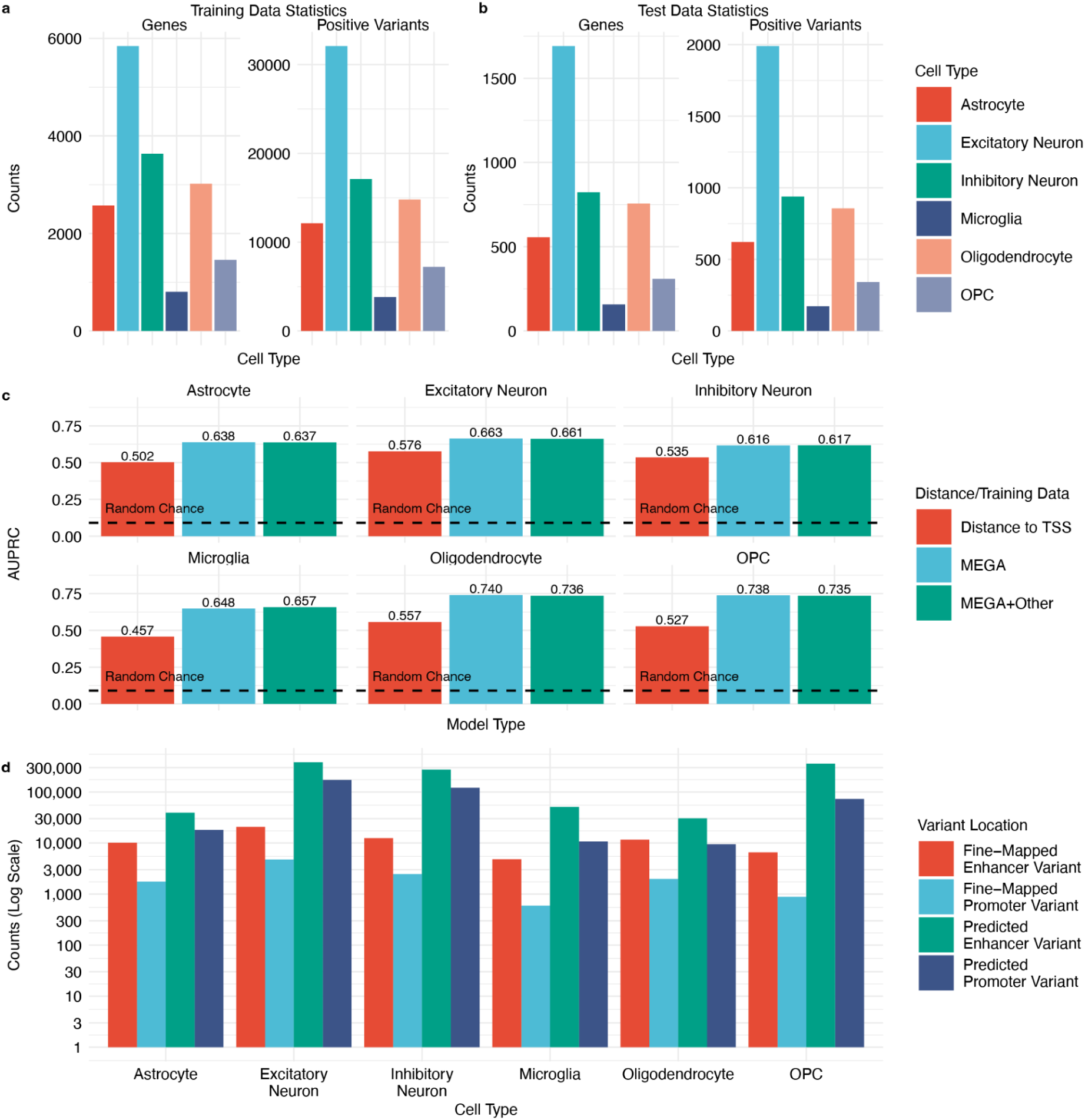
**Training statistics and variant discovery across brain cell types** a) Number of eQTLs and eGenes used as training data for each cell type scEEMS model. b) Number of eQTLs and eGenes used as testing data for each cell type scEEMS model. c) AUPRC for cell-specific prediction of eQTLs using just distance to TSS (log transformed), scEEMS trained on MEGA genes only, and scEEMS trained on combined MEGA+Other genes. Dashed line represents the AUPRC based on random chance. d) Number of fine-mapped eQTLs (PIP > 0.10) and predicted eQTLs (at cell-specific thresholds) identified using scEEMS. Stratified by whether the variant is considered a promoter (within 10KB of TSS) or enhancer.

We aggregate all test predictions to calculate a single AUPRC per cell type (this specific cross-validation based scoring is referred to as *prevalidation*). In EMS^21^, it was found that distance to TSS is a strong predictor so we compare to a baseline which predicts eQTL status as a function of distance from the gene TSS alone. Model training using *MEGA* generally performed similarly to using *MEGA+Other* (**Figure 2c**). Both models performed much better than the simple baseline model (**Figure 2c**). The best performance was for oligodendrocytes (AUPRC_Baseline_ = 0.557, AUPRC_MEGA_ = 0.740, AUPRC_MEGA+Other_ = 0.736) and the worst performance was in inhibitory neuron (AUPRC_Baseline_ = 0.535, AUPRC_MEGA_ = 0.616, AUPRC_MEGA+Other_ = 0.617). Since there was no noticeable boost in test AUPRC from using the *MEGA+Other* training data, we used the model trained only on MEGA data for all subsequent analyses.

A key challenge was to determine a meaningful prediction probability threshold for determining what variant-gene pairs are predicted eQTLs. Similar to the DL-VEP weighting analysis, we used the scEEMS models to create multiple cell-specific binary annotations across a range of predicted probability thresholds (p = 0.80 to p = 0.99) (**Online Methods**). For each cell type, there was no clear cut universal prediction threshold that had both desirable characteristics of high 𝑝_*h*^2^_ and 𝑒_*h*^2^_ (**Supplementary Figures 9-14**). Invariably, choosing a more stringent probability threshold would prioritize variants which would have higher 𝑒_*h*^2^_ but at the expense of explaining less of the total AD GWAS heritability (𝑝_*h*^2^_). We took a data-driven approach where, for each cell type, we selected the prediction probability threshold with the highest

per-standardized-annotation effect size^43^ (τ*) (**Online Methods**). The τ* metric balances finding annotations with high 𝑒_*h*^2^_ against having a larger proportion of variants in the annotation, which increases 𝑝_*h*^2^_. Based on this criterion we identified different prediction probability thresholds for each cell type (p_Ast_ = 0.97, p_Exc_ = 0.94, p_Inh_ = 0.94, p_Mic_ = 0.95, p_Oli_ = 0.98, and p_OPC_ = 0.91) (**Supplementary Figure 15**).

Using these thresholds, there was an average 20.9-fold increase of predicted eQTLs compared to fine-mapped eQTLs. Following Mostafavi et al.^4^, we classified all variants within 10KB of the gene TSS to be promoter-like and all beyond 10KB to be an enhancer-like variant. Among all cell types, there was an average 33.6-fold increase in the number of promoter-like variants identified and an average 18.7-fold increase in the number of enhancer variants identified (**Figure 2d**).

### Deep learning features collectively contribute more to model predictions than distance to TSS

We calculated SHAP^30,31^ values for all variants predicted to be an eQTL (based on their cell type-specific thresholds) in order to determine the features driving the model prediction (**Online Methods**). As expected from the EMS^21^, across all cell types, the distance feature had the largest SHAP value for both promoter-like and enhancer-like variants. However, collectively, the sum of the absolute value of Enformer SHAP values contributes more than the distance feature (**Figure 3a**).

**Figure 3:**
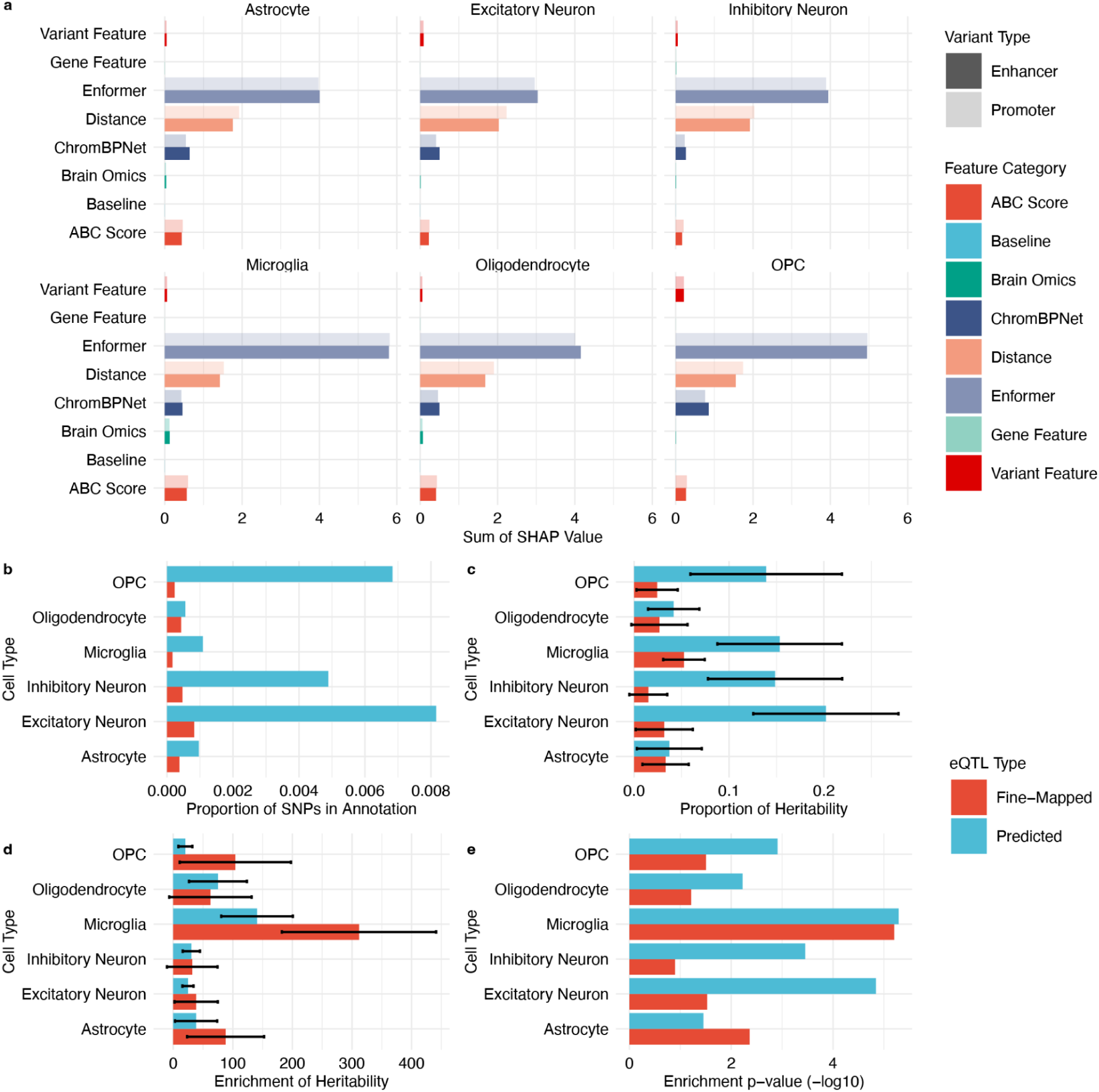
**Feature importance and heritability contributions of predicted versus fine-mapped eQTLs** a) Sum of absolute SHAP values for feature categories across six brain cell types, separated by promoter-like and enhancer-like variants. SLDSC heritability analysis comparing binary annotations from fine-mapped eQTLs (PIP > 0.10) versus predicted eQTLs (cell-type-specific thresholds): b) Proportion of variants. c) Proportion of AD GWAS heritability explained (𝑝_*h*^2^_). d) Enrichment of heritability (𝑒_*h*^2^_). e) Statistical significance of enrichment (p-value).

### Predicted eQTLs explain more to AD GWAS heritability than fine-mapped eQTLs

We tested whether predicted eQTLs could explain more AD GWAS heritability compared to fine-mapped eQTLs. We generated cell type-specific binary annotations from both fine-mapped and predicted eQTLs to estimate their contribution to AD heritability. For fine-mapped eQTLs, we created binary annotations by assigning a value of 1 to variants with PIP > 0.10 for any *MEGA+Other* gene and 0 otherwise. For predicted eQTLs, we assigned a value of 1 if the variant’s predicted probability exceeded its cell-type-specific threshold, and 0 otherwise (**Online Methods**). We used SLDSC^42^ to estimate both the enrichment and proportion of heritability explained by these annotations in the Bellenguez AD GWAS. Across all cell types, fine-mapped eQTLs showed higher 𝑒_*h*^2^_ but explained only a small proportion of 𝑝_*h*^2^_ (**Figure 3c-d**). In contrast, predicted eQTLs explained substantially more trait heritability, with excitatory neurons (𝑝_*h*^2^_ = 0.20) and microglia (𝑝_*h*^2^_ = 0.15) being the largest contributors (**Figure 3c-d**). Notably, predicted microglia eQTLs maintained high enrichment of heritability (𝑒_*h*^2^_ = 140.6) while capturing this larger proportion of trait heritability.

### Priors from predicted eQTLs can improve fine-mapping and explain more AD GWAS heritability

Similar to the EMS analysis^21^, we performed single cell eQTL fine-mapping using the held out scEEMS prediction probability scores as priors within the SuSiE pipeline (**Online Methods**).

We recalibrated the prior predicted probabilities based on the most recent GTEx analysis^1^ which found, for the better powered tissues, there were an average of 1.8 cis-eQTLs per gene. In our analysis we tested, on average, 14,716 cis-variants per gene resulting in a prior probability of being a cis-eQTL to be 1.8/14,716 = 0.00012. The current scEEMS model, based on the class weighting scheme, assumes the prior probability of being a variant being an eQTL is 50%. We recalibrated the predicted probabilities from scEEMS using this more realistic probability (prior probability = 0.00012) of a variant being an eQTL (**Supplementary Methods**). We then re-ran the fine-mapping pipeline on all MEGA genes using both a uniform prior and a prior using the recalibrated predicted probability (scEEMS prediction) and kept all CS - gene pairs with a minimum absolute correlation (purity) above 0.50. We restricted our analysis to the microglia and astrocyte cell types. We found a net increase in discovered eGenes when using the scEEMS prediction prior (n_Mic_ = 107, n_Ast_ = 271) compared to the uniform prior as well the net number of genes with an increase in independent CS (n_Mic_ = 147, n_Ast_ = 428) which represent independent genetic signals for each gene (**Figure 4a**). We also ranked each CS based on their highest PIP value (e.g. the CS with highest PIP value gets rank 1) and then paired the CS found with the uniform and scEEMS prediction prior based on their gene and CS ranking. For each pair of CS, on average, the top PIP value was higher for the scEEMS prediction CS with a mean PIP increase of 0.169 (t-test p-value: 1.8x10^-58^) for microglia and 0.111 (t-test p-value: 2.5x10^-58^) for astrocyte. For all genes, we calculated the mean purity of CSs within that gene and found an increase in mean purity of 0.006 (t-test p-value: 0.317) for microglia and 0.01 (t-test p-value: 0.003) for astrocytes when using the scEEMS priors, with the non-significant result in microglia likely attributable to the minimum purity threshold already filtering for high-quality credible sets (**Supplementary Figure 16a**). We also found the mean CS size per gene also decreased with an average decrease of 6.26 variants (t-test p-value: 0.03) for microglia and 5.28 variants (t-test p-value: 0.001) for astrocytes when using the scEEMS prediction prior (**Supplementary Figure 16b**). Finally, while not significant overall, we found the mean log Bayes factor (lbf) significantly increased for genes where the mean lbf was low with respect to fine-mapping with the uniform prior (mean lbf < 10). The mean lbf increased by 1.85 (t-test p-value: 5.4x10^-17^) for microglia and 2.02 for astrocytes (t-test p-value: 5.1x10^-49^) (**Supplementary Figure 16c**). Based on comparisons of fine-mapping using both a uniform and scEEMS prediction prior we conclude that eQTL fine-mapping with a scEEMS prior aids in both finding additional eGenes and independent statistical signals not found through fine-mapping with a uniform prior. Fine-mapping with the scEEMS prior also improves localization of the credible set increasing our ability to find the true causal eQTL, especially in settings where the statistical evidence (e.g. CS with low lbf) is less clear. This is of particular importance in single cell eQTL datasets where a rare cell type is of disease importance such as the case of microglia for AD.

**Figure 4:**
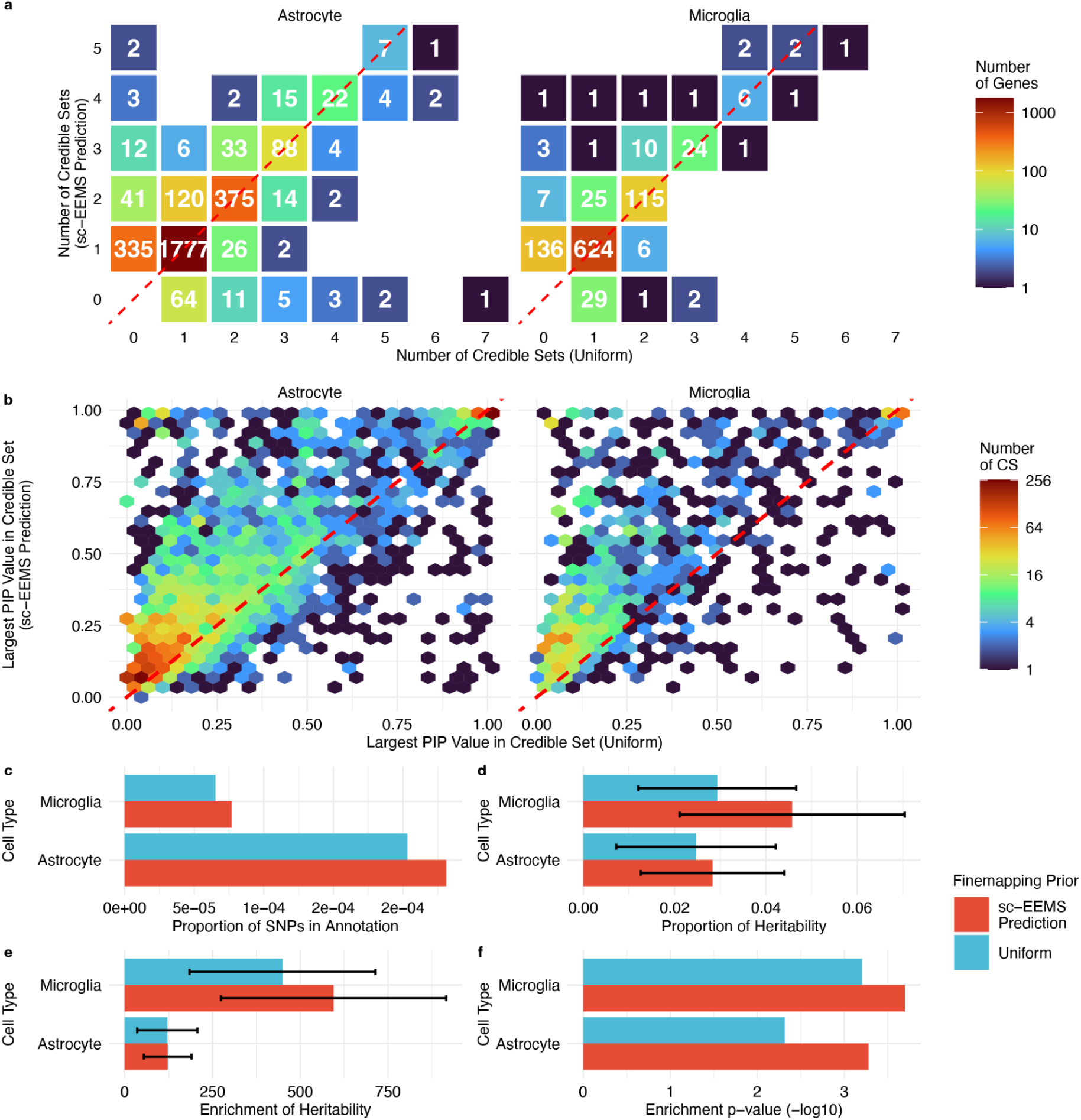
**eQTL fine-mapping performance with scEEMS versus uniform priors** a) Number of credible sets per gene comparing scEEMS prediction prior versus uniform prior. b) Highest PIP values for credible sets matched by gene and rank, comparing scEEMS prediction prior versus uniform prior. SLDSC heritability analysis of binary annotations from fine-mapped eQTLs (PIP > 0.10) using scEEMS prediction versus uniform priors: c) Proportion of variants. d) Proportion of AD GWAS heritability explained (𝑝_*h*^2^_). e) Enrichment of heritability (𝑒_*h*^2^_). f) Statistical significance of enrichment (p-value).

We also compared whether the fine-mapped variants found using the scEEMS prediction prior aided in explaining additional AD GWAS heritability. Similar to our previous analysis, we created binary functional annotations based on the fine-mapped eQTLs and then used SLDSC to estimate both AD GWAS 𝑝_*h*^2^_ and 𝑒_*h*^2^_. In particular, for each cell type and fine-mapping method, we created a binary functional annotation where a variant with PIP > 0.10 in any *MEGA* gene was assigned a value of 1 otherwise it was assigned a value of 0. We compared the annotations found using the uniform and scEEMS prediction priors and found the scEEMS prediction prior approach had a larger proportion (prop) of variants in the annotation (prop_Mic_ = 7.68x10^-5^, prop_Ast_ = 2.34x10^-4^) compared to the uniform prior (prop_Mic_ = 6.51x10^-5^, prop_Ast_ = 2.03x10^-4^) (**Figure 4c**). The annotations using the scEEMS prediction priors also captured more AD heritability 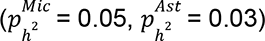 compared to the uniform prior 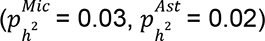 (**Figure 4d**). The scEEMS predicted prior also had higher enrichment of heritability 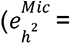 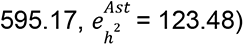 compared to the uniform prior 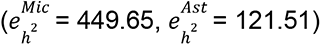 (**Figure 4e**).

This shows that annotations using the scEEMS prediction prior, especially in microglia, uncover more fine-mapped eQTLs which explain AD GWAS heritability as well as better localize the AD polygenic signal.

### Predicted eQTLs nominate more putative AD risk genes than fine-mapped eQTLs

The increased proportion of heritability and high enrichment observed for predicted eQTLs, combined with their predicted variant-gene linkages, provides an opportunity to discover new AD risk genes. To identify cell-specific putative AD risk genes, we used eMAGMA^32^ to integrate our predicted and fine-mapped (uniform prior) cell-specific variant-gene linkages (*MEGA+Other Genes*) with summary statistics from the Bellenguez AD GWAS (**Online Methods**). We also performed standard MAGMA analysis, which maps all variants within 10KB of a gene body to that particular gene, as a baseline. We found more putative AD risk cell type-gene pairs (215) and unique genes (111) using predicted eQTLs (**Supplementary Table 3**) compared to fine-mapped eQTLs (76 cell type-gene pairs, 55 genes) (**Supplementary Table 4**). As an additional control, we created a k-nearest neighbor (kNN) eMAGMA baseline where each gene was assigned its k nearest variants (matching the number of variants for that gene in the respective predicted or fine-mapped eQTL dataset) to control for gene-specific variant density effects (**Online Methods**). This kNN eMAGMA baseline resulted in fewer significant genes for both predicted eQTLs (n_cell type-gene_ = 98, n_unique_ = 41) and fine-mapped eQTLs (n_cell type-gene_ = 32, n_unique_ = 17). The predicted microglia eQTLs resulted in the largest percentage (0.58%) of testable genes identified as AD risk genes of which 44% are not found by the standard MAGMA analysis (**Figure 5a and Supplementary Figure 17**). Among the 215 cell type-gene pairs identified by eMAGMA with scEEMS, 23 were microglia-specific followed by 12 that were excitatory neuron-specific (**Figure 5b and Supplementary Figure 18**).

**Figure 5:**
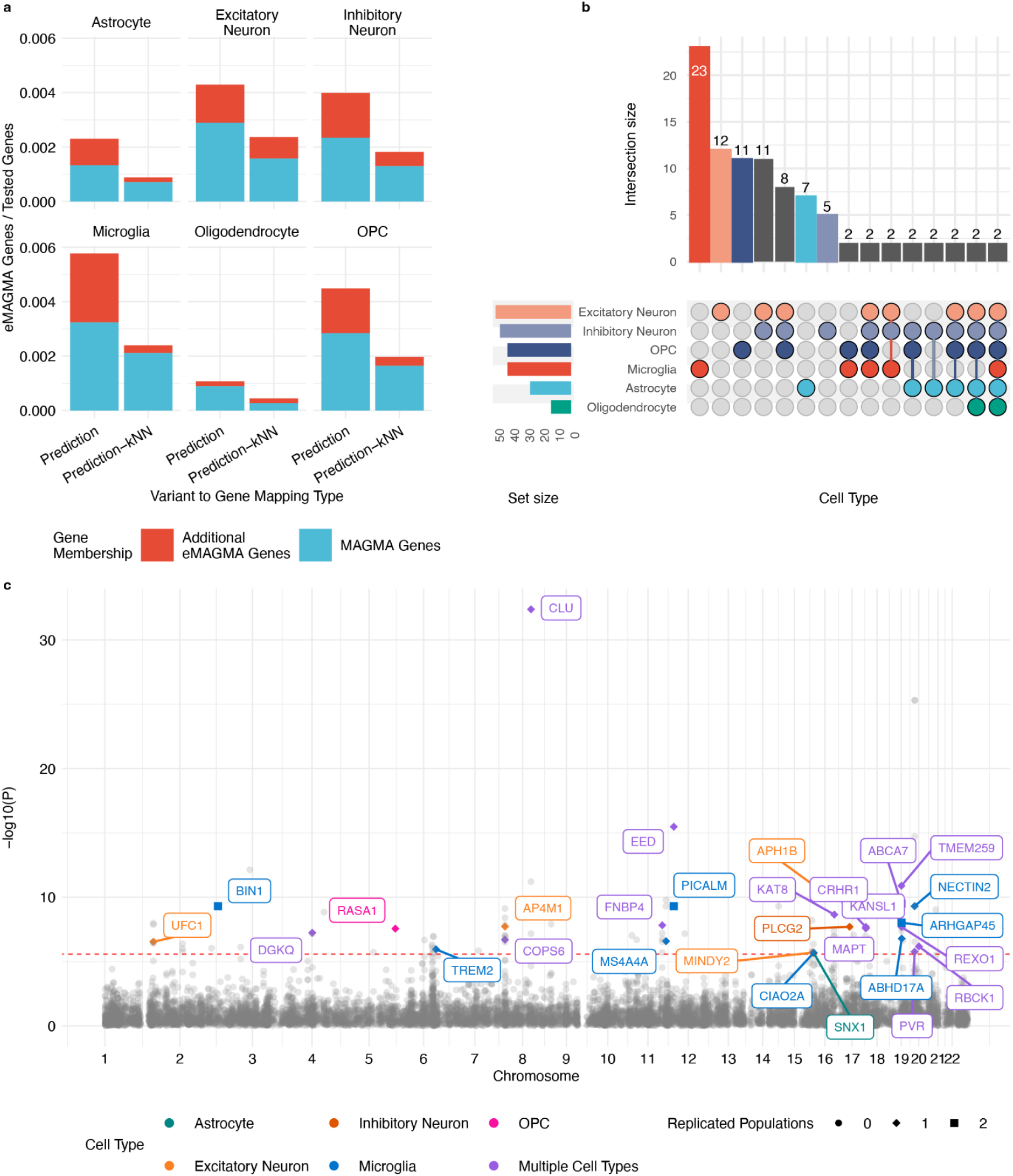
**Cell type-specific Alzheimer’s disease risk gene discovery** a) Percentage of significant eMAGMA genes (Bonferroni-corrected P < 2.67×10⁻⁶) using scEEMS predicted eQTLs for each cell type and a gene-matched kNN baseline. Blue bars: genes also found by standard MAGMA; red bars: eMAGMA-specific genes. b) Cell-type membership of all significant scEEMS predicted eMAGMA genes (UpSet plot; sets of size 1 excluded for clarity; see Supplementary Figure 18 for complete data) c) Manhattan plot of significant scEEMS predicted eMAGMA genes in the European AD GWAS. Gray: no replication in non-European populations; colored: replicated (P < 0.05) in non-European populations. Circles: zero replicated populations; squares: one replicated population; diamonds: two replicated populations. Color indicates cell type.

We also conducted the same eMAGMA analysis on AD GWAS summary statistics from African American (AFR), Hispanic (AMR), and East Asian (EAS)^33^ populations. We sought to find eMAGMA risk genes found from the EUR AD GWAS which replicated in an AD GWAS from a non-European population^33^ (**Online Methods**). Of the 215 cell type-gene pairs found from the predicted eQTLs, 62 replicated in a non-European population including 31 between the EUR/AMR populations, 13 between the EUR/AFR populations, and 13 between the EUR/EAS populations (**Supplementary Figure 20**). We also found a total of 5 cell type-gene pairs replicating across EUR and 2 non-EUR populations (**Supplementary Figure 20**). In contrast, for the fine-mapped eQTLs, only 29 cell type-gene pairs replicated between a European and non-European population, including 17 cell type-gene pairs between the EUR/AMR populations, 5 between the EUR/AFR populations, and 5 between EUR/EAS populations (**Supplementary Figure 21**). The 5 cell type-gene pairs that replicated across 2 non-European ancestries were *BIN1* (Microglia), *PICALM* (Microglia), *ABCA7* (Astrocyte and Oligodendrocyte), and *ARHGAP45* (Microglia) (**Figure 5c**). *BIN1*, while broadly expressed in the brain, has a well-known AD risk SNP located in what is thought to be a microglia-specific enhancer^44^. *PICALM* is another AD risk gene which is thought to disrupt lipid metabolism in microglia^45^. Recent analysis has demonstrated a strong AD GWAS risk variant functions as a *PICALM* eQTL in microglia^45^. *ABCA7* is another well-known AD risk gene which has been known to have high expression in astrocytes and oligodendrocytes^46^. The exact mechanism for *ABCA7*’s influence is not known but it is believed to play roles in AD-relevant processes such as lipid metabolism, phagocytosis, and amyloid deposition^46^. The connection between *ARHGAP45* in microglia and Alzheimer’s is not currently understood. Mendelian randomization approaches have linked *ARHGAP45* to have a protective effect in Alzheimer’s disease^47,48^ but was found to not replicate or had inconsistent direction of effects. The distance between the TSSs of *ARHGAP45* and ABCA7 is only ∼25KB therefore, another possibility is the effect attributed to *ARHGAP45* is actually due to *ABCA7*.

Of the 62 replicated cell type-gene pairs (based on predicted eQTLs) we found 18 which were not MAGMA or fine-mapped eMAGMA cell type-gene pairs, representing 10 novel AD specific risk genes only found through scEEMS (**Supplementary Figure 22**). Of these cell type-gene pairs, 6 are specific to one cell type with the top cell types being microglia (2) and OPC (2).

These scEEMS specific genes are *FNBP4*, *SNX1*, *RBCK1*, *TMEM259*, *UFC1*, *COPS6*, *CIAO2A*, *ABHD17A*, *EED*, and *RASA1* (**Supplementary Figure 23**).

### scEEMS provides unique insights about AD risk in the *PICALM*, *SNX*, and *BLNK* loci

Finally, we demonstrate three interesting phenomena enabled by scEEMS through an in-depth analysis of the *PICALM*, *SNX1*, and *BLNK* loci. In the Bellenguez AD GWAS, the *PICALM* locus contains multiple variants of interest. This behavior is illustrated by the *PICALM*/*EED* locus where two intergenic variants (*rs10792832* and *rs3851179*) are in almost perfect LD and each receives PIP ∼0.5 in statistical fine-mapping of the Bellenguez GWAS^40^ (**Supplementary Figures 36-37**). In Lakhani et al.^40^, using functional informed fine-mapping with deep learning annotations, it was found that *rs10792832* is the likely causal AD risk variant with an Enformer DL-VEP score suggesting the variant disrupts *PU.1* TF binding (**Supplementary Figure 37**). We reasoned *rs10792832* is an eQTL in microglia, which confers risk through modulation of gene expression due to decreased binding affinity of the *PU.1* TF. However, eQTL fine-mapping returns *rs10792832* and *rs3851179* in a credible set for *PICALM*, but *rs3851179* has a higher PIP (PIP = 0.90) than *rs10792832* (PIP = 0.09). Statistical eQTL fine-mapping likely misidentified the causal eQTL in this case (**Supplementary Figure 37**). The scEEMS model predicted *rs3851179* (probability = 0.75) had a lower probability compared to *rs10792832* (probability = 0.97). Recent experimental validation^45^ confirmed that *rs10792832* functions as a *PICALM* eQTL in microglia and disrupts *PU.1* TF binding as predicted by our analysis, demonstrating the utility of scEEMS for identifying functionally relevant variants. Additionally, we identified an independent secondary signal in the *PICALM* region which replicated in both AMR and EAS populations (**Supplementary Figure 38**), suggesting multiple causal variants may contribute to AD risk at this locus.

Through our eMAGMA analysis, we found the gene *SNX1* to be a strong astrocyte-specific AD risk gene. While *SNX1*-associated variants are not GWAS significant in the Bellenguez AD GWAS, the gene shows statistically significant association through our eMAGMA framework and this finding replicates in the EAS AD GWAS (**Supplementary Figure 39**). This demonstrates how scEEMS can identify cell type-specific risk genes that may be missed by standard GWAS approaches, particularly for variants with more modest effect sizes that become detectable when linked to their appropriate cellular context through predicted eQTLs. Analysis of older AD GWAS^49^ has shown that variants in the *SNX1* locus are nominally significant for AD risk but none have implicated the scEEMS predicted astrocyte eQTL (*rs332258*).

Finally, through our eMAGMA analysis, we found *BLNK* to be a significant microglia eMAGMA gene based on the European AD GWAS (eMAGMA p-value: 2.04×10⁻⁷). Recent colocalization analysis^28^ has also identified this gene as a microglia AD risk gene. However, as reported in that analysis, this locus is quite complex, as many variants are in high LD; therefore, it is difficult to determine the number of independent genetic signals that contribute to AD risk (**Supplementary Figure 40a**). Fine-mapping the microglia *BLNK* eQTLs using the uniform prior identified two independent credible sets. The first CS (CS1) contains only one variant (*rs1870170*), whereas the second CS (CS2), which contains the reported AD risk variant *rs6584063*, includes multiple variants with a top PIP value of 0.12 (**Supplementary Figure 40b**). Using the scEEMS prediction prior, one variant in CS2 (*rs1870171*) instead reaches PIP > 0.30 (**Supplementary Figure 40c**), primarily driven by its high scEEMS prediction probability (0.97; **Supplementary Figure 40d**). Interestingly, *rs1870170* and *rs1870171* are separated by only 4 bp but are not in high LD (r² = 0.0008)⁵⁰, further supporting the assumption that these represent independent signals. We also found that *rs1870170* is protective for AD risk and decreases *BLNK* expression, whereas *rs1870171* increases AD risk and increases *BLNK* expression (**Supplementary Table 6),** suggesting that increased *BLNK* expression confers increased AD risk.

We investigated DL-VEP scores for *rs1870170*, *rs1870171*, and *rs6584063* and found high TF DL-VEP scores for *rs1870171* (*PU.1*) and *rs6584063* (*SP1*) and a moderate score for *rs1870170* (*NRF1*) (**Supplementary Figure 41b**). However, *rs1870171* has the highest ChromBPNet DL-VEP score, followed by *rs1870170*, with a negligible score for *rs6584063* (**Supplementary Figure 41c**). Using functional genomics data, we found that *rs1870170* and *rs1870171* are located within ∼900 bp of an open chromatin peak, whereas *rs6584063* is 2,627 bp from the nearest open chromatin region (**Supplementary Figure 41d**). The microglia ABC score linking *rs1870170* and *rs1870171* to *BLNK* is close to 0.04, whereas *rs6584063* has an ABC score of 0.

We conducted an in-depth analysis of ChromBPNet microglia predictions and found that, relative to the reference sequence, *rs1870170* decreases chromatin accessibility while *rs1870171* increases chromatin accessibility (**Supplementary Figure 42a–c**), concordant with the microglia *BLNK* eQTL effect sizes. Using deep learning model interpretability tools^50,51^, we found that the strongest model attribution scores map to a *PU.1* motif within the microglia open chromatin region for both reference and alternate sequence predictions (**Supplementary Figure 42d–f**). Integrating both DL-VEP scores and functional genomics data, we hypothesize that *rs1870170* and *rs1870171* likely each independently modify microglia chromatin accessibility, potentially altering *BLNK* expression in opposite directions.

## Discussion

We developed single cell Enhanced Expression Modifier Scores (scEEMS) to predict putative causal eQTLs in pseudobulk single cell types. A distinctive feature of scEEMS is the modified training procedure which can accommodate lower powered eQTL datasets. This procedure can aid in finding more enhancer-like eQTLs^4^ in specific cell contexts^5^ (typically found in single cell eQTL datasets) which are likely to be disease relevant. This is of particular importance in AD where it is believed that much of the polygenic signal is found in cis-regulatory regions (CRE) of a rare cell type called microglia^37,52^. The scEEMS models have improved predictive capability to detect eQTLs compared to the standard method of mapping a variant to its nearest gene. The models utilize deep learning variant effect prediction scores to better discriminate between causal and non-causal eQTLs and ABC scores to aid in variant to gene linkages. While scEEMS has learned long range variant-gene links using the ABC score feature, the model still has difficulty in discriminating between variant-gene pairs in complex enhancer regions which are linked to multiple genes. This can be attributed to the fact that standard ABC scores^22,23^ use an average Hi-C profile from multiple cell types to make enhancer-gene linkages. In future analysis, we will investigate the ability of adding features derived from enhancer-gene linkage methods based on single cell multiome data^53^ and cell-specific chromosome conformation^37^ to better discriminate variant-gene linkages in such enhancer regions.

### Beyond prediction alone, scEEMS also serves as a valuable tool for enhancing eQTL

fine-mapping, particularly in single-cell datasets from rare cell types where statistical power is limited. A fundamental challenge in linking AD GWAS signals to regulatory mechanisms is the substantial sample size disparity between our eQTL cohort (788 individuals) and the AD GWAS (nearly 800,000 individuals). This disparity, combined with the rarity of disease-relevant cell types like microglia, makes it particularly difficult to detect rarer variant eQTLs that colocalize with AD GWAS signals. By using scEEMS predicted probabilities as priors in fine-mapping, we can overcome these limitations. We demonstrated substantial improvements in both discovery and localization of causal eQTLs in microglia and astrocytes, with the scEEMS prediction prior leading to discovery of additional eGenes and independent credible sets not found with uniform priors, while also increasing the posterior inclusion probabilities of likely causal variants and reducing credible set sizes. These improvements were especially pronounced for genes with weaker statistical evidence, where scEEMS priors helped discriminate true signals from noise. Through in-depth analysis of the complicated *BLNK* locus, the scEEMS approach helped us identify two independent microglia eQTL signals which colocalize with AD risk variants.

Integration with DL-VEP revealed potential mechanisms including modulation of microglia chromatin accessibility through disruption of *PU.1* binding sites. More broadly, this integration of prediction and mechanistic interpretation addresses a key challenge in complex trait genetics: establishing gene regulatory network connections from common variation, which recent studies have shown to be particularly difficult for complex traits^54^. Our approach uncovered disruption of *PU.1* binding at two separate AD risk genes, *PICALM* and *BLNK*, revealing parts of the microglia gene regulatory network that modulate AD risk. In the *BLNK* locus, our model also discovered two variants with antagonistic effects on the same open chromatin region in microglia, concluding that an increase of *BLNK* expression in microglia increases AD risk.

scEEMS has aided in the discovery of additional putative causal eQTLs in each cell type whether through fine-mapping using scEEMS predictions as priors or simply using model predictions. In both cases, these additional variants explain AD GWAS heritability that was not captured by fine-mapped eQTLs alone. For rare cell types such as microglia, which is thought to be an important AD cell type, predicted eQTLs explain a larger percentage of AD GWAS heritability while also maintaining very high enrichment of heritability. However, it is known that other cell types play a role in AD pathogenesis and some subset of AD risk variants is mediated through their disruption of gene regulation in those cell types. scEEMS has implicated AD risk genes within these cell types, but predicted eQTLs from any individual non-microglial cell type were not a significant annotation when predicted microglia eQTLs were also included as an annotation. This is partly because microglia is a rarer cell type resulting in fewer *MEGA* or *Other* genes. Our scoring procedure only predicted eQTLs in known *MEGA* or *Other* genes for that cell type which results in higher enrichment. More abundant cell types, in contrast, will have more *MEGA* or *Other* genes resulting in more predicted eQTLs as well which partly explains the lower enrichment of heritability of these annotations. In future analysis, we plan to develop cell type-specific annotations that include predicted eQTLs for eGenes with membership in certain gene programs^55^ of AD relevance. Quantifying both the heritability explained and enrichment of heritability of such cell type-specific pathways can aid in the discovery of the cell type-specific biological processes important in AD disease biology.

Finally, predicted eQTLs found by the scEEMS model enabled the discovery of more putatively causal cell type-specific AD risk genes compared to just fine-mapped eQTLs. We utilized AD GWAS summary statistics from non-European populations in order to replicate our findings using other cohorts. Using scEEMS, we found 62 cell specific AD risk genes discovered in a European cohort replicated in a non-European cohort compared to only 29 using fine-mapped eQTLs. In some cases these are genes which are considered AD risk genes in multiple cell types, raising the possibility there may be some AD risk variants which have pleiotropic effects across many cell types. In the context of AD, certain regions such as the *APOE* region have a high density of AD risk variants. In such cases, the predicted eQTLs may implicate genes that are false positives due to the difficulty of scEEMS to determine the correct variant-gene linkage in complicated enhancer regions. In future analysis, we will use scEEMS to predict rare variant eQTLs and perform rare variant association testing. We believe this will be an orthogonal method^56^ to assess cell specific AD risk genes allowing us to converge on the cell specific gene programs implicated in AD genetics.

## Data Availability

All training data resources and scEEMS model predictions will be available at https://www.synapse.org/Synapse:syn69670587 for reviewers. These resources will be made publicly available upon publication of the manuscript.

## Online Methods

### Dataset Processing and eQTL Mapping

#### Dataset Composition and Harmonization

The FunGen-xQTL consortium integrated three independent single-cell brain atlases to create the harmonized MEGA cohort: CUIMC1 (Columbia University Irving Medical Center - Batch 1)^10^, MIT (Massachusetts Institute of Technology)^9^, and CUIMC2 (Columbia University Irving Medical Center - Batch 2)^34^. All datasets were generated from post-mortem brain tissue samples from participants in the Religious Orders Study and Rush Memory and Aging Project (ROSMAP) cohort, with paired whole-genome sequencing and single-nucleus RNA-sequencing data available for 788 individuals. Cell type annotations were standardized across datasets using consistent marker genes and clustering approaches to ensure comparable cell populations for six consensus brain cell types: astrocytes, excitatory neurons, inhibitory neurons, microglia, oligodendrocytes, and oligodendrocyte precursor cells.

To maximize statistical power while avoiding sample duplication, datasets were merged sequentially following a priority-based approach:

1. All samples from CUIMC1 were retained as the reference dataset
2. Novel samples from MIT not present in CUIMC1 were identified and added
3. Novel samples from CUIMC2 not present in the combined CUIMC1+MIT cohort were identified and added

This sequential merging strategy ensured no duplicate participants were included while maximizing the final sample size for the *MEGA* cohort.

#### Pseudobulk Expression Data Processing

Raw single-cell count matrices were aggregated to pseudobulk expression profiles for each individual-cell type combination. Individuals with fewer than 10 cells per cell type were excluded to ensure sufficient cellular representation for robust pseudobulk profiles. For each source dataset independently, the following normalization steps were applied:

1. Gene filtering using the filterByExpr function from the edgeR package to retain genes with adequate expression across samples
2. TMM (trimmed mean of M-values) normalization using calcNormFactors to adjust for composition effects between samples
3. Voom transformation to convert count data to log₂ counts per million with appropriate precision weights for linear modeling

After within-dataset normalization, cross-dataset integration was performed. Only genes detected in all three source datasets were retained for downstream analysis. The removeBatchEffect function from the limma package was applied to eliminate systematic

differences between the three source datasets, treating each dataset as a separate batch. Following batch correction, genes with mean log₂CPM less than 2.0 across all samples were removed to focus on reliably detected transcripts. Finally, quantile normalization was applied to ensure consistent expression value distributions across all samples in the final merged dataset.

#### eQTL Mapping and Fine-mapping

Following data processing and normalization, genes were classified based on whether they met the mean log₂CPM ≥ 2.0 filtering criterion after combining all three datasets. Genes that fulfilled this log₂CPM criterion were designated as *MEGA* genes and underwent fine-mapping using the combined genotype matrices and expression data from all three cohorts. For genes that did not meet this threshold, fine-mapping was performed using genotype matrices and expression data from their original constituent cohorts (CUIMC1^9^ or MIT^10^) where they had sufficient expression levels. These single-cohort analyses are referred to as *Other* genes. This dual approach balanced the statistical power gained from larger sample sizes against the risk of diluting true eQTL signals for genes with insufficient expression in the combined dataset.

For each gene, expression values were residualized for covariates including batch effects, age, sex, and postmortem interval prior to fine-mapping. Statistical fine-mapping was performed using SuSiE^36^ (Sum of Single Effects) applied directly to genotype matrices (for cis-genetic variants within 1MB of gene TSS) and residualized expression phenotypes. For *MEGA* genes, SuSiE used the merged genotype matrix from all cohorts; for *Other* genes, SuSiE used the genotype matrix from the specific constituent cohort. SuSiE explicitly accounts for linkage disequilibrium patterns in the genotype data to improve causal variant localization and reduce confounding from correlated variants. The algorithm outputs credible sets (CS) containing likely causal variants for each eQTL locus, along with posterior inclusion probabilities (PIPs) for each variant representing the probability that the variant is causal for the observed eQTL signal. For subsequent analyses, we retained only credible sets with a minimum absolute correlation (purity) of 0.8 and variants not in a CS with PIP > 0.50.

#### Training Datasets for scEEMS Models

The processed eQTL mapping results were used to create two complementary training datasets for the scEEMS machine learning models. The *MEGA* dataset contained training variants derived exclusively from genes analyzed in the combined cohort, prioritizing signal robustness from larger sample sizes but with more limited gene coverage. The *MEGA+Other* dataset contained training variants from both gene categories, providing broader transcriptomic coverage at the potential cost of some analytical heterogeneity due to different source datasets and sample sizes. This dual training approach allowed assessment of the trade-off between maximizing gene coverage versus maintaining analytical homogeneity in downstream machine learning model performance.

### Variant Annotations

#### Distance

We used the gene annotation file from Ensembl to calculate the log transform of the absolute value of the distance between the variant and transcription start site (TSS) for the gene of interest.

#### ABC Scores

For each variant, we calculated the 4 ABC scores^22,23^ between each variant and gene based on the gene activity as measured by H3K27ac and ATAC-seq data^37^ for 4 neuronal cell types (astrocyte, microglia, neuron, and oligodendrocyte). Contact was measured by the averaged Hi-C data from multiple cell types (provided by the ABC codebase)^23^. All 4 ABC scores are provided to the scEEMS in order for the model to select the most appropriate ABC score model.

#### Non cell type-specific Annotations

Following EMS, we annotated each variant with values corresponding to 82 baseline annotations used by PolyFun^57^. Some of the annotations include whether a variant falls into a UCSC browser defined 5’ UTR region, coding region, intron region, or the allelic age of the variant. We also annotate each variant with its global minor allele frequency (MAF) from gnomAD^39^.

#### Cell type-specific Annotations

The cell type-specific annotations consisted of whether a variant falls into a cis-regulatory element (CRE) of a brain cell type (astrocyte, microglia, neuron, and oligodendrocyte) based on H3K27ac, H3K4me1, and ATAC-seq generated by Nott et al.^37^ within each cell type. The definition of CRE is based on definitions provided by the Nott et al. paper^37^ which include enhancers, promoters, and open chromatin regions. We also generate annotations corresponding to all combinations (union and intersection) of these regions, as well as these regions with 500 bp flanking regions.

#### Deep Learning Annotations

We used Enformer^15^, BPNet^18^, and ChromBPNet^17^ to generate deep learning variant effect prediction scores (DL-VEP) for each variant. Enformer uses ∼200KB input DNA sequence to predict 128 bp bin outputs (across the ∼200KB input sequence) for 5,313 functional genomic assays (CAGE-Seq, transcription factor ChIP-seq, histone ChIP-seq, and chromatin accessibility). For each variant, we generate a DL-VEP score for all non CAGE-seq assays by calculating the difference in model predictions between the reference and alternate sequence. The alternate sequence is simply the reference sequence centered at the alternate allele, in which we replace the reference allele with the alternate allele. For each assay, the DL-VEP score is the predicted outcome between reference and alternate allele sequences summed across the middle 32 bins (representing the 4096 bp region centered at the variant). We exclude the CAGE-seq predictions in order to both avoid the circularity of predicting gene expression modulation using a gene-expression assay feature and to be able to use model interpretation tools to determine the proximal molecular features driving gene expression changes for a particular variant. Following our approach for functionally-informed finemapping of AD GWAS^40^, we generated composite DL-VEP scores that take the maximum and minimum transcription factor DL-VEP score for a set of transcription factors that have motifs enriched in enhancer regions of a cell type^37^. We generate these composite DL-VEP scores for astroctye, microglia, neuron, and oligodendrocytes. The list of transcription factors used for each score is based on the TFs and enriched motifs found in the Nott et al. paper^37^.

We trained BPNet for H3K27ac and H3K4me1 and ChromBPNet for chromatin accessibility for astrocyte, microglia, neuron, and oligodendrocytes. These models have ∼2KB input sequence and predict the sum of log counts and base-resolution profile probabilities for the middle 1KB of the input sequence. We used the methodology validated in Lakhani et al.^40^ to calculate cell type-specific histone and chromatin accessibility DL-VEP scores for all variants. Namely, for each variant within 1KB of a cell type-specific histone or chromatin accessibility peak, the DL-VEP score is the difference in the sum of log counts between the reference and alternate sequence in which both sequences are centered at the peak, otherwise variant receives a score of DL-VEP score of zero.

#### Gene Feature

For each gene-variant pair we also use a log transformed GeneBayes score^24^, which measures gene constraint. The GeneBayes score was found to not suffer from the gene length bias commonly found in other measures of gene constraint.

### Model Training

#### Training and Test Data

Due to the difficulty of eQTL discovery in rare cell types, we used two different criteria when selecting positive class variants for the training and test data.

*Training Data.* For each cell type, positive class variants were defined as all variant-gene pairs in a credible set where the maximum posterior inclusion probability (PIP) among variants in the credible set exceeded 0.10 and where the particular variant had PIP > 0.05. We also selected variant-gene pairs not in a credible set that had PIP > 0.50 as positive class variants. For each positive class variant, we selected 10 negative class variants that had PIP < 0.01 in the same gene and were not in a credible set. The negative variant-gene pairs were matched on variant type (SNP, insertion, deletion) to account for distributional differences in features between SNPs and indels during training.

*Test Data.* For the test set, we used more stringent selection criteria to better assess model performance. Specifically, we selected positive class variants as variant-gene pairs in a credible set with PIP > 0.90. Negative class variants were selected using the same procedure as in the training data.

*Training Data Cohorts.* We assessed the benefit of adding more training data by creating two training sets. The first used training data only from variant-gene pairs where the gene was part of *MEGA* . The second combined training data from variant-gene pairs where genes could be in either *MEGA* or *Other* (referred to as *MEGA+Other*). After variant selection, we featurized each variant using the associated variant and gene features described in the ’Variant Annotations’ section.

#### Model Fitting

Once the training and test sets were determined, we used a leave-one-out chromosome (LOCO) approach for model fitting and assessment. For each cell type, we trained a model for each of the 22 chromosomes using all training variants from every chromosome except the heldout chromosome, then tested the model’s performance using test variants from the heldout chromosome. In cases where no test variants were available, we still fit a model using all training variants from the non-heldout chromosomes for downstream variant scoring.

*Sample Weighting.* For each model, we used sample weighting to upweight variants with higher PIP values, allowing the model to prioritize examples more likely to be causal eQTLs. Each of the N negative variants was given a sample weight of 1. We calculate the sum of PIP values for all N/10 positive variants 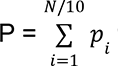 where p_i_ is the PIP for variant i. Each positive variant is given sample weight 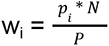. This weighting scheme provides equal balance between positive and negative variants while upweighting positive variants with higher PIP values.

*Feature Weighting.* We also considered upweighting deep learning features during model fitting. This was motivated by our recent analysis^40^ on improved fine-mapping of AD GWAS variants, where annotations prioritizing variants with high DL-VEP scores in cell-specific CRE regions helped localize the AD polygenic signal by maintaining approximately the same proportion of heritability (compared to the CRE alone) while substantially reducing the number of variants in the annotation. We defined a ’Weighted’ model where all non-deep learning features were given a feature weight f_w_ = 1 and all deep learning features were given a feature weight f_w_ = 10 **(Supplementary Methods)**.

*Model Comparisons.* We have three model types for comparison: 1) a model trained using *MEGA* with no feature upweighting, 2) a model trained using *MEGA* with deep learning feature upweighting, and 3) a model trained using *MEGA+Other* with deep learning feature upweighting. All three model types were tested on the same test set, consisting of variants from *MEGA*. For each test variant-gene pair, we calculated the predicted probability of the variant being an eQTL for that gene using the model trained with that variant’s chromosome heldout. We aggregated the predictions from all variants in the test data and then calculated a single AUPRC for each of the three model types for model comparison (a procedure known as prevalidation)

*Model Inference.* After model comparison, we selected model 2 for variant scoring. For each cell type, we selected all variants from *MEGA+Other* for that cell type and then scored each variant using the corresponding chromosome model. In sum, we generated a large database of variant-gene pairs along with a predicted probability of the variant being an eQTL for that particular gene. For downstream analysis, for each cell type, we selected a GWAS dependent prediction probability threshold based on LD score regression^42,43^. We estimated the contribution of fine-mapped and predicted eQTLs from the 6 brain cell types to the heritability of the most recent AD GWAS^11^. For each cell type, we defined a binary fine-mapped eQTL annotation, which has a value of 1 if the variant has a PIP > 0.10 for any gene in *MEGA+Other* and a value of 0 otherwise. Similarly, a predicted eQTL annotation has a value of 1 if the variant has a predicted probability > p for any gene and 0 otherwise. We generated a series of predicted binary annotations where p ranged from 0.80 to 0.99. We used different threshold criteria for these two annotation types. The relatively low PIP threshold (>0.10) for fine-mapped annotations was chosen to include enhancer-like variants, which typically have lower PIP values than promoter variants^4^. In contrast, we used stricter thresholds (0.80-0.99) for predicted eQTL annotations to minimize false positives, given that our model training procedure equally weighted positive and negative training variants which inflates prediction probabilities for variants which may be in LD with a causal eQTL. For each annotation, we use stratified LD score regression^42^ to estimate to calculate the enrichment and proportion of heritability explained by these annotations in the AD GWAS. We also include the standard baseline annotations from Polyfun^57^ as additional annotations to be used in the LD score regression model. For each cell type, we selected the probability threshold p with the highest normalized effect size 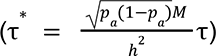, which balances annotation size and enrichment of heritability, where p_a_ is the proportion of variants in the annotation, M is the total number of variants, and h² is GWAS heritability, τ is the average per-variant heritability of the annotation. Using the criteria from Mostafavi et al.^4^ we categorized predicted eQTLs as being in a promoter-like class if they are within 10KB of the TSS and an enhancer-like class if they are further away.

### SHAP Analysis

We used the SHapley Additive exPlanations^30,31^ (SHAP) tool for model interpretation of our CatBoost models. First, we calculated SHAP scores for each predicted eQTL using its corresponding model. We assigned each feature to one of the following feature classes: ABC score, baseline, brain omics, ChromBPNet, distance, Enformer, gene feature, and variant feature. For each variant, we calculated the total contribution of each feature class to the model prediction by taking the sum of the absolute values of its SHAP scores for features in that class. Finally, for both the promoter and enhancer-like variant classes, we calculated the mean contribution across all variants in their class to estimate the relative contribution of each feature class to the model predictions.

### Fine-mapping with scEEMS prediction and uniform prior

We ran a modified SuSiE eQTL fine-mapping pipeline using raw genotype matrices and pseudo-bulk RNA-seq read counts for astrocytes and microglia. To maximize sample size, we restricted our analysis to expression data from MEGA genes, which allowed us to combine genotype and expression data from all three single-cell cohorts (CUIMC1, MIT, and CUIMC2). For each cell type and gene pair, we performed eQTL fine-mapping using either a uniform prior (no prior information) or a recalibrated scEEMS prediction-probability prior (see **Online Methods**) derived from scEEMS predictions for those genes. For each gene, the scEEMS model predictions were chromosome-specific, meaning the model was trained on variants from all chromosomes except the one containing the gene. In the SuSiE pipeline, we lowered the purity threshold from 0.80 to 0.50, retaining only credible sets (CS) where the minimum absolute correlation among variants within the set was at least 0.50.

Using these CS, we calculated the following statistics for each gene: (1) the number of CS, (2) the mean purity value of each CS, (3) the mean CS size (number of variants per CS), and (4) the mean log Bayes factor for each CS. We also compared CS across the two fine-mapping methods by first ranking all CS within a gene according to the highest PIP value in that CS (for example, the CS with the highest PIP in a gene was assigned rank 1). We then matched CS between the two methods based on both gene and rank. Next, we matched CS between methods based on both gene and rank, and recorded the highest PIP value for each matched pair.

## MAGMA Analysis

We used eMAGMA^32^ to identify AD risk genes by leveraging the cell-specific variant-gene links from both fine-mapped and predicted eQTLs and AD GWAS summary statistics to obtain cell type-specific gene-level estimates of AD risk. For a specific cell type, eMAGMA calculates a gene-level statistic of AD risk based on fine-mapped eQTLs (PIP > 0.10) or predicted eQTLs (GWAS dependent threshold) that overlap with the Bellenguez AD GWAS^11^. We identified a gene as being a cell type-specific risk gene if the p-value of the gene-level statistic was less than the Bonferroni adjusted threshold (p < 0.05/18700 = 2.67x 10^-6^). We also conducted a knn-matched eMAGMA analysis and standard MAGMA analysis as means of comparison to the eMAGMA analysis. The knn-matched eMAGMA analysis consisted of calculating the gene statistic for the n closest variants to the gene TSS, where n is the number of fine-mapped eQTLs or predicted eQTLs found for that gene. The standard MAGMA analysis maps all variants within 10KB of the genic region to the gene and then calculates the gene statistic. We also calculated these gene-level statistics using AD GWAS from African-American, Hispanic, and East Asian populations^33^ and then defined a gene as a replicated AD risk gene if it was a putative cell-specific risk gene with a p-value < 0.05 within one of these non-European populations.

**Supplementary Figure 1:**
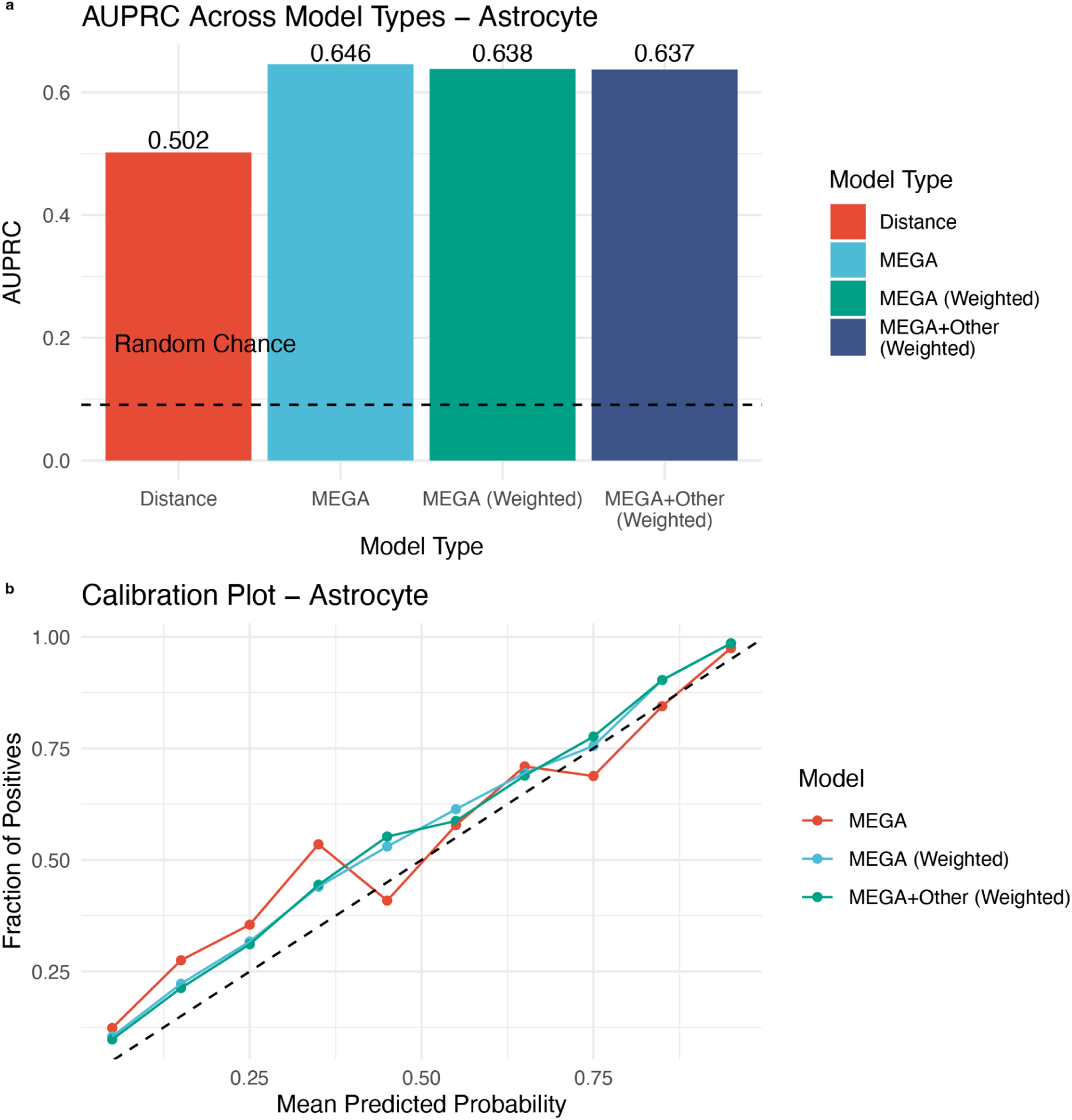
**Model performance and calibration for astrocytes** a) AUPRC values for four model variants - distance-only baseline, MEGA DL-VEP unweighted, MEGA DL-VEP upweighted, and MEGA+Other DL-VEP upweighted. Dashed line indicates expected AUPRC under random chance (0.091). b) Calibration plots showing predicted probability versus observed positive fraction across 10 probability bins. Diagonal line indicates perfect calibration.

**Supplementary Figure 2.**
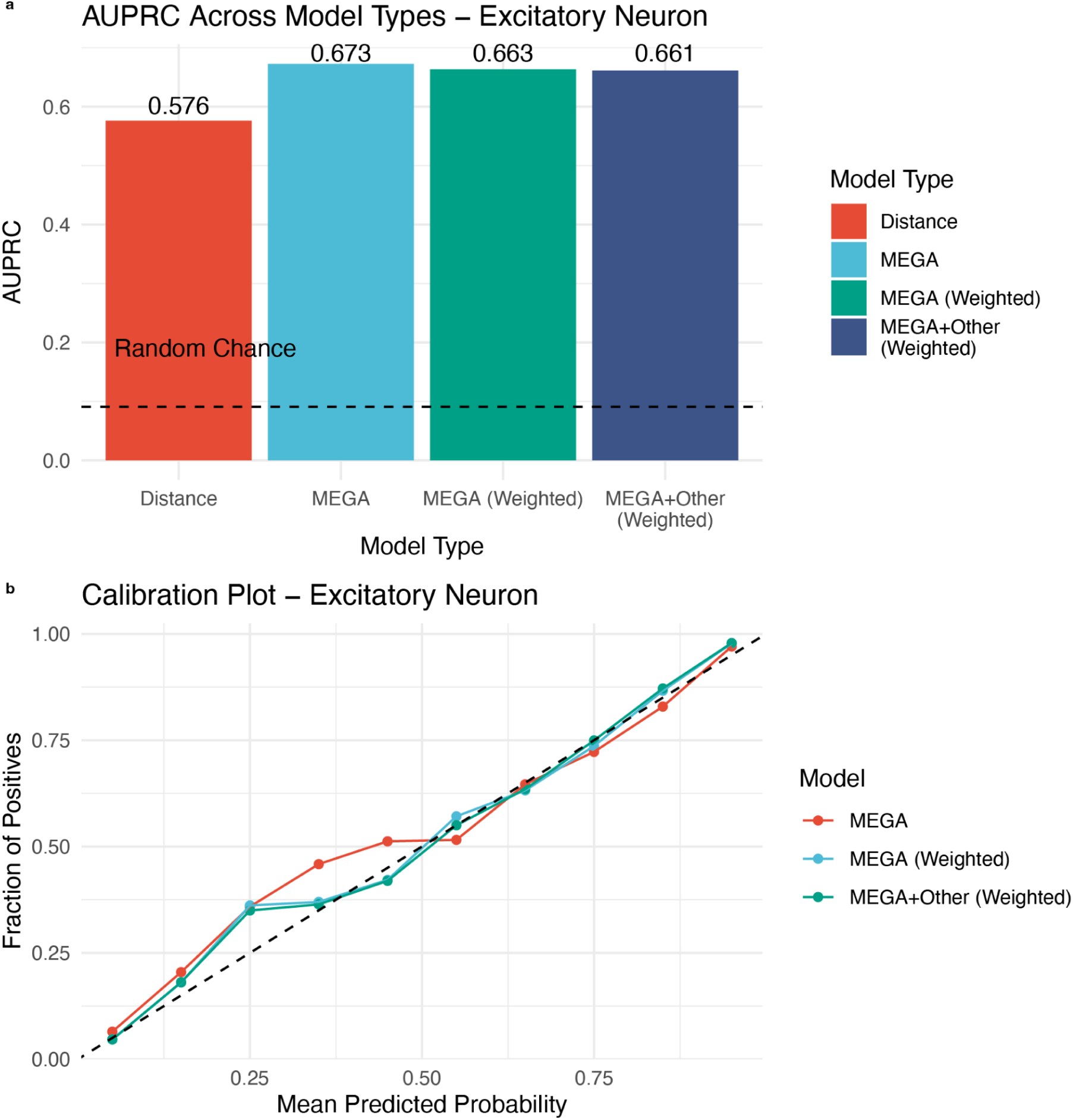
: Model performance and calibration for excitatory neurons a) AUPRC values for four model variants - distance-only baseline, MEGA DL-VEP unweighted, MEGA DL-VEP upweighted, and MEGA+Other DL-VEP upweighted. Dashed line indicates expected AUPRC under random chance (0.091). b) Calibration plots showing predicted probability versus observed positive fraction across 10 probability bins. Diagonal line indicates perfect calibration.

**Supplementary Figure 3.**
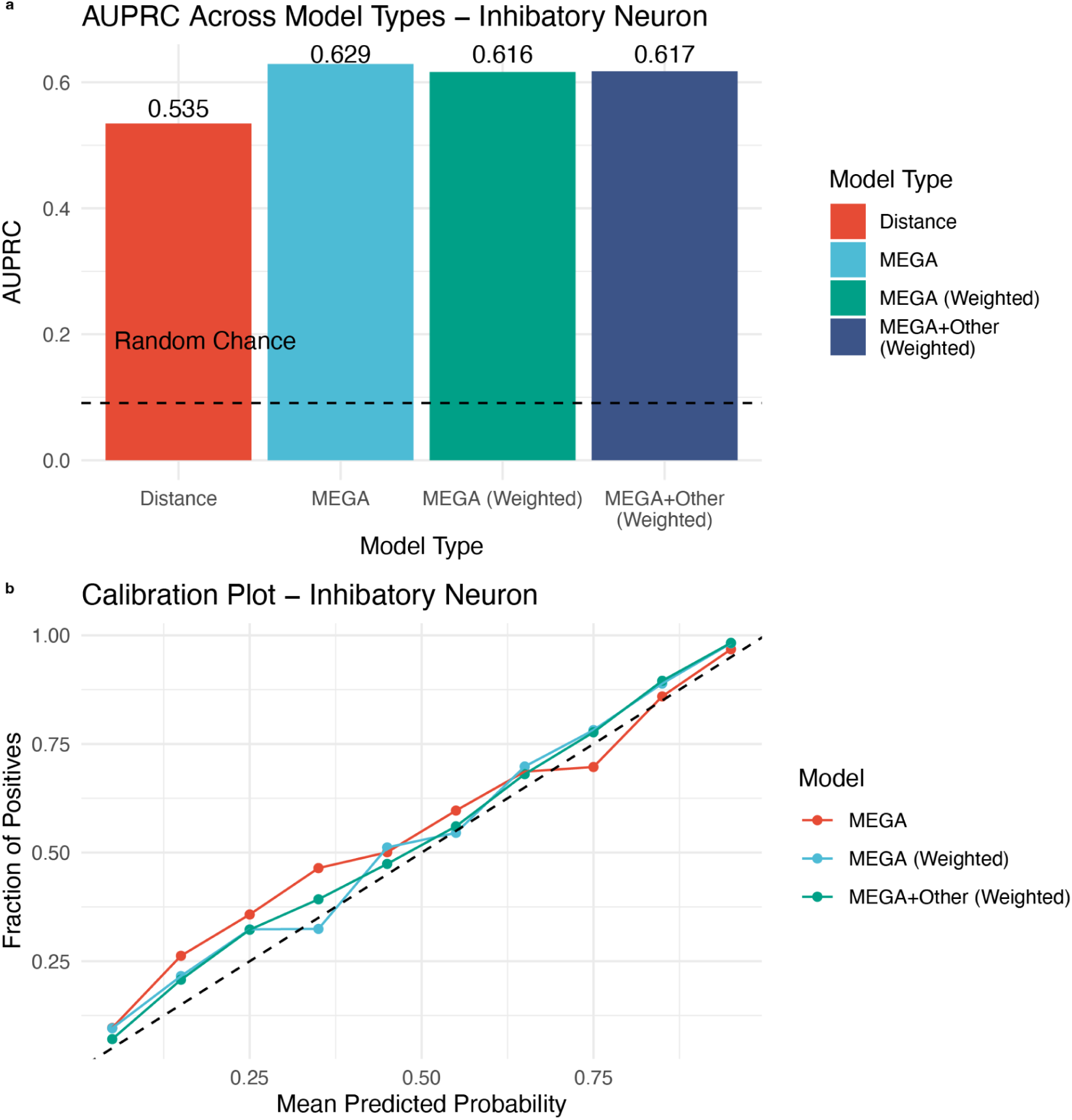
: Model performance and calibration for inhibitory neurons a) AUPRC values for four model variants - distance-only baseline, MEGA DL-VEP unweighted, MEGA DL-VEP upweighted, and MEGA+Other DL-VEP upweighted. Dashed line indicates expected AUPRC under random chance (0.091). b) Calibration plots showing predicted probability versus observed positive fraction across 10 probability bins. Diagonal line indicates perfect calibration.

**Supplementary Figure 4.**
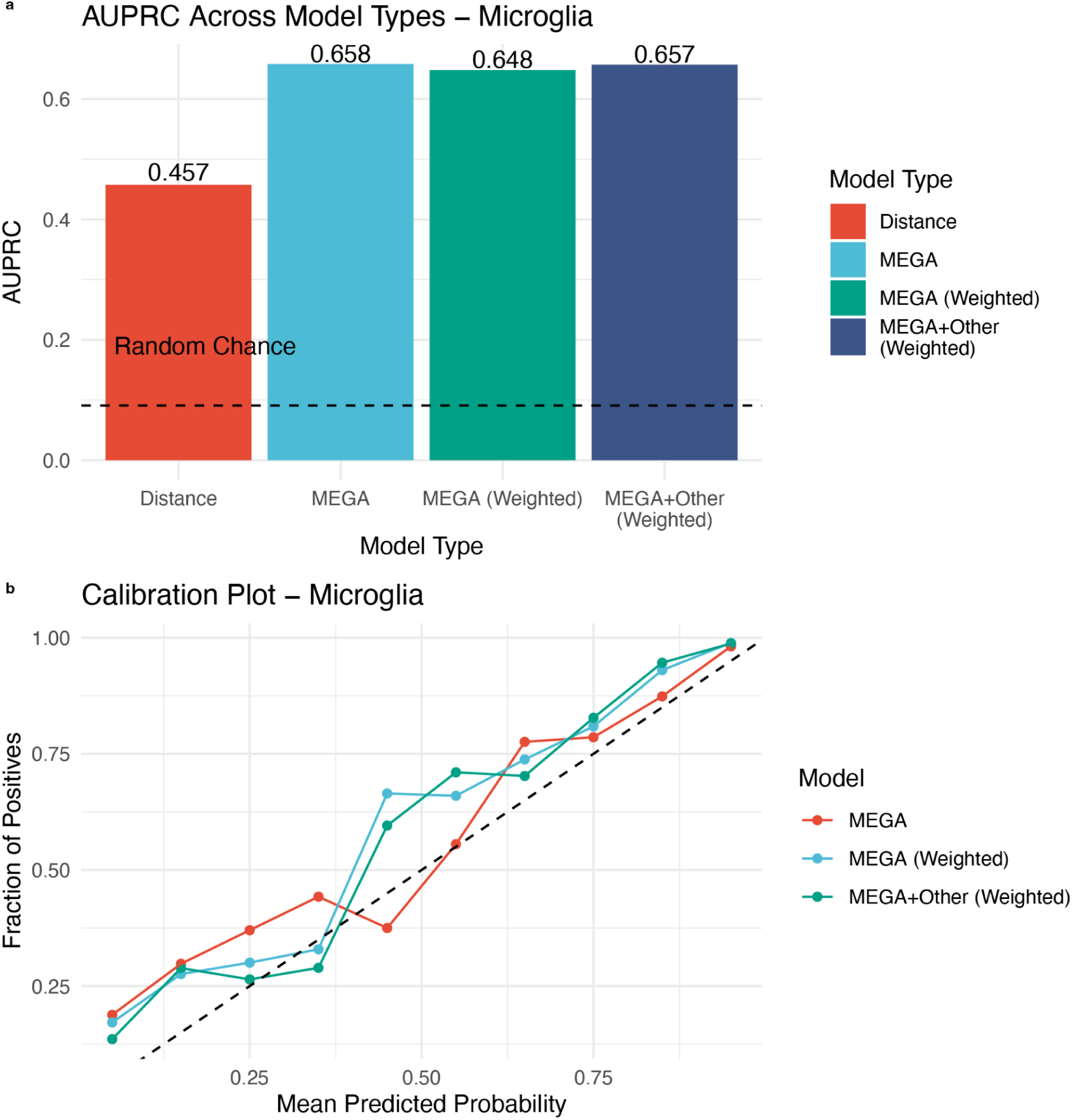
: Model performance and calibration for microglia a) AUPRC values for four model variants - distance-only baseline, MEGA DL-VEP unweighted, MEGA DL-VEP upweighted, and MEGA+Other DL-VEP upweighted. Dashed line indicates expected AUPRC under random chance (0.091). b) Calibration plots showing predicted probability versus observed positive fraction across 10 probability bins. Diagonal line indicates perfect calibration.

**Supplementary Figure 5.**
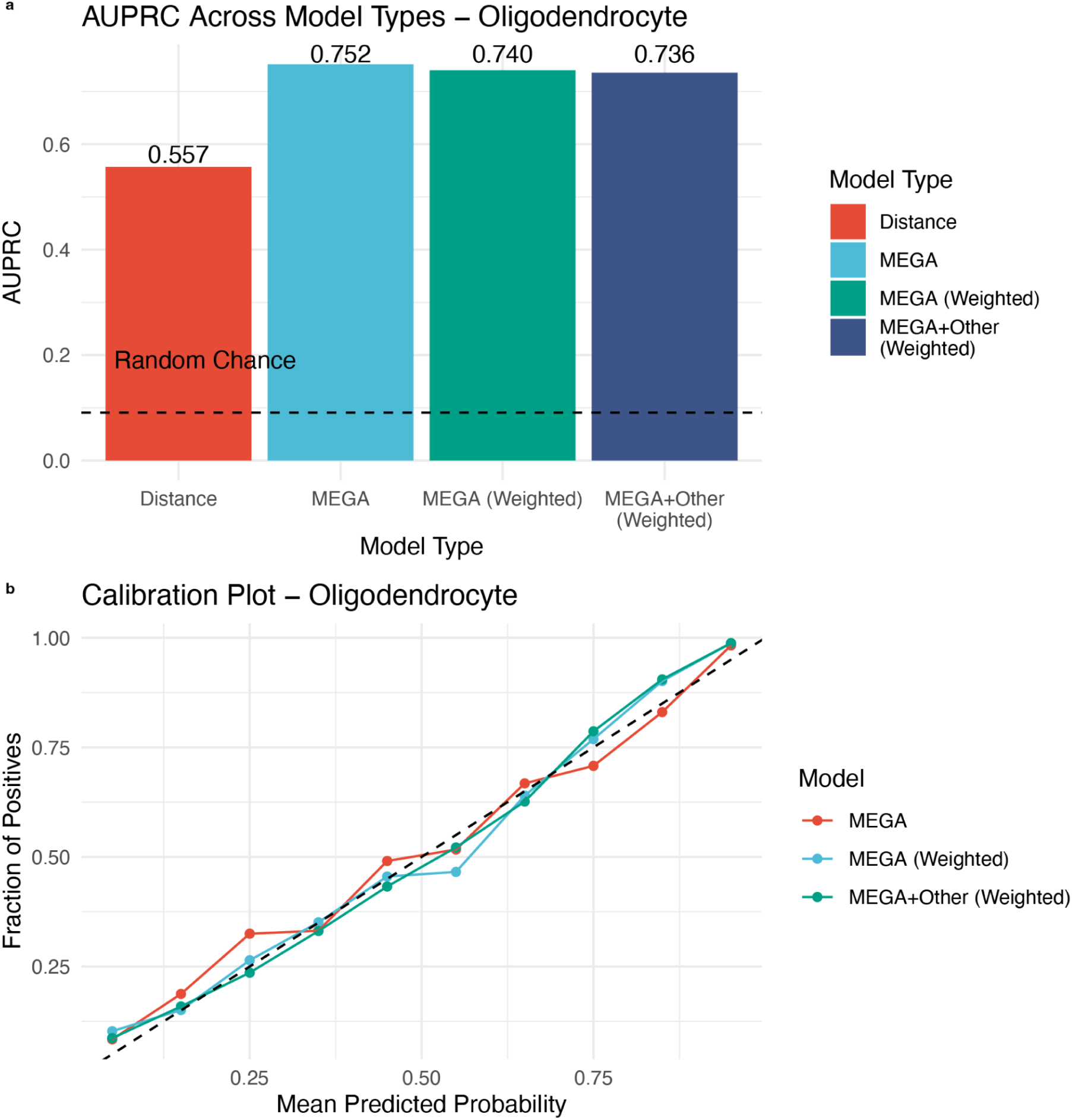
: Model performance and calibration for oligodendrocytes a) AUPRC values for four model variants - distance-only baseline, MEGA DL-VEP unweighted, MEGA DL-VEP upweighted, and MEGA+Other DL-VEP upweighted. Dashed line indicates expected AUPRC under random chance (0.091). b) Calibration plots showing predicted probability versus observed positive fraction across 10 probability bins. Diagonal line indicates perfect calibration.

**Supplementary Figure 6:**
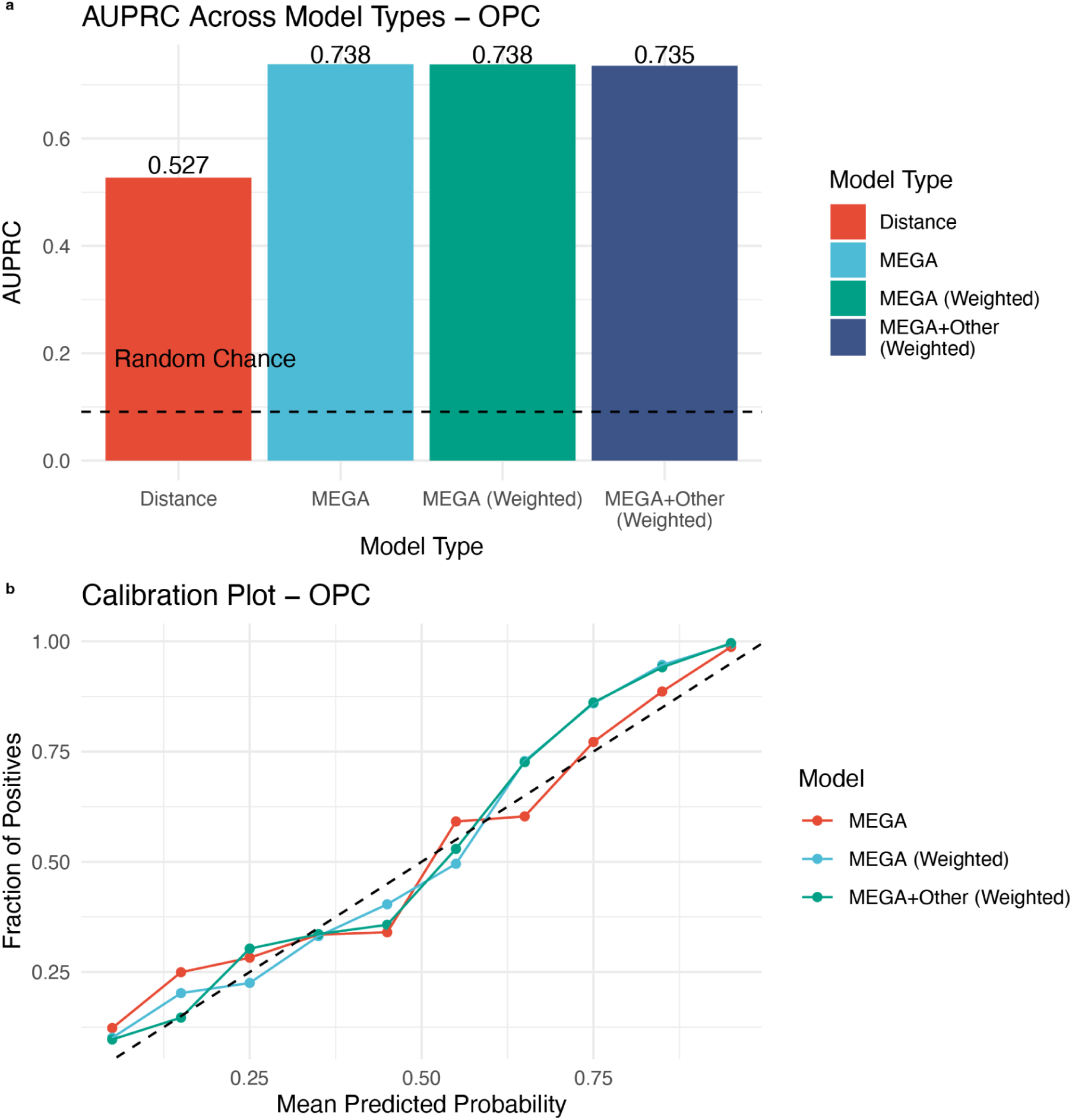
Model performance and calibration for oligodendrocyte precursor cells a) AUPRC values for four model variants - distance-only baseline, MEGA DL-VEP unweighted, MEGA DL-VEP upweighted, and MEGA+Other DL-VEP upweighted. Dashed line indicates expected AUPRC under random chance (0.091). b) Calibration plots showing predicted probability versus observed positive fraction across 10 probability bins. Diagonal line indicates perfect calibration.

**Supplementary Figure 7:**
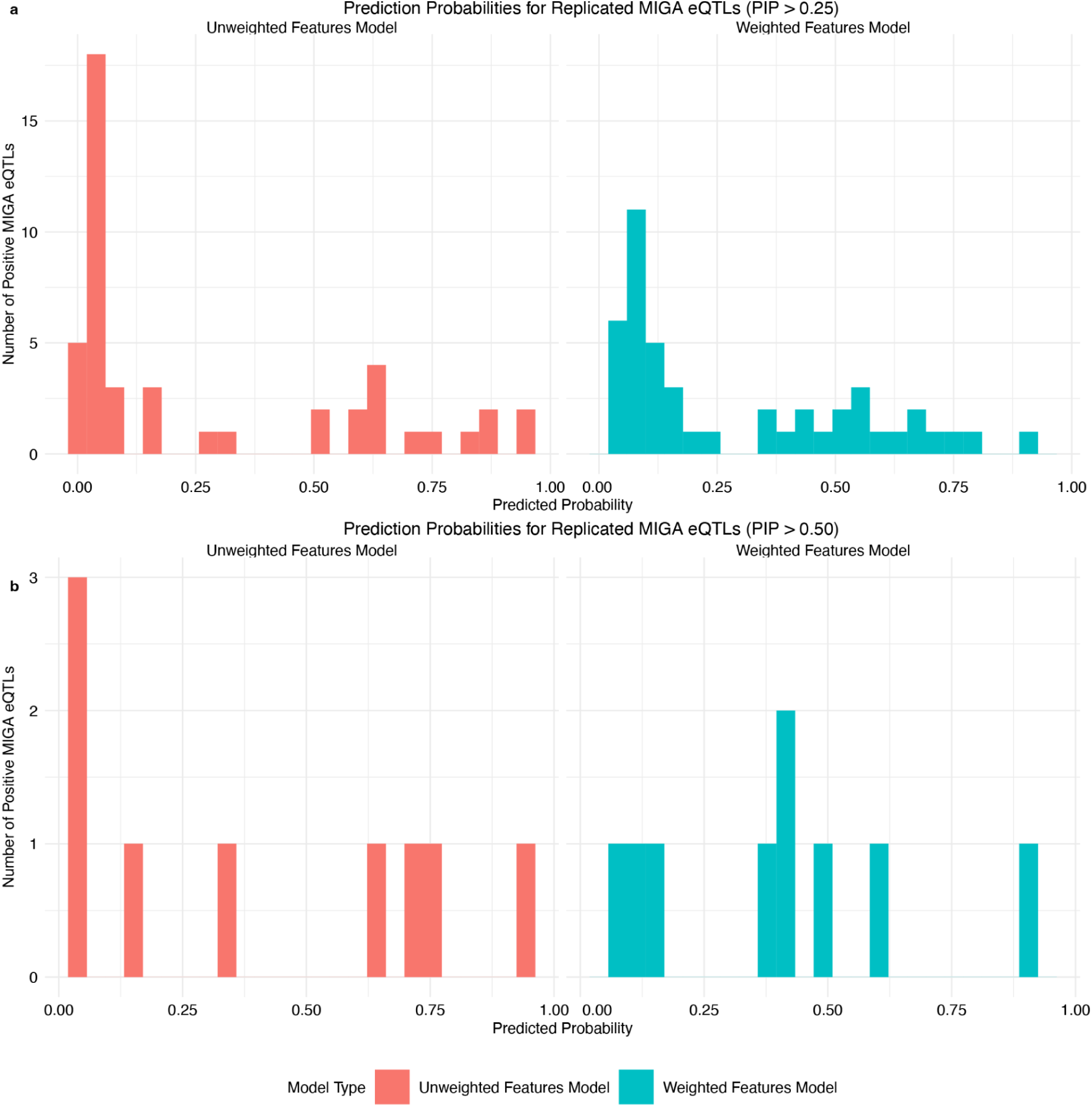
External validation using MIGA bulk microglia dataset a) Distribution of predicted probability values for fine-mapped eQTLs from the MIGA dataset (PIP > 0.25 in at least two brain regions) for both the DL-VEP unweighted (red) and DL-VEP upweighted models (blue). b) Distribution of predicted probability values for fine-mapped eQTLs from the MIGA dataset (PIP > 0.50 in at least two brain regions) for both the DL-VEP unweighted (red) and DL-VEP upweighted models (blue).

**Supplementary Figure 8:**
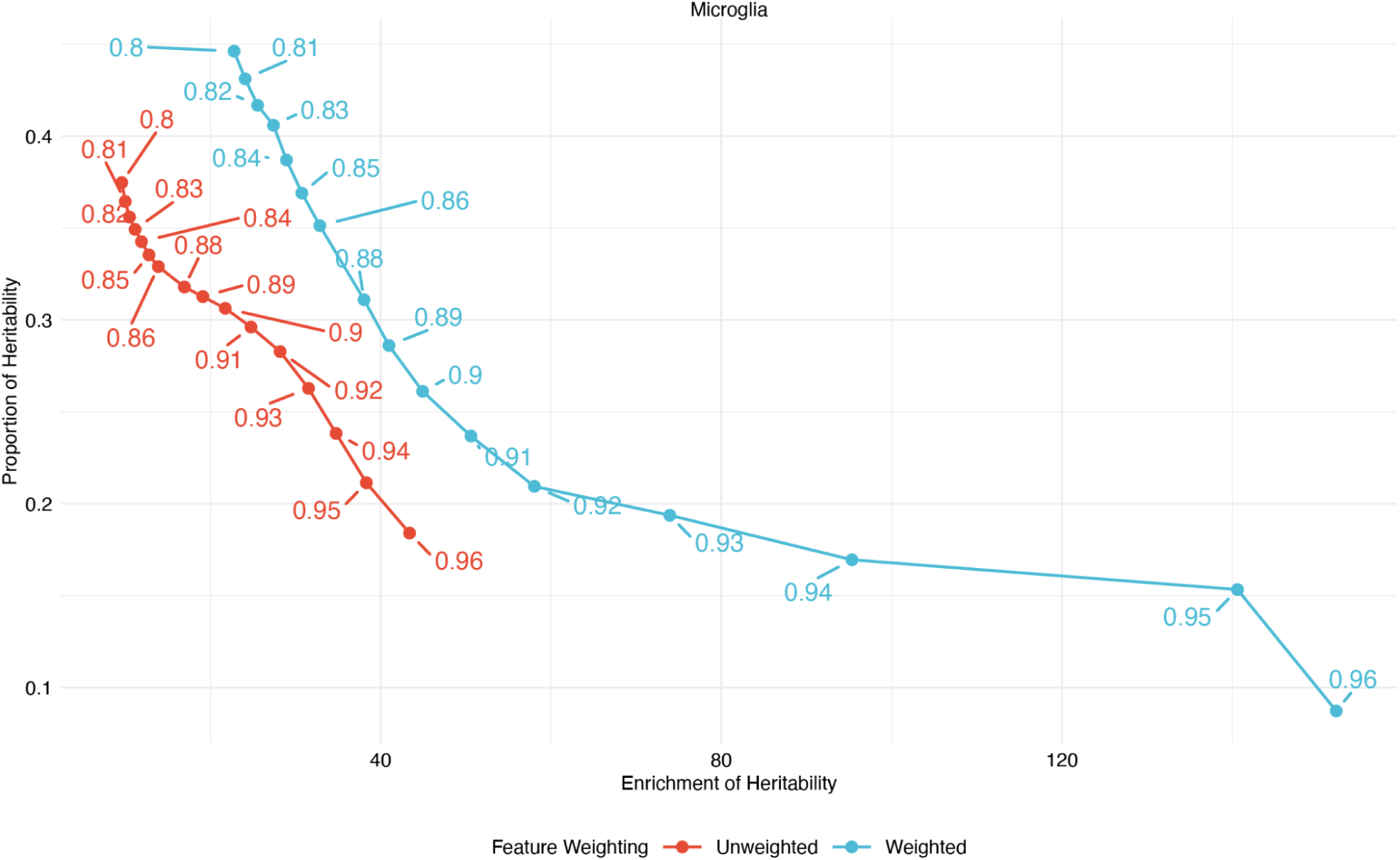
AD GWAS heritability comparisons for DL-VEP unweighted and upweighted models Proportion of heritability explained (x-axis) versus enrichment of heritability (y-axis) for binary annotations created using model predictions of DL-VEP unweighted and DL-unweighted models. Binary annotations consist of variants with predicted probability above a certain threshold (denoted by numerical value) for each model.

**Supplementary Figure 9:**
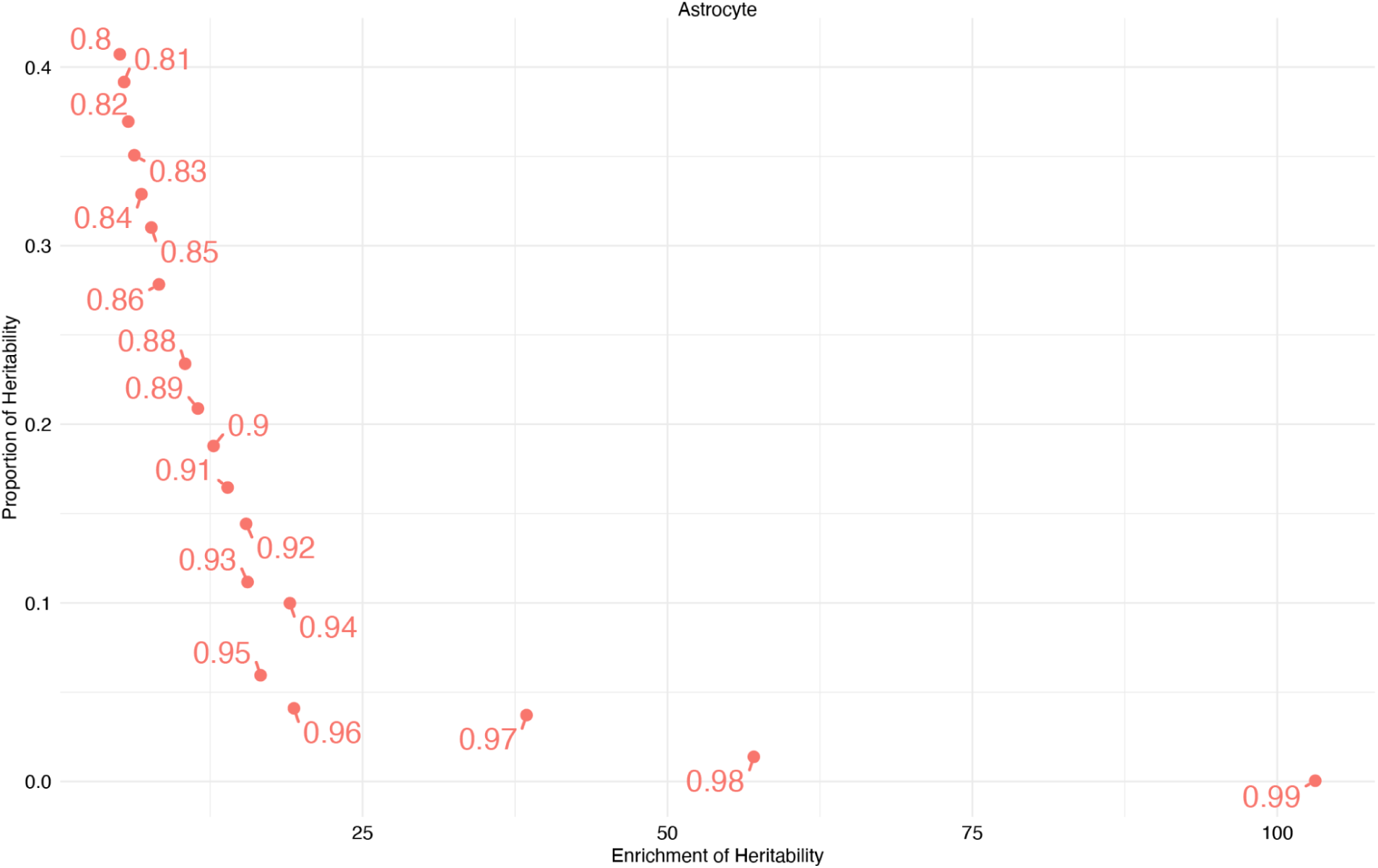
AD GWAS heritability vs enrichment of heritability for predicted eQTLs (Astrocyte) Proportion of heritability explained (x-axis) versus enrichment of heritability (y-axis) for binary annotations created using model predictions from astroctye scEEMS model. Binary annotations consist of variants with predicted probability above a certain threshold (denoted by numerical value) for each model. (Not shown: prediction probabilities with negative enrichment of heritability)

**Supplementary Figure 10:**
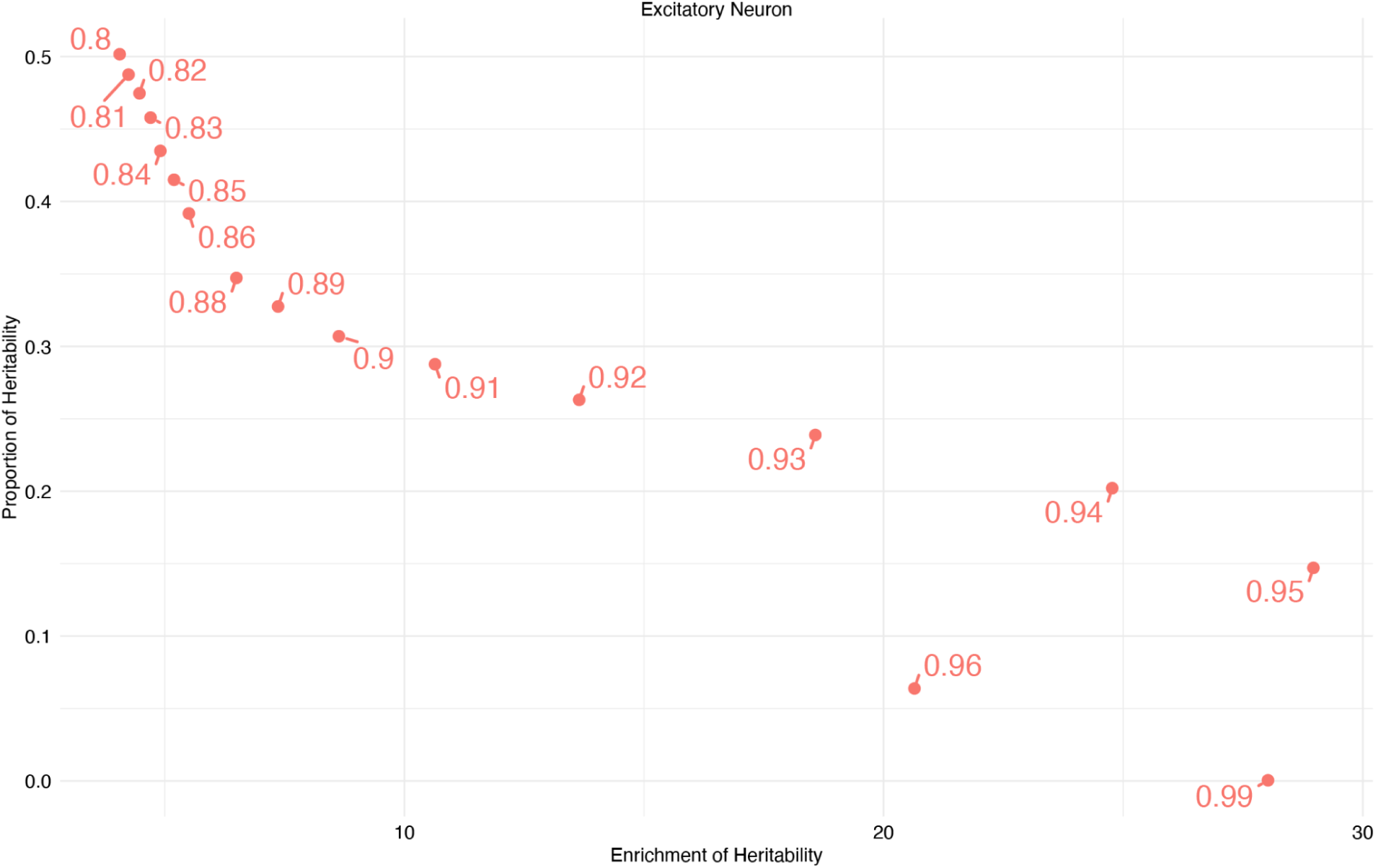
AD GWAS heritability of predicted eQTLs (Excitatory Neuron) Proportion of heritability explained (x-axis) versus enrichment of heritability (y-axis) for binary annotations created using model predictions from excitatory neuron scEEMS model. Binary annotations consist of variants with predicted probability above a certain threshold (denoted by numerical value) for each model. (Not shown: prediction probabilities with negative enrichment of heritability)

**Supplementary Figure 11:**
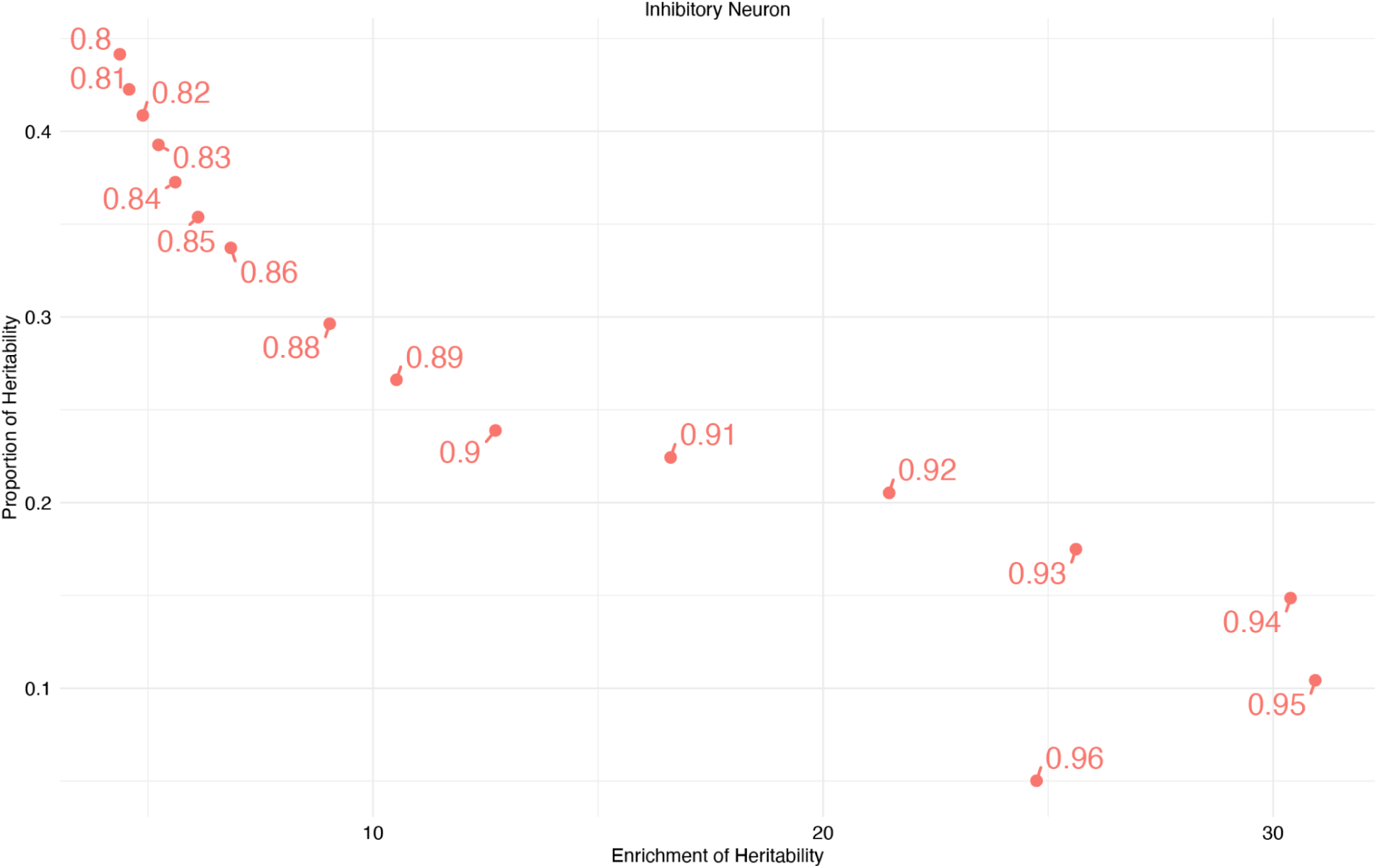
AD GWAS heritability of predicted eQTLs (Inhibitory Neuron) Proportion of heritability explained (x-axis) versus enrichment of heritability (y-axis) for binary annotations created using model predictions from inhibitory neuron scEEMS model. Binary annotations consist of variants with predicted probability above a certain threshold (denoted by numerical value) for each model. (Not shown: prediction probabilities with negative enrichment of heritability)

**Supplementary Figure 12:**
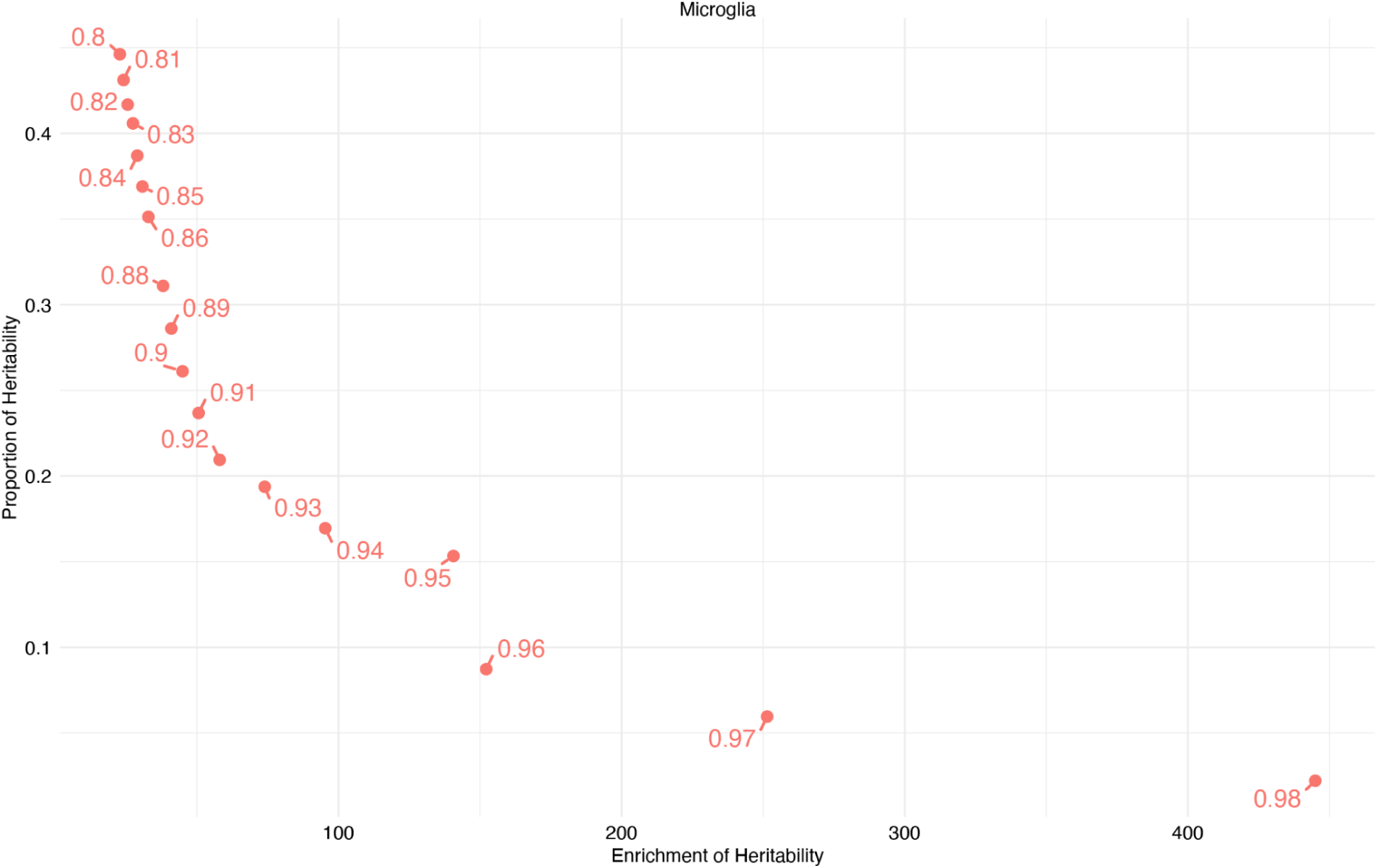
AD GWAS heritability of predicted eQTLs (Microglia) Proportion of heritability explained (x-axis) versus enrichment of heritability (y-axis) for binary annotations created using model predictions from microglia scEEMS model. Binary annotations consist of variants with predicted probability above a certain threshold (denoted by numerical value) for each model. (Not shown: prediction probabilities with negative enrichment of heritability)

**Supplementary Figure 13:**
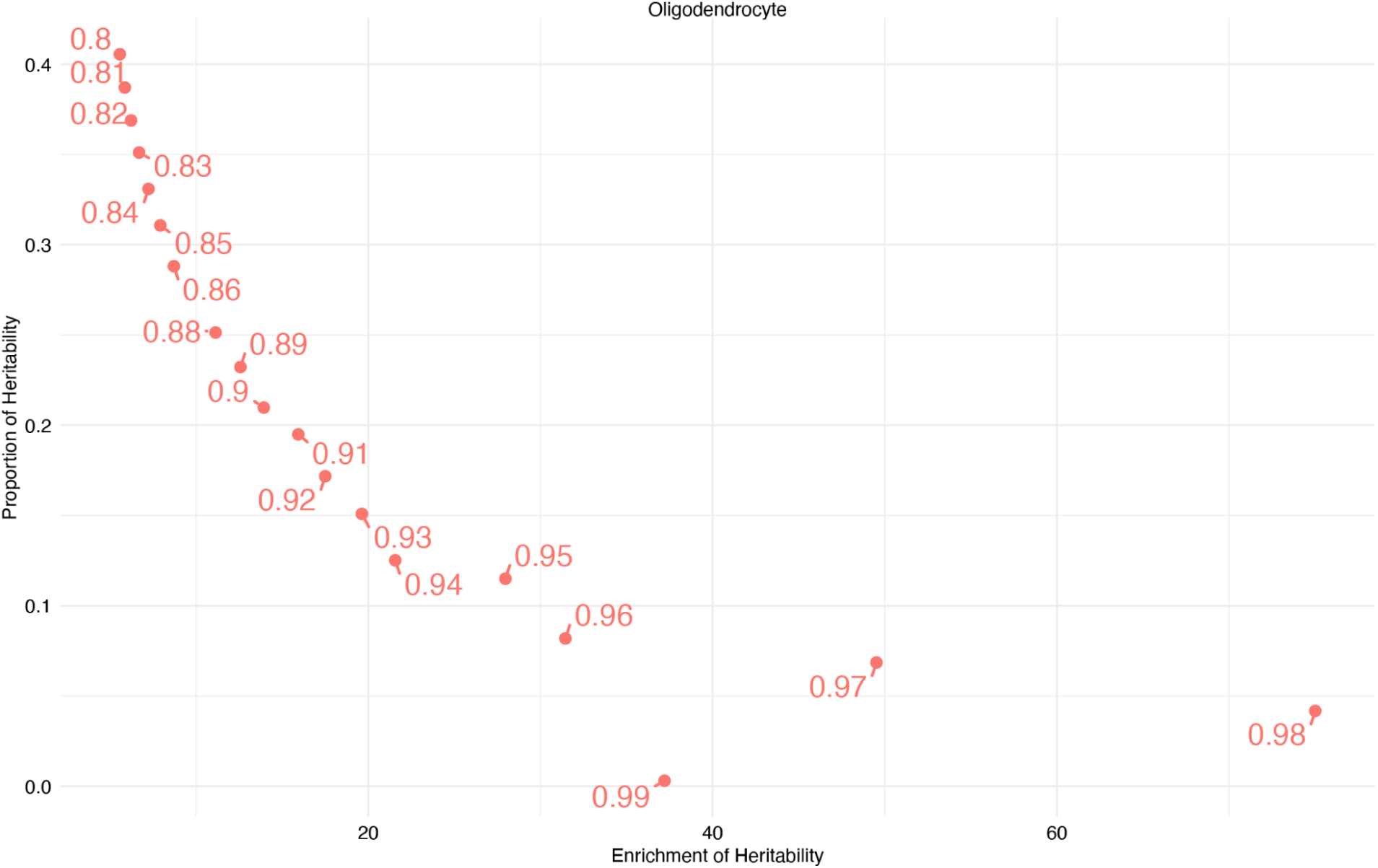
A**D GWAS heritability of predicted eQTLs (Oligodendrocyte)** Proportion of heritability explained (x-axis) versus enrichment of heritability (y-axis) for binary annotations created using model predictions from oligodendrocyte scEEMS model. Binary annotations consist of variants with predicted probability above a certain threshold (denoted by numerical value) for each model. (Not shown: prediction probabilities with negative enrichment of heritability)

**Supplementary Figure 14:**
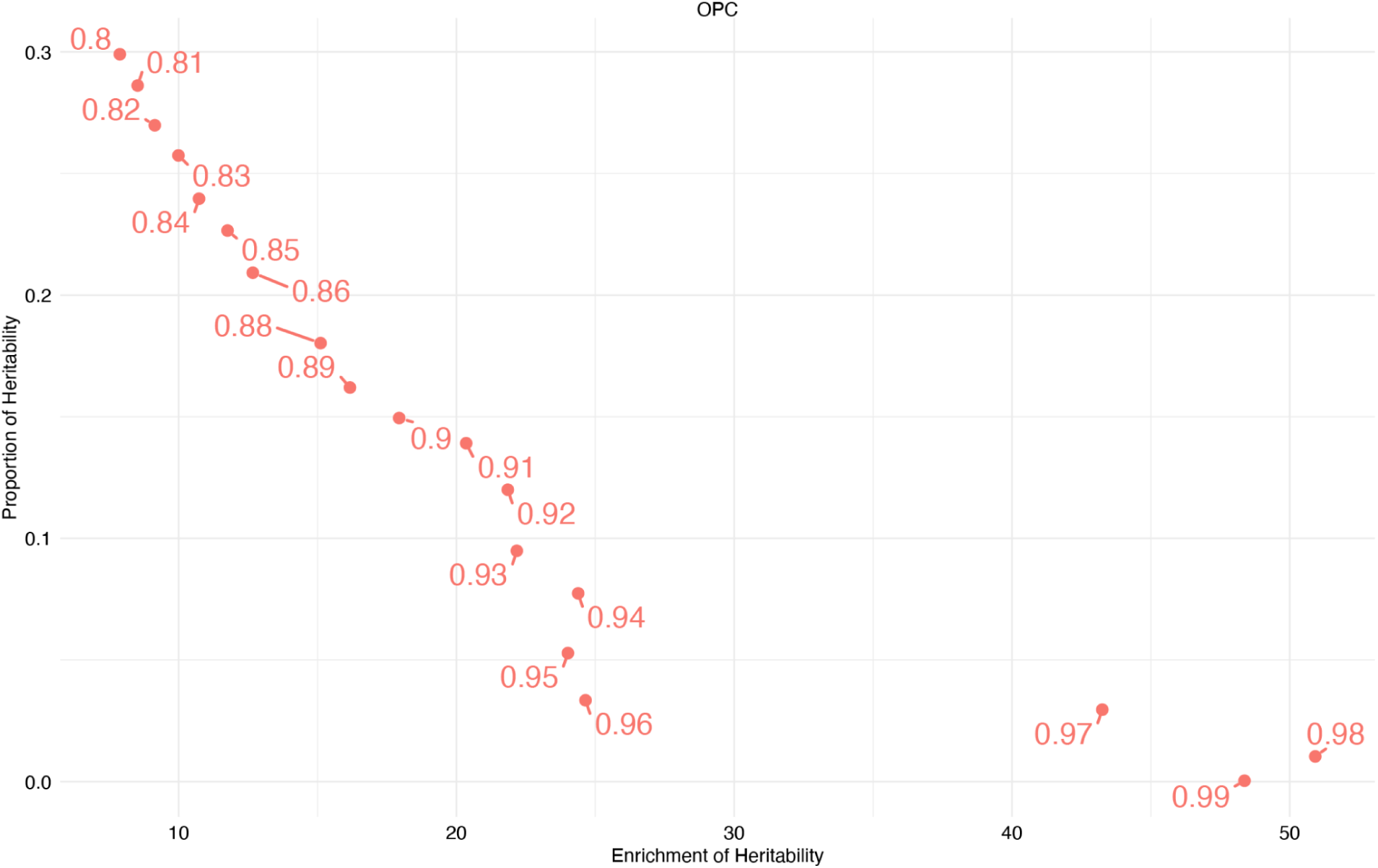
AD GWAS heritability of predicted eQTLs (OPC) Proportion of heritability explained (x-axis) versus enrichment of heritability (y-axis) for binary annotations created using model predictions from OPC scEEMS model. Binary annotations consist of variants with predicted probability above a certain threshold (denoted by numerical value) for each model. (Not shown: prediction probabilities with negative enrichment of heritability)

**Supplementary Figure 15:**
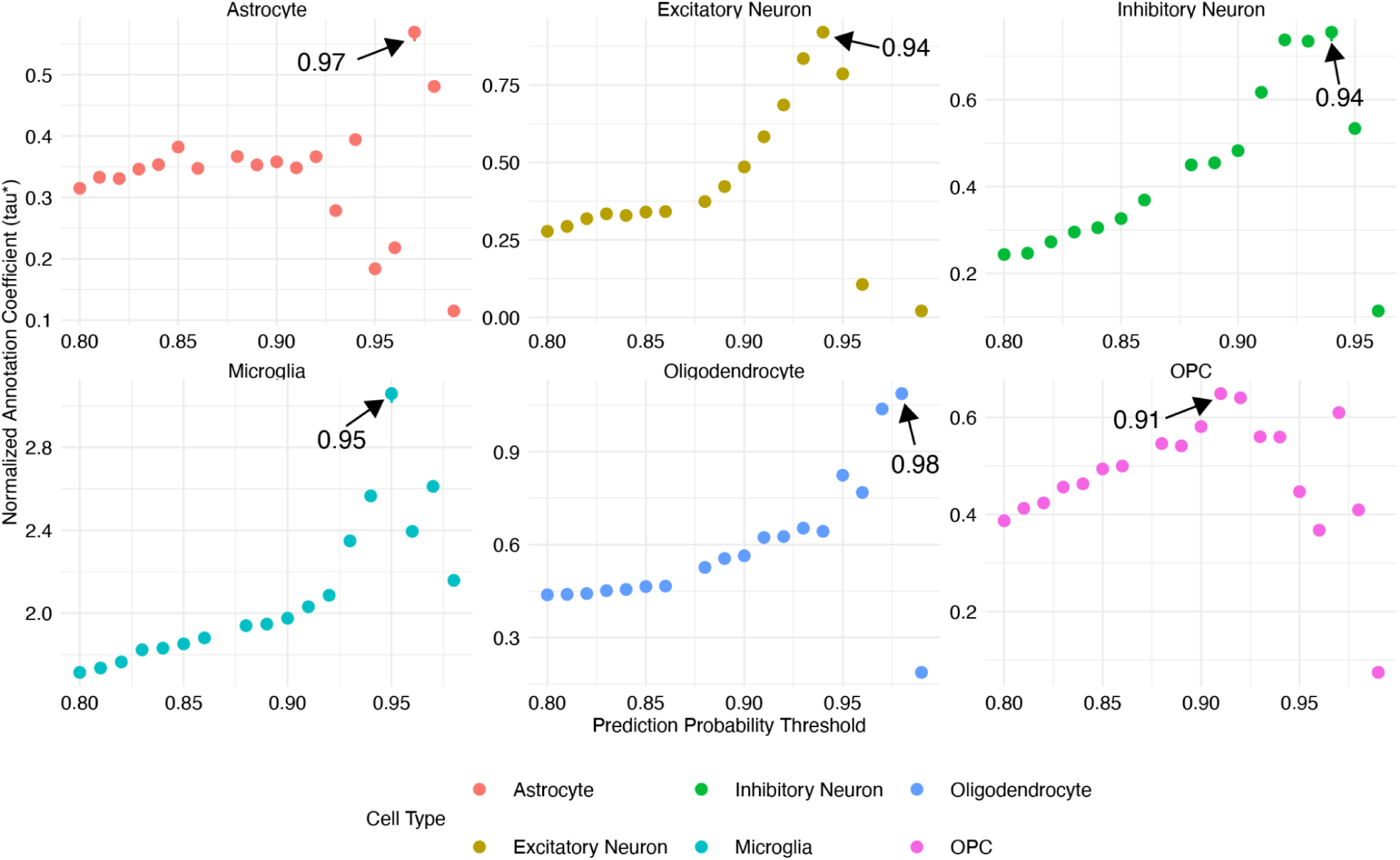
Cell type-specific tau* values (y-axis) across prediction probability thresholds (x-axis). The labeled point denotes the prediction probability threshold with the highest tau* value for each cell type.

**Supplementary Figure 16:**
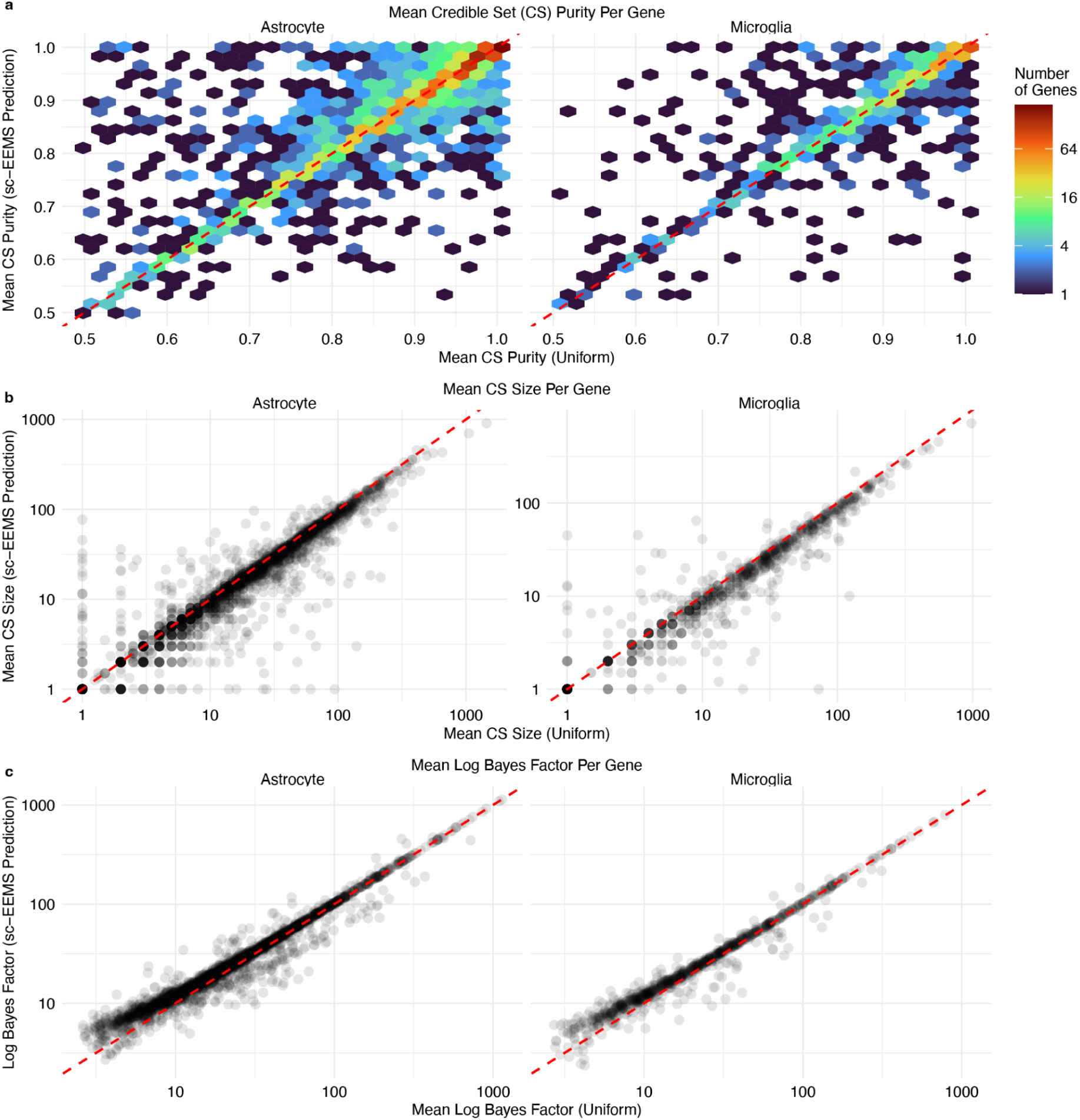
a) Mean purity across credible sets for each gene comparing the scEEMS prediction prior versus uniform prior. b) Mean credible set size for each gene comparing the scEEMS prediction prior versus uniform prior. c) Mean log Bayes factor (lbf) for each gene comparing the scEEMS prediction prior versus uniform prior.

**Supplementary Figure 17:**
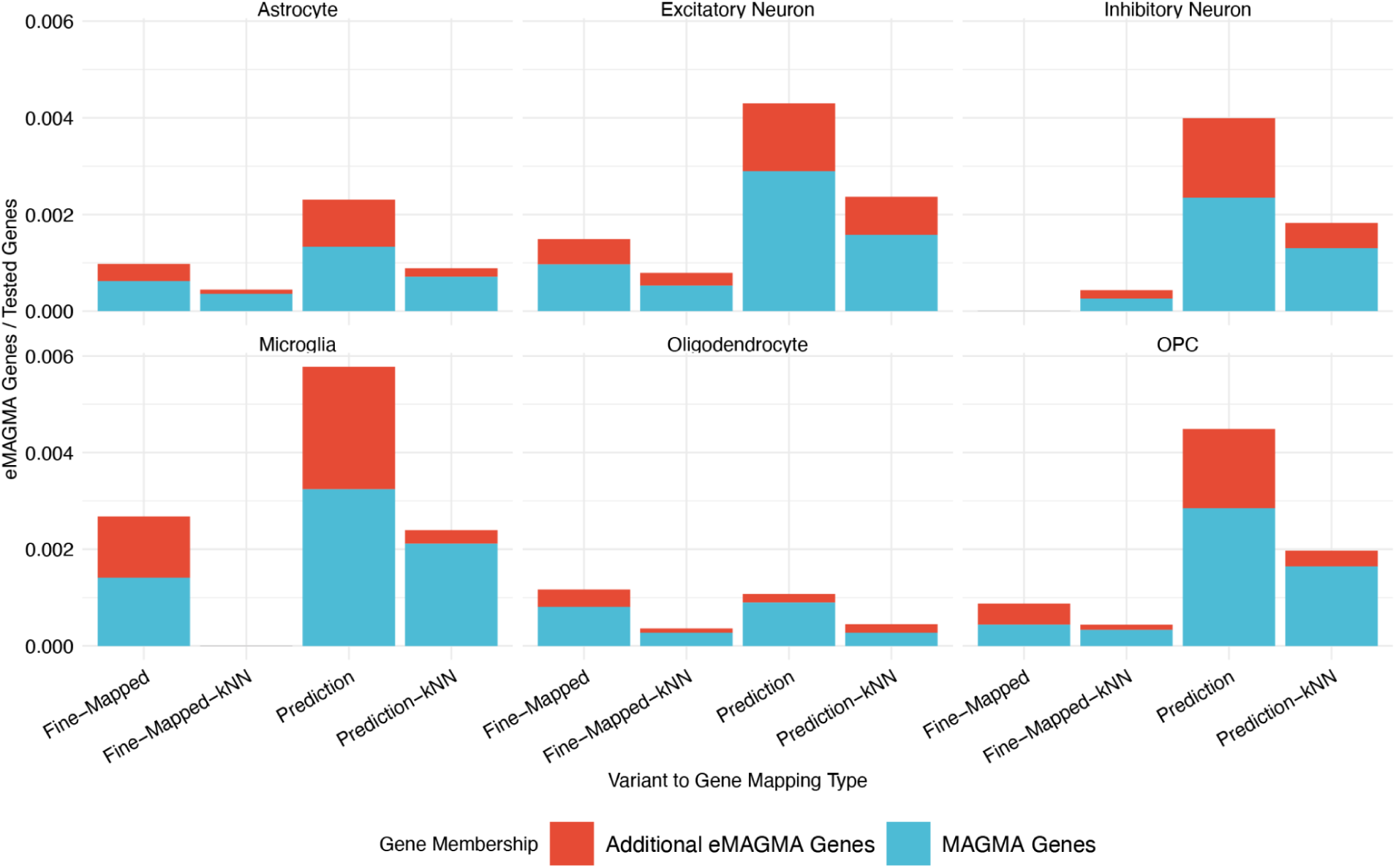
Percentage of significant eMAGMA genes (Bonferroni-corrected P < 2.67×10⁻⁶) using fine-mapped and predicted eQTLs for each cell type as well as their respective gene-matched kNN baseline. Blue bars: genes also found by standard MAGMA; red bars: eMAGMA-specific genes.

**Supplementary Figure 18:**
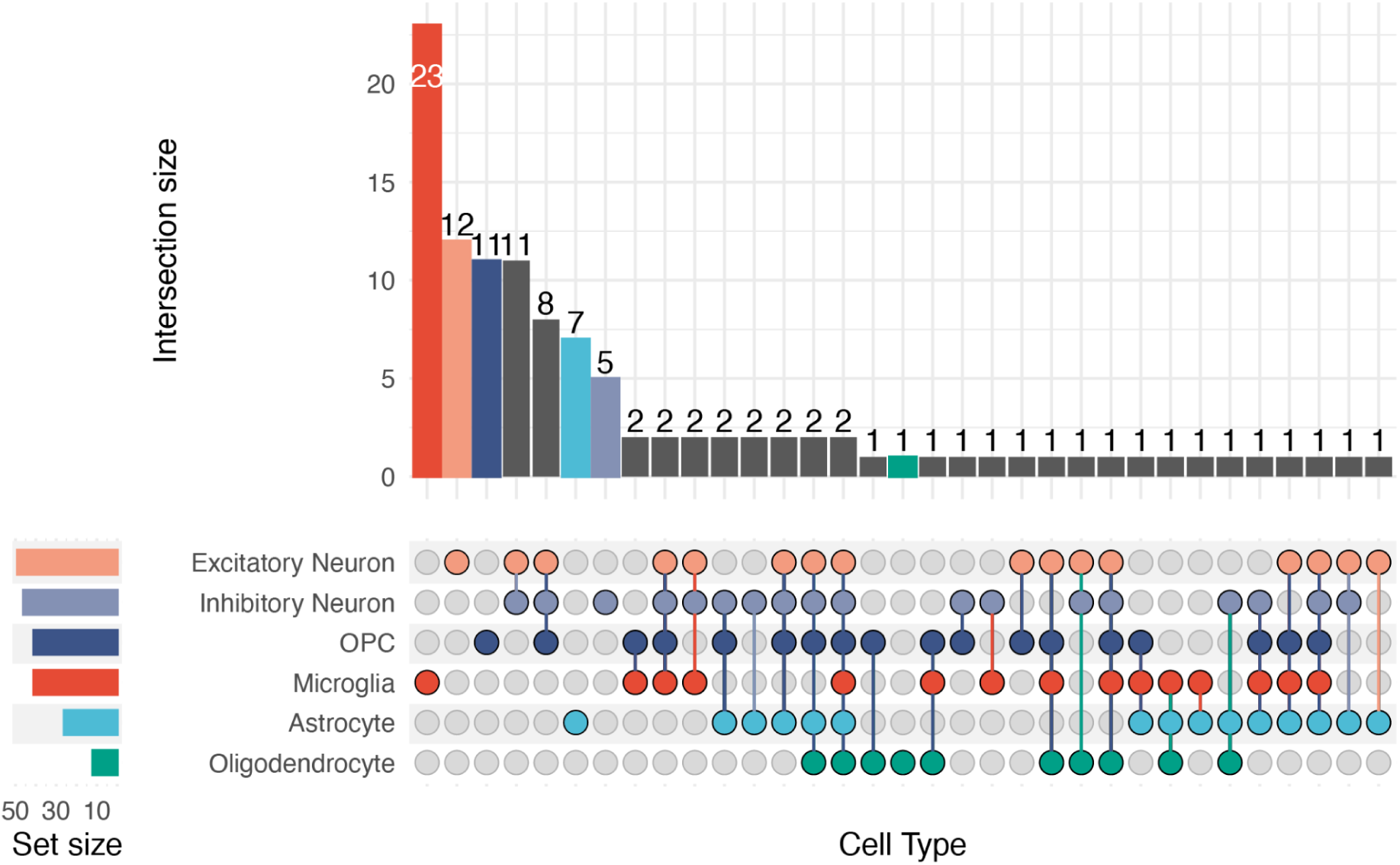
UpSet plot showing cell-type membership of all significant scEEMS predicted eMAGMA genes, including all sets (untruncated version of Figure 5b).

**Supplementary Figure 19:**
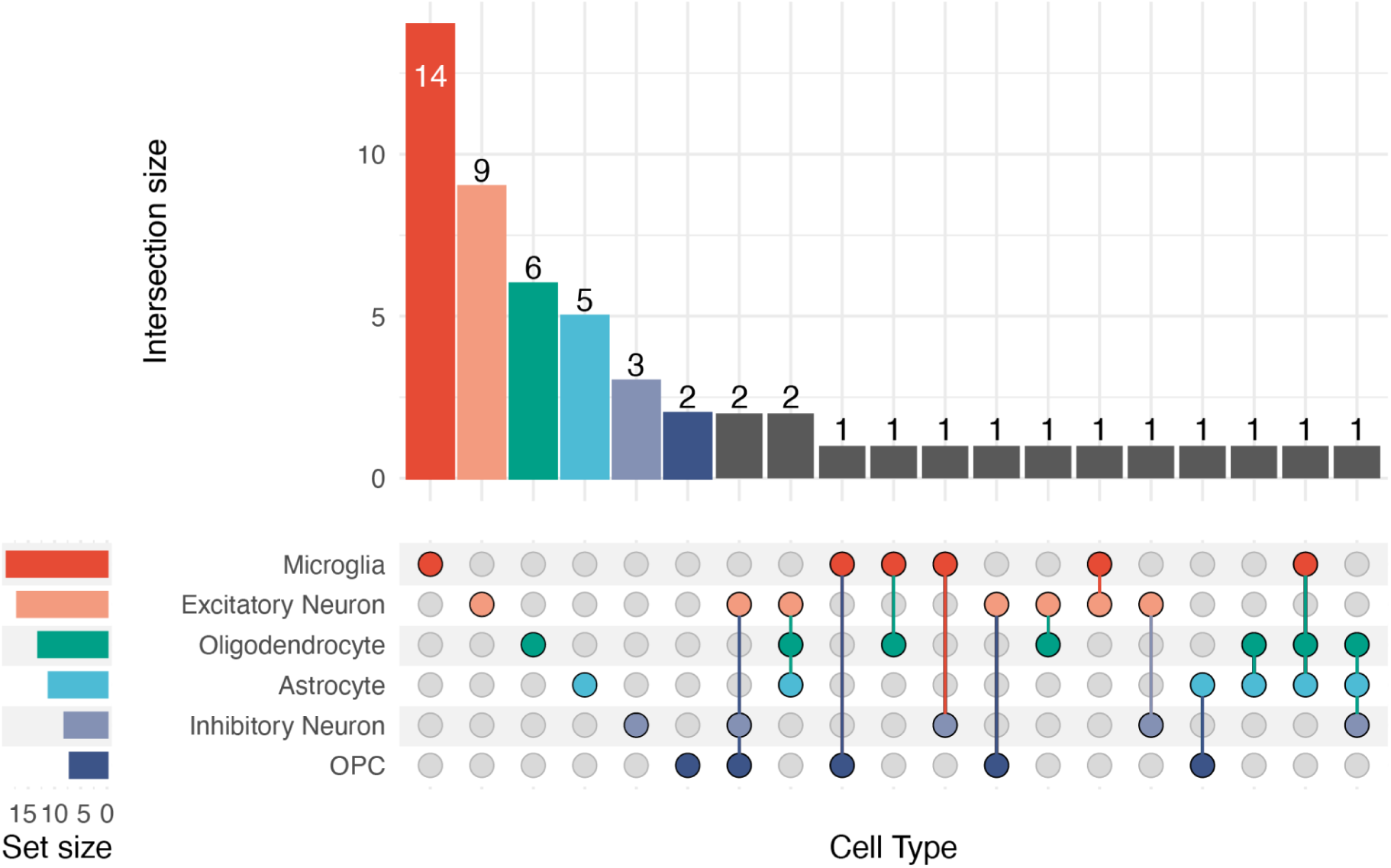
UpSet plot showing cell-type membership of all significant fine-mapped eMAGMA genes.

**Supplementary Figure 20:**
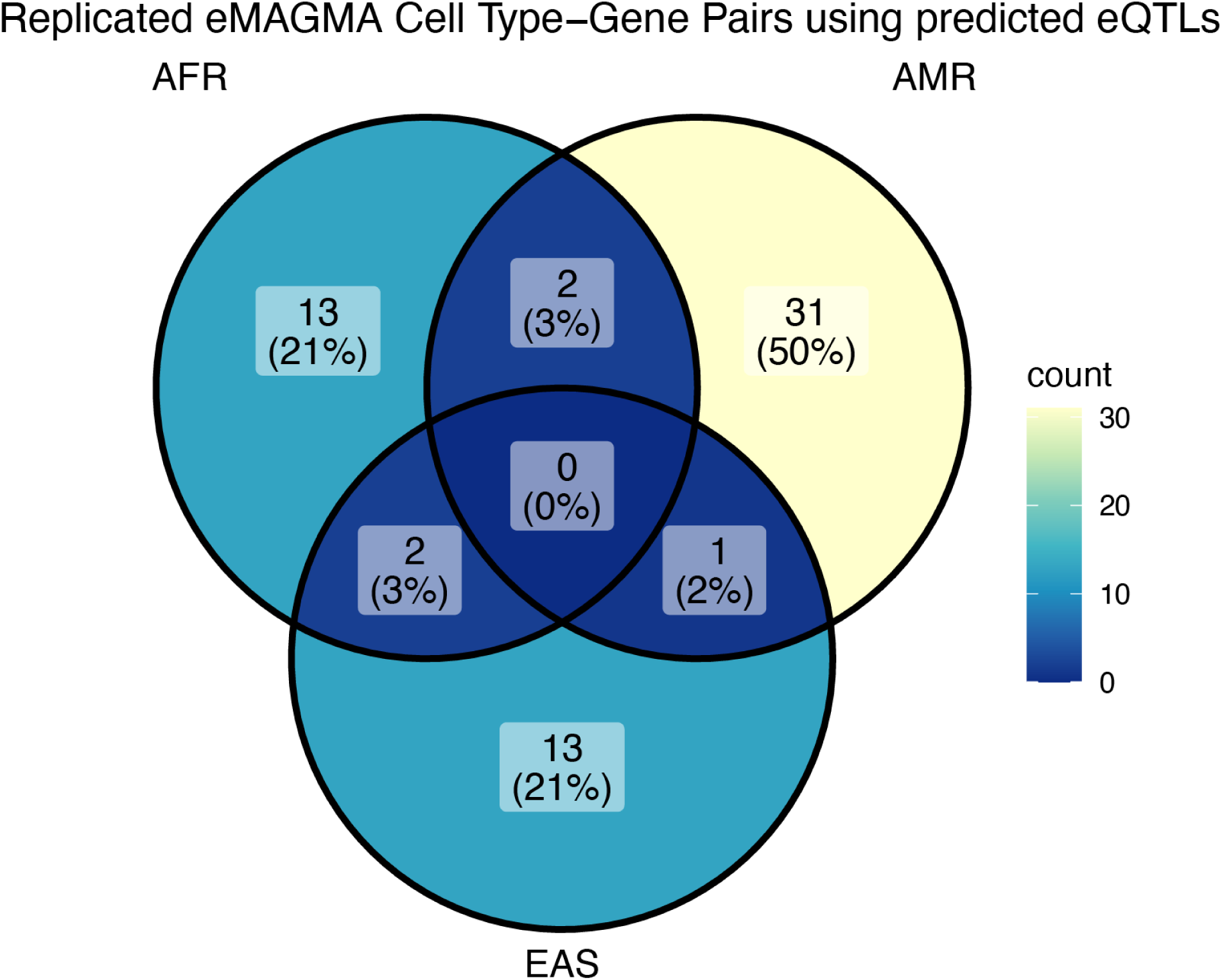
Venn diagram of the number of replicated scEEMS predicted eQTL eMAGMA genes shared between EUR and various non-EUR populations.

**Supplementary Figure 21:**
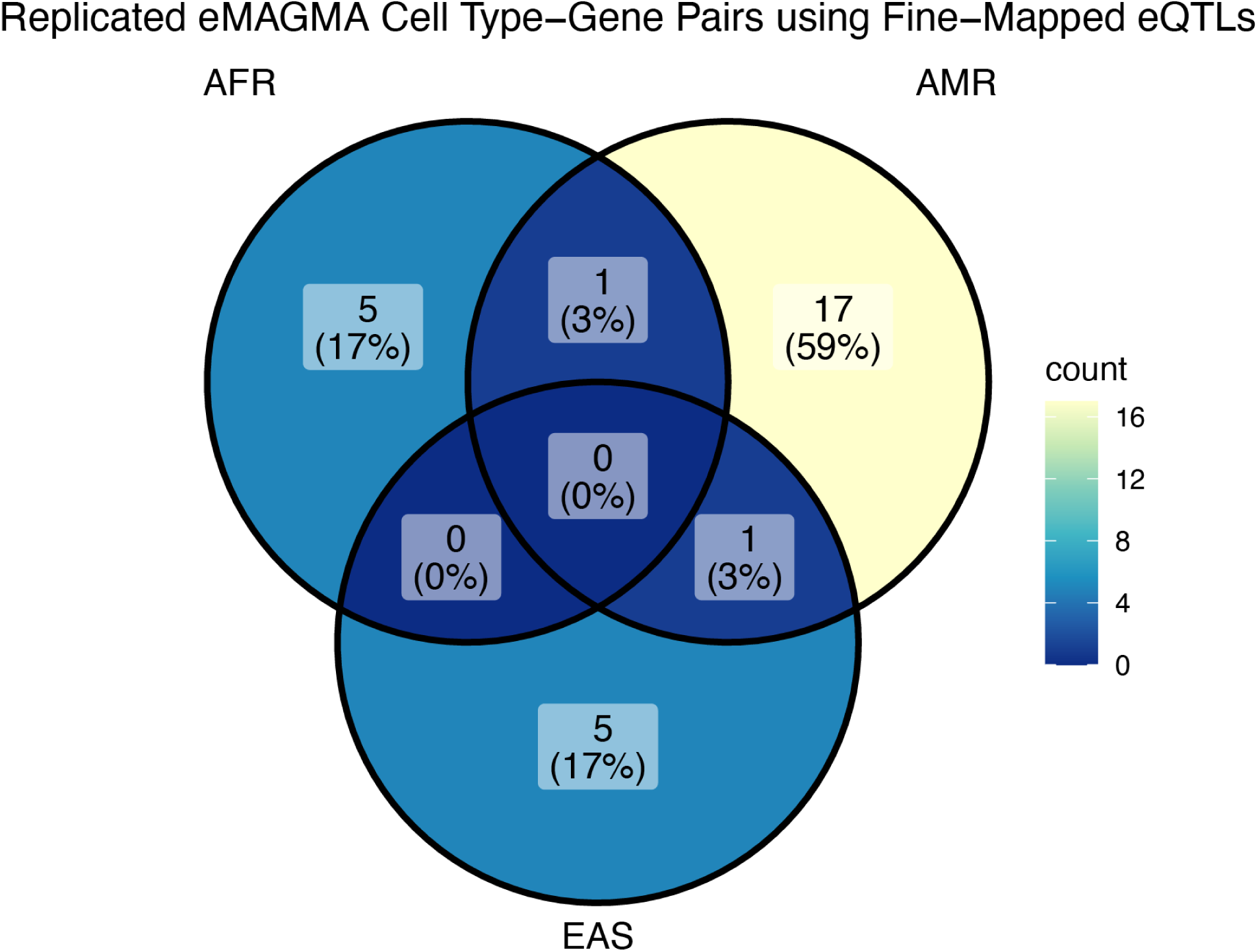
Venn diagram of the number of replicated fine-mapped eQTL eMAGMA genes shared between EUR and various non-EUR populations.

**Supplementary Figure 22:**
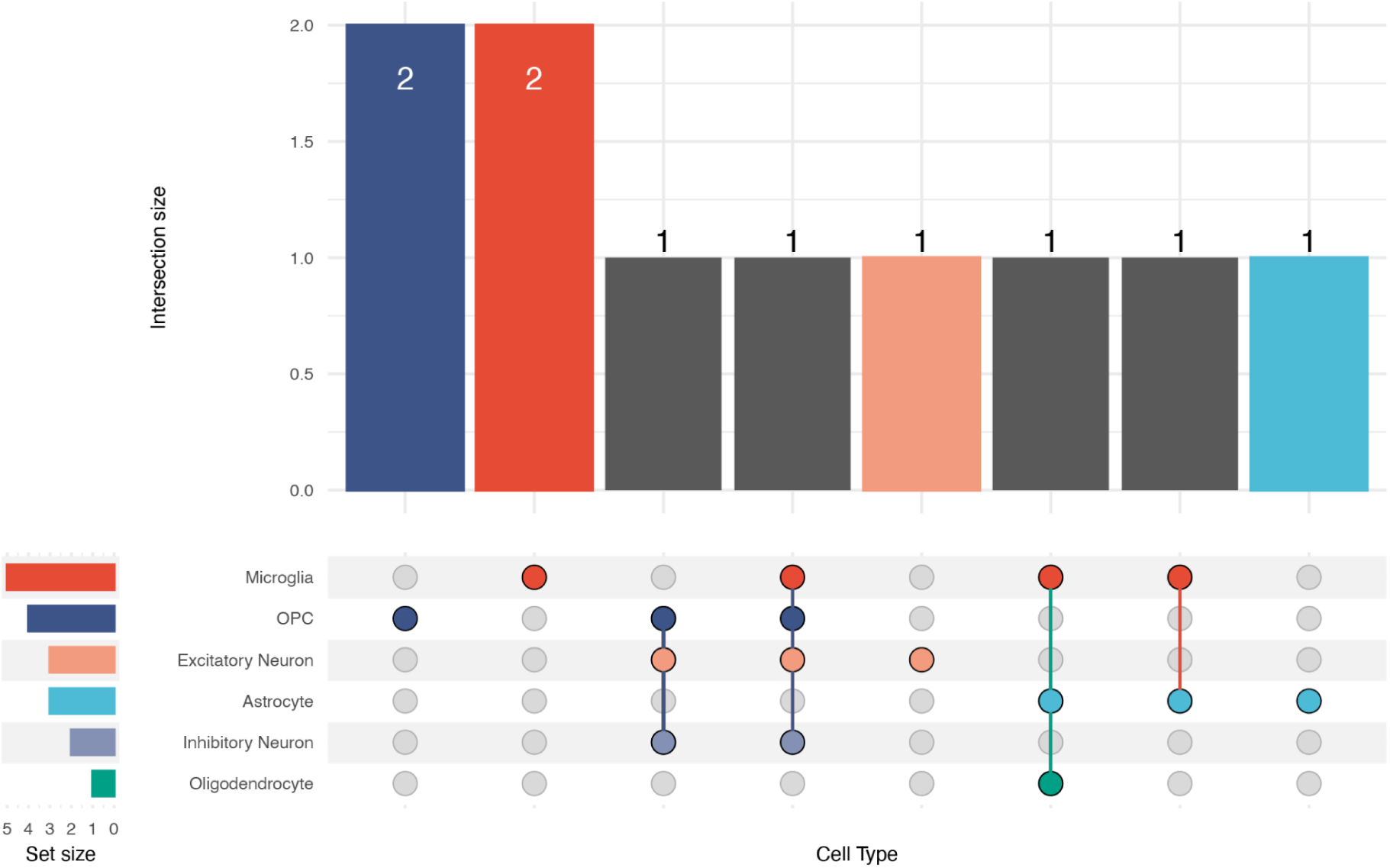
UpSet plot showing cell-type membership of all significant fine-mapped eMAGMA genes after filtering out MAGMA and fine-mapped eMAGMA genes.

**Supplementary Figure 23:**
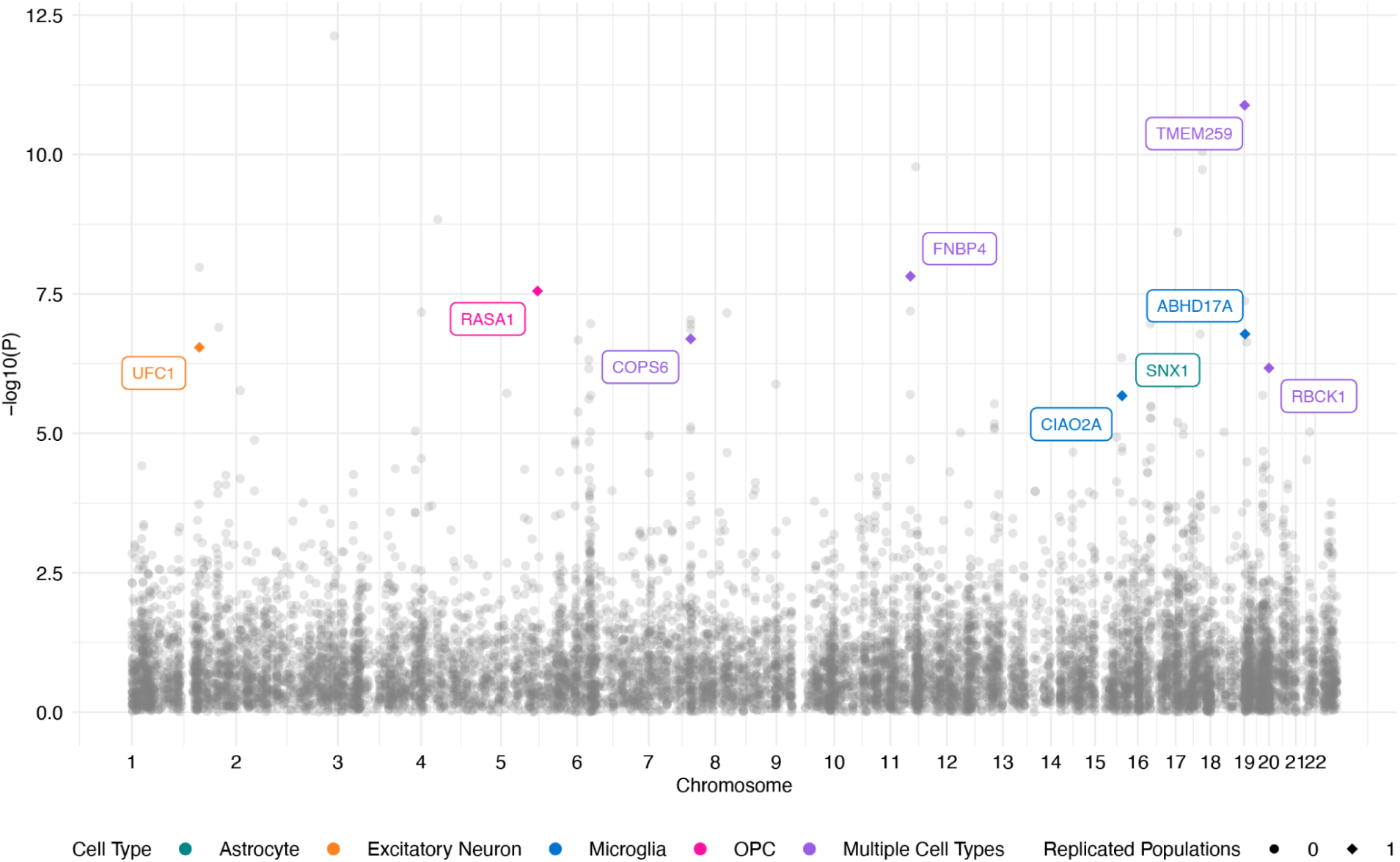
Manhattan plot of significant scEEMS predicted eMAGMA genes after filtering out fine-mapped eMAGMA and MAGMA genes in the European AD GWAS. Gray: no replication in non-European populations; colored: replicated (P < 0.05) in non-European populations. Circles: zero replicated populations; diamonds: one replicated population; squares: two replicated populations. Color indicates cell type.

**Supplementary Figure 24:**
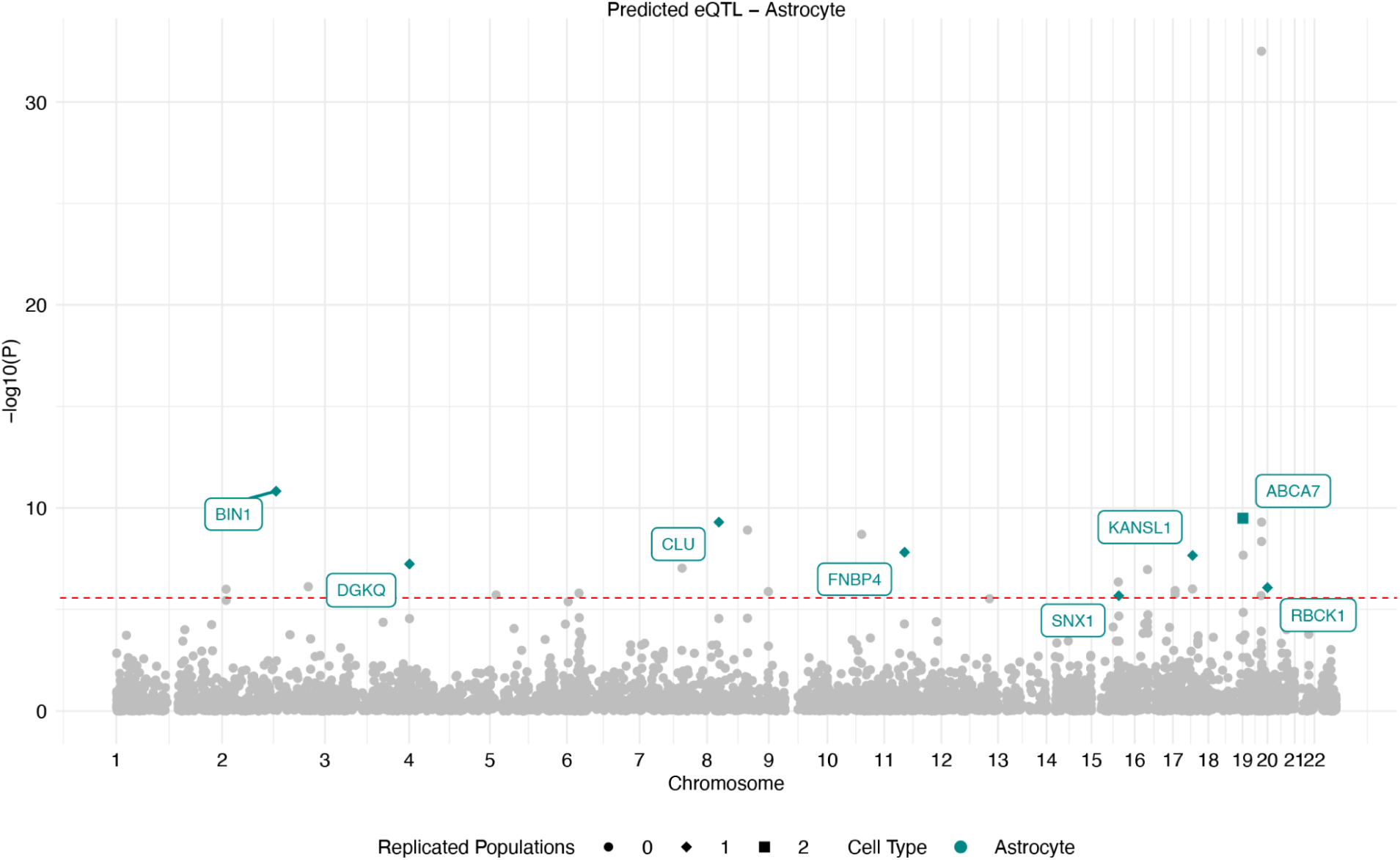
Manhattan plot of astrocyte-specific scEEMS predicted eMAGMA genes after filtering out fine-mapped eMAGMA and MAGMA genes in the European AD GWAS. Gray: no replication in non-European populations; colored: replicated (P < 0.05) in non-European populations. Circles: zero replicated populations; diamonds: one replicated population; squares: two replicated populations. Color indicates cell type.

**Supplementary Figure 25:**
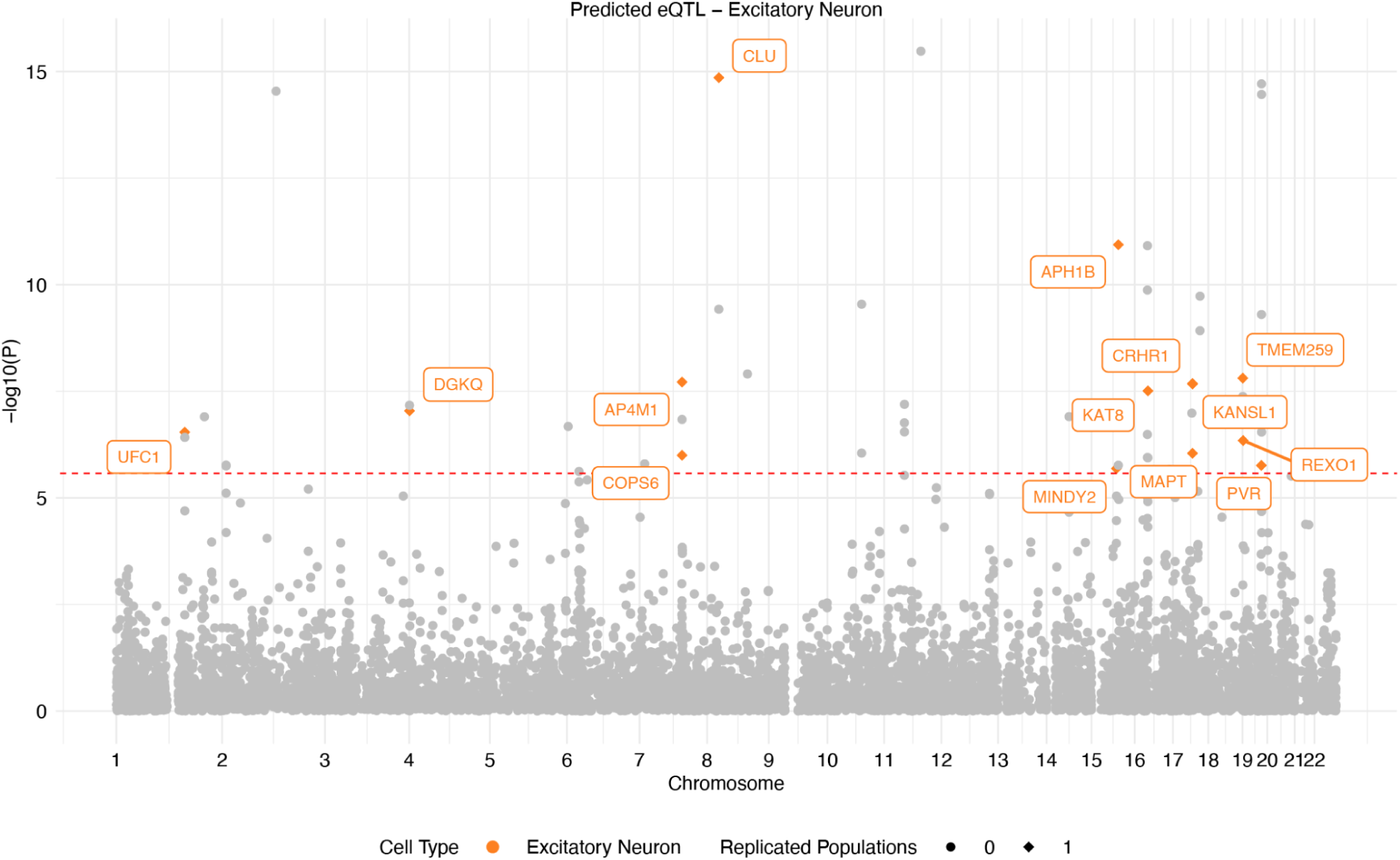
Manhattan plot of excitatory neuron-specific scEEMS predicted eMAGMA genes after filtering out fine-mapped eMAGMA and MAGMA genes in the European AD GWAS. Gray: no replication in non-European populations; colored: replicated (P < 0.05) in non-European populations. Circles: zero replicated populations; diamonds: one replicated population; squares: two replicated populations. Color indicates cell type.

**Supplementary Figure 26:**
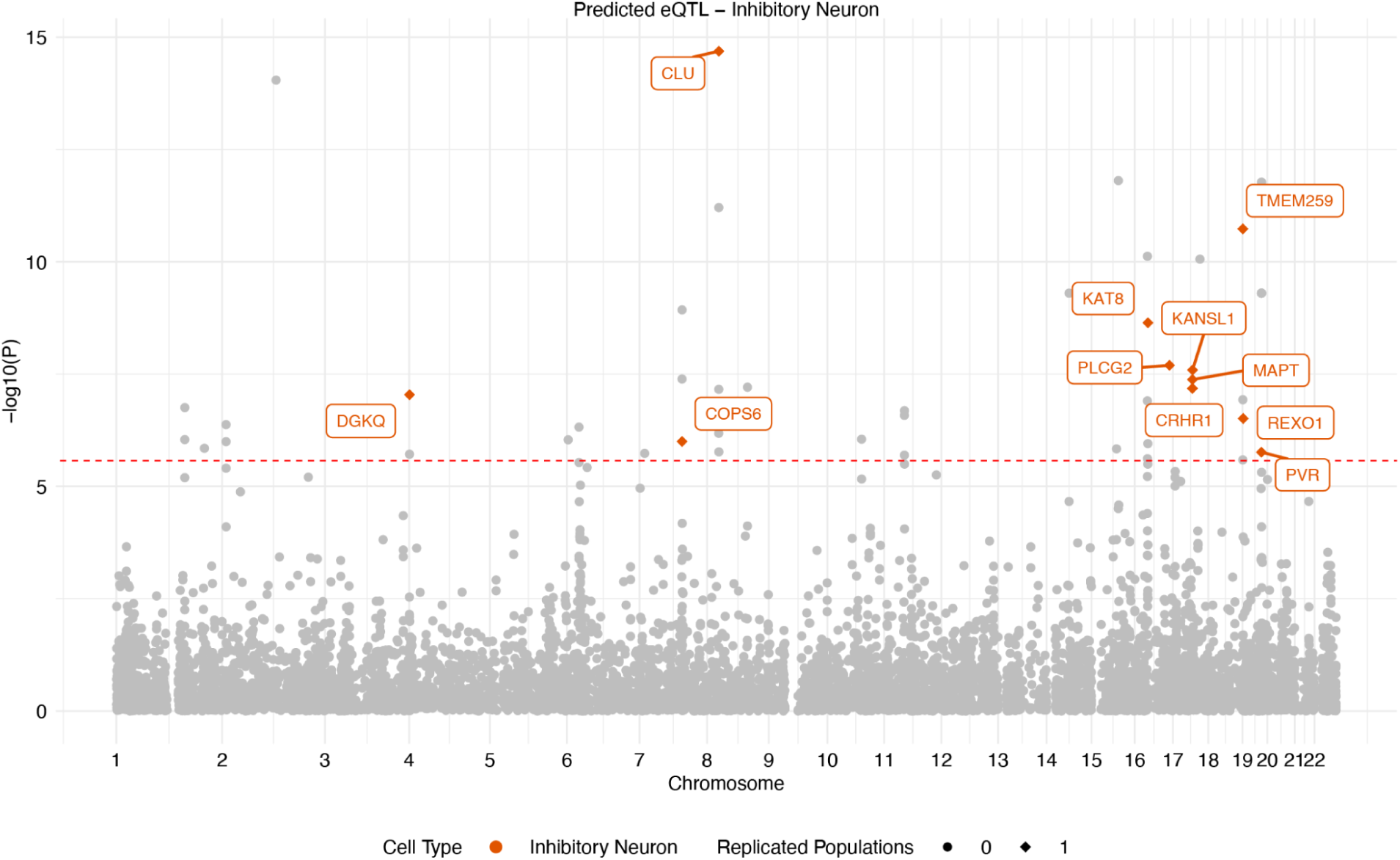
Manhattan plot of inhibitory neuron-specific scEEMS predicted eMAGMA genes after filtering out fine-mapped eMAGMA and MAGMA genes in the European AD GWAS. Gray: no replication in non-European populations; colored: replicated (P < 0.05) in non-European populations. Circles: zero replicated populations; diamonds: one replicated population; squares: two replicated populations. Color indicates cell type.

**Supplementary Figure 27:**
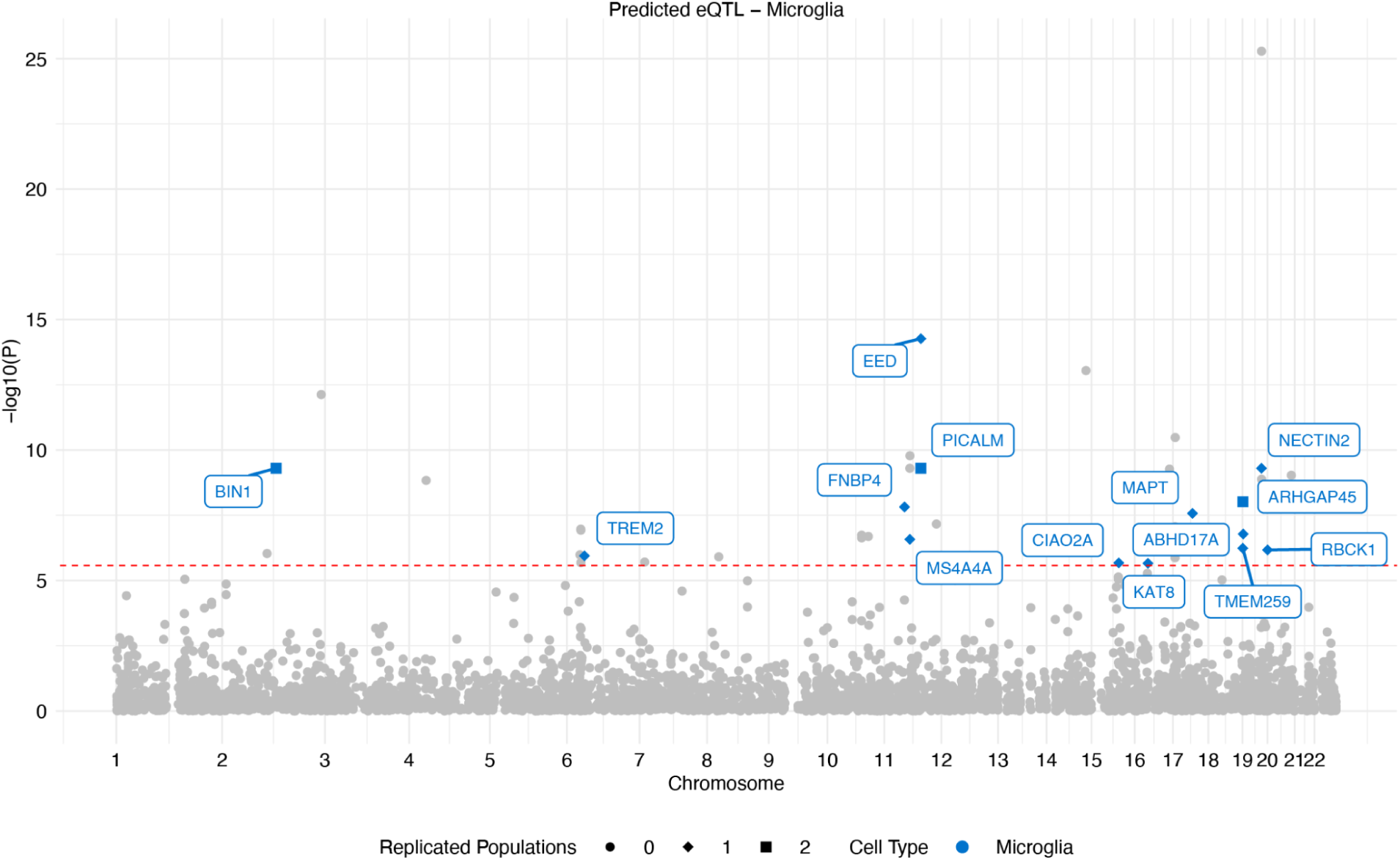
Manhattan plot of microglia-specific scEEMS predicted eMAGMA genes after filtering out fine-mapped eMAGMA and MAGMA genes in the European AD GWAS. Gray: no replication in non-European populations; colored: replicated (P < 0.05) in non-European populations. Circles: zero replicated populations; diamonds: one replicated population; squares: two replicated populations. Color indicates cell type.

**Supplementary Figure 28:**
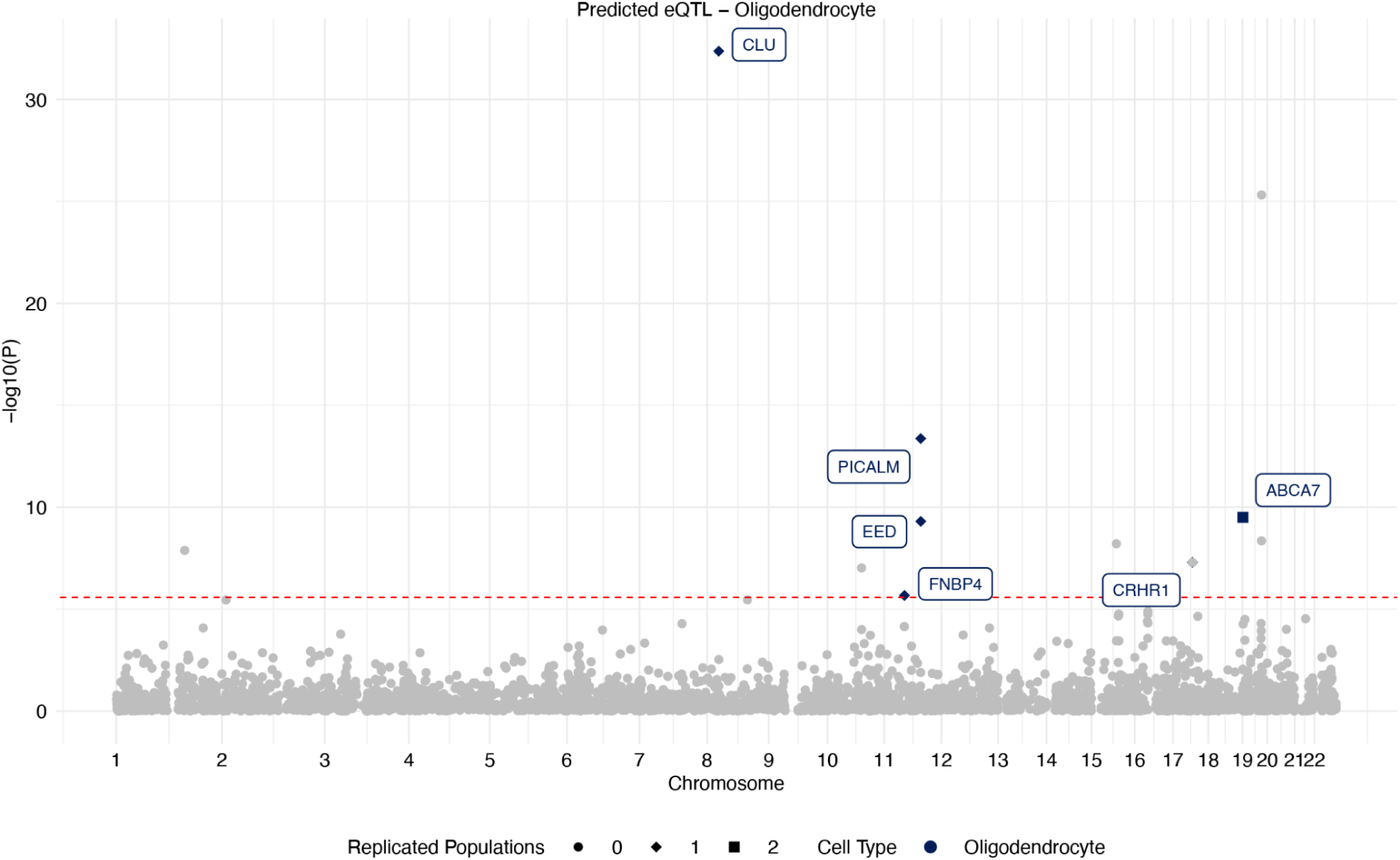
Manhattan plot of oligodendrocyte-specific scEEMS predicted eMAGMA genes after filtering out fine-mapped eMAGMA and MAGMA genes in the European AD GWAS. Gray: no replication in non-European populations; colored: replicated (P < 0.05) in non-European populations. Circles: zero replicated populations; diamonds: one replicated population; squares: two replicated populations. Color indicates cell type.

**Supplementary Figure 29:**
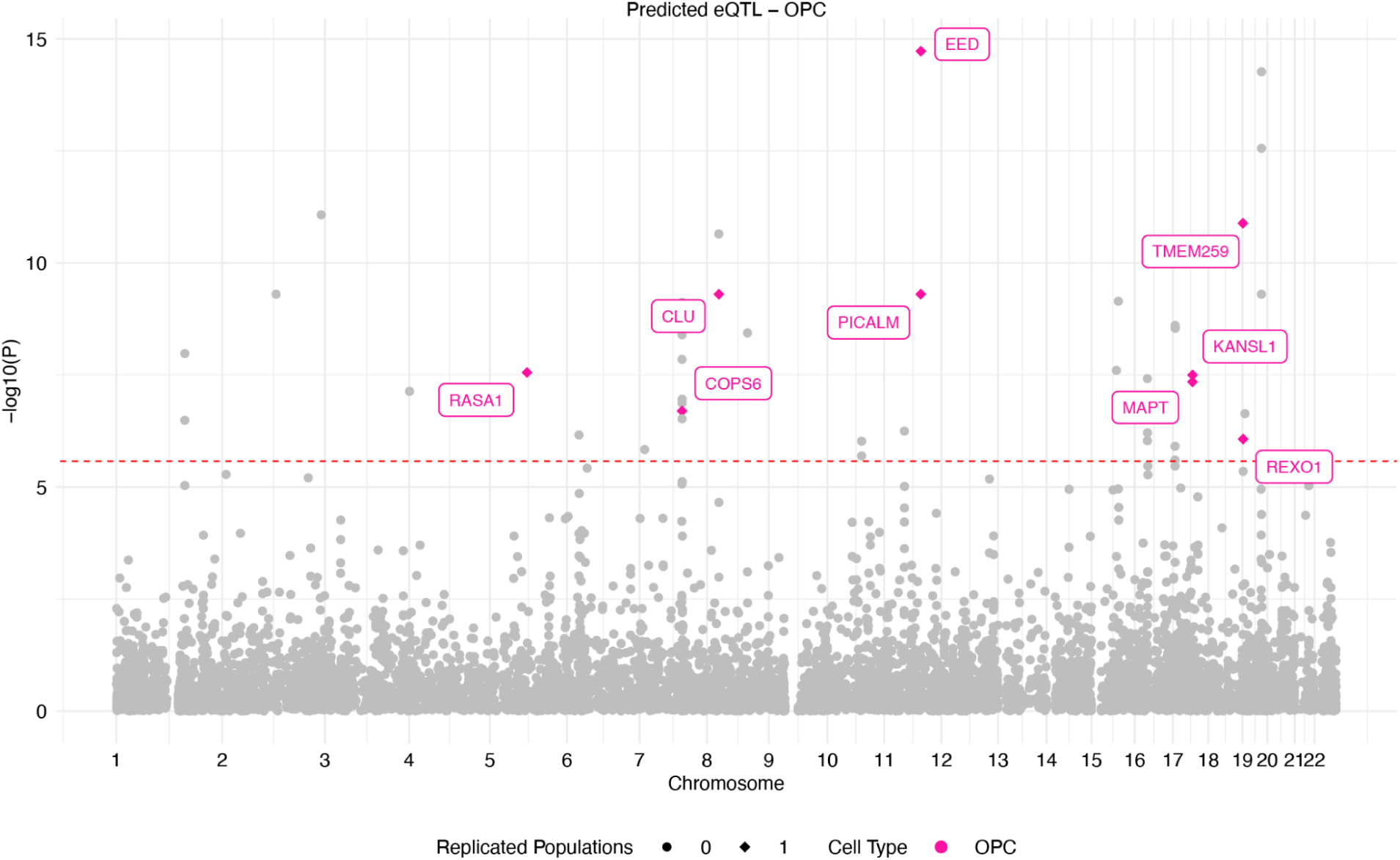
Manhattan plot of OPC-specific scEEMS predicted eMAGMA genes after filtering out fine-mapped eMAGMA and MAGMA genes in the European AD GWAS. Gray: no replication in non-European populations; colored: replicated (P < 0.05) in non-European populations. Circles: zero replicated populations; diamonds: one replicated population; squares: two replicated populations. Color indicates cell type.

**Supplementary Figure 30:**
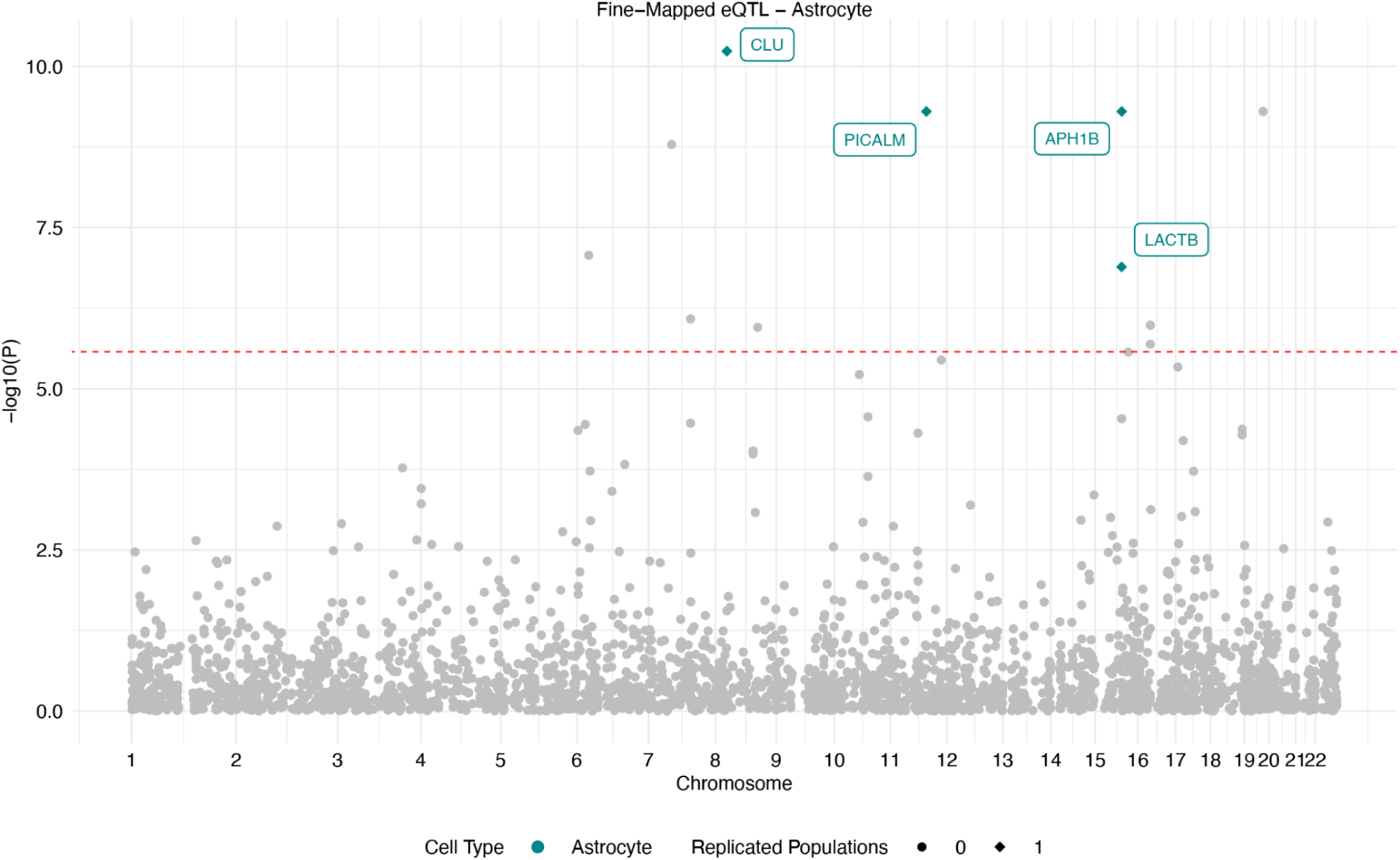
Manhattan plot of astrocyte-specific finemapped eMAGMA genes after filtering out fine-mapped eMAGMA and MAGMA genes in the European AD GWAS. Gray: no replication in non-European populations; colored: replicated (P < 0.05) in non-European populations. Circles: zero replicated populations; diamonds: one replicated population; squares: two replicated populations. Color indicates cell type.

**Supplementary Figure 31:**
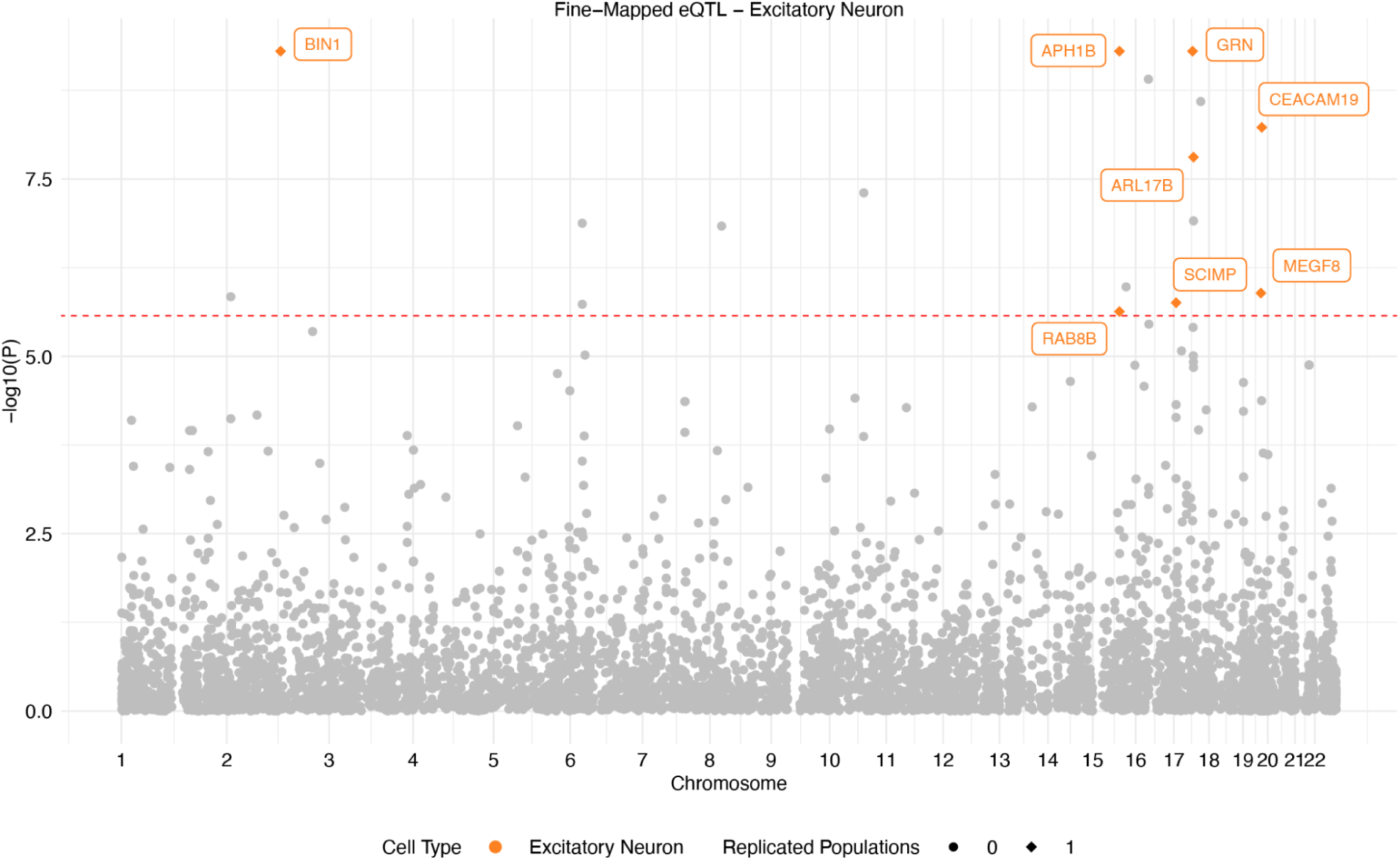
Manhattan plot of excitatory neuron-specific finemapped eMAGMA genes after filtering out fine-mapped eMAGMA and MAGMA genes in the European AD GWAS. Gray: no replication in non-European populations; colored: replicated (P < 0.05) in non-European populations. Circles: zero replicated populations; diamonds: one replicated population; squares: two replicated populations. Color indicates cell type.

**Supplementary Figure 32:**
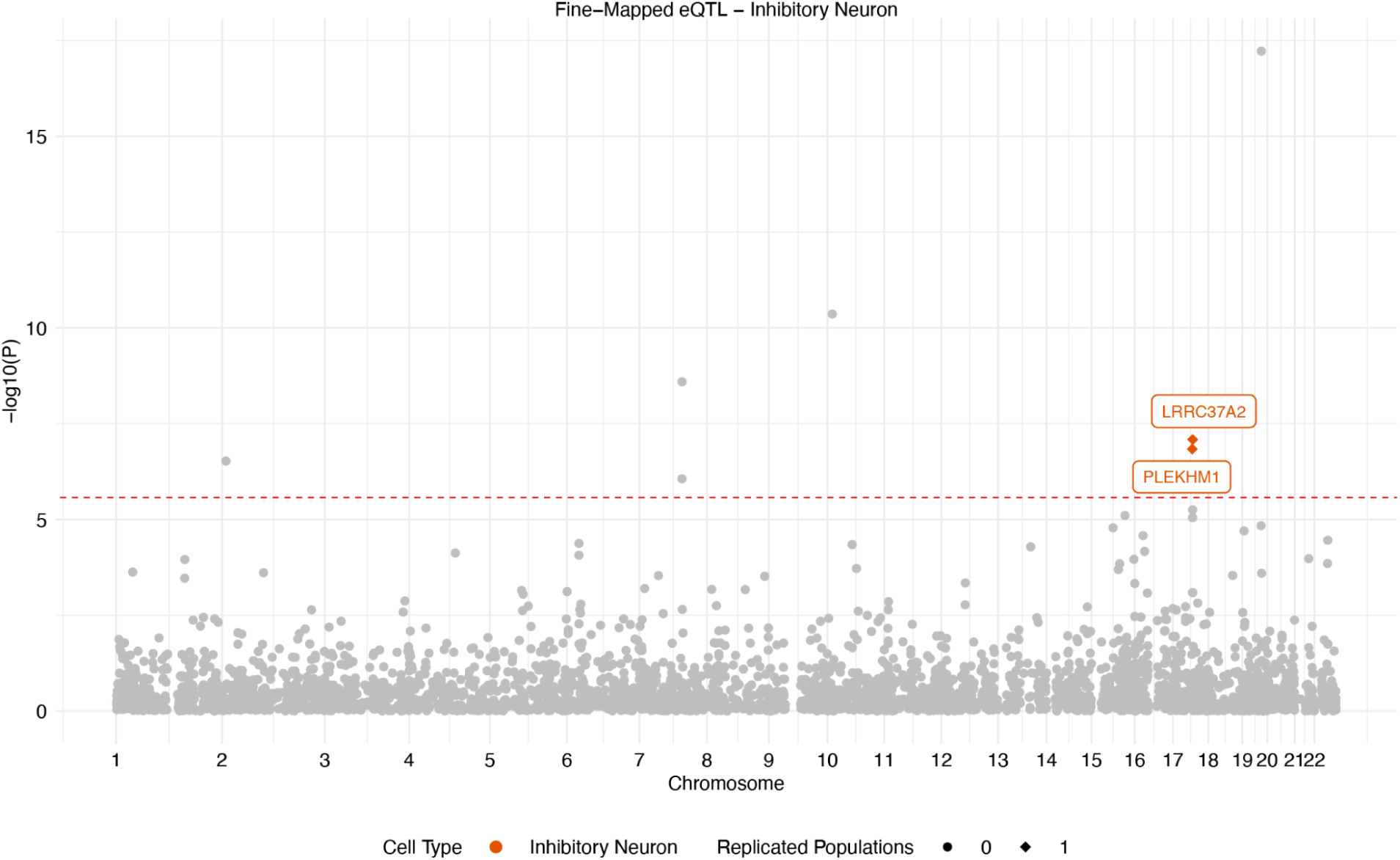
Manhattan plot of inhibitory neuron-specific finemapped eMAGMA genes after filtering out fine-mapped eMAGMA and MAGMA genes in the European AD GWAS. Gray: no replication in non-European populations; colored: replicated (P < 0.05) in non-European populations. Circles: zero replicated populations; diamonds: one replicated population; squares: two replicated populations. Color indicates cell type.

**Supplementary Figure 33:**
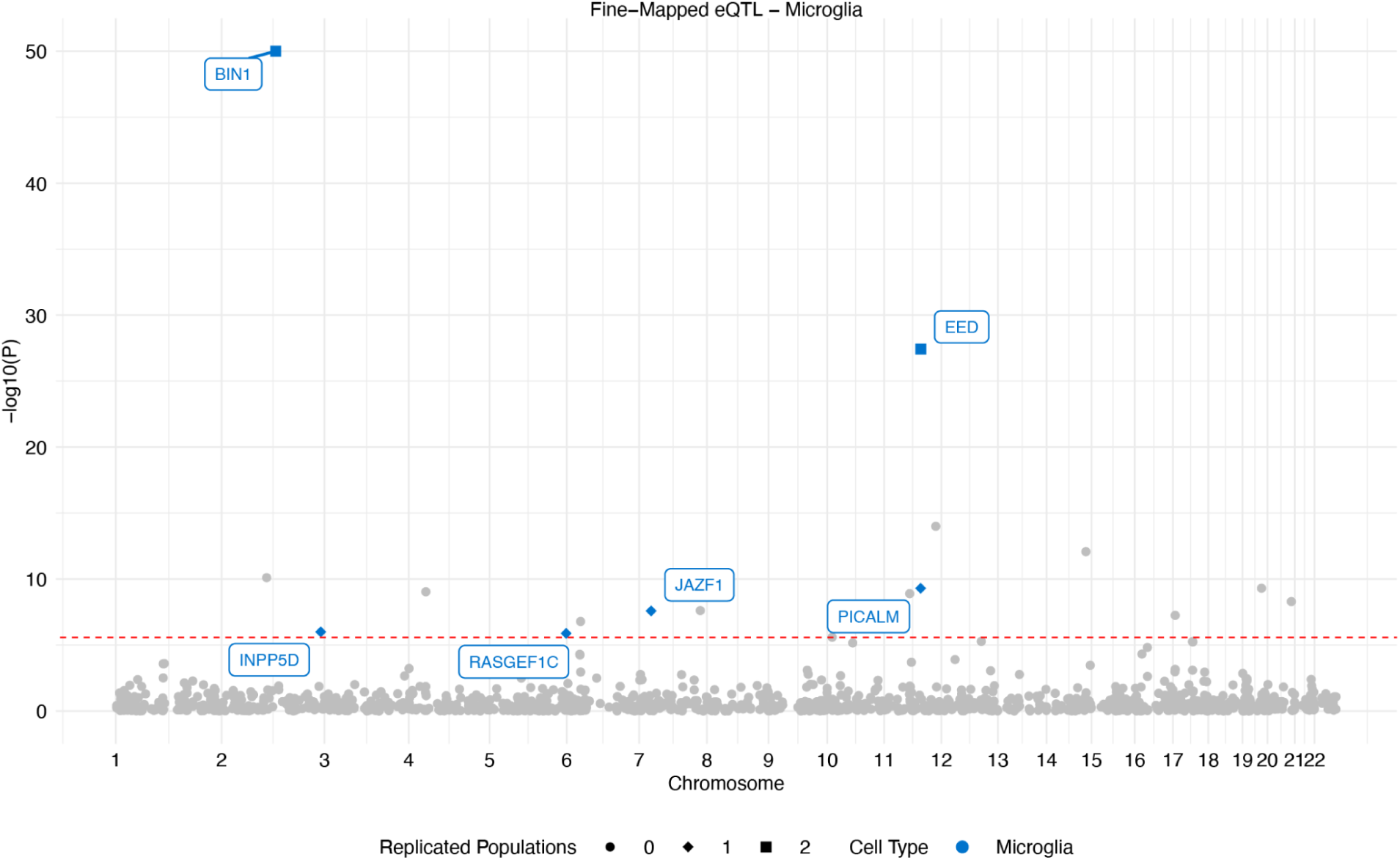
Manhattan plot of microglia-specific finemapped eMAGMA genes after filtering out fine-mapped eMAGMA and MAGMA genes in the European AD GWAS. Gray: no replication in non-European populations; colored: replicated (P < 0.05) in non-European populations. Circles: zero replicated populations; diamonds: one replicated population; squares: two replicated populations. Color indicates cell type.

**Supplementary Figure 34:**
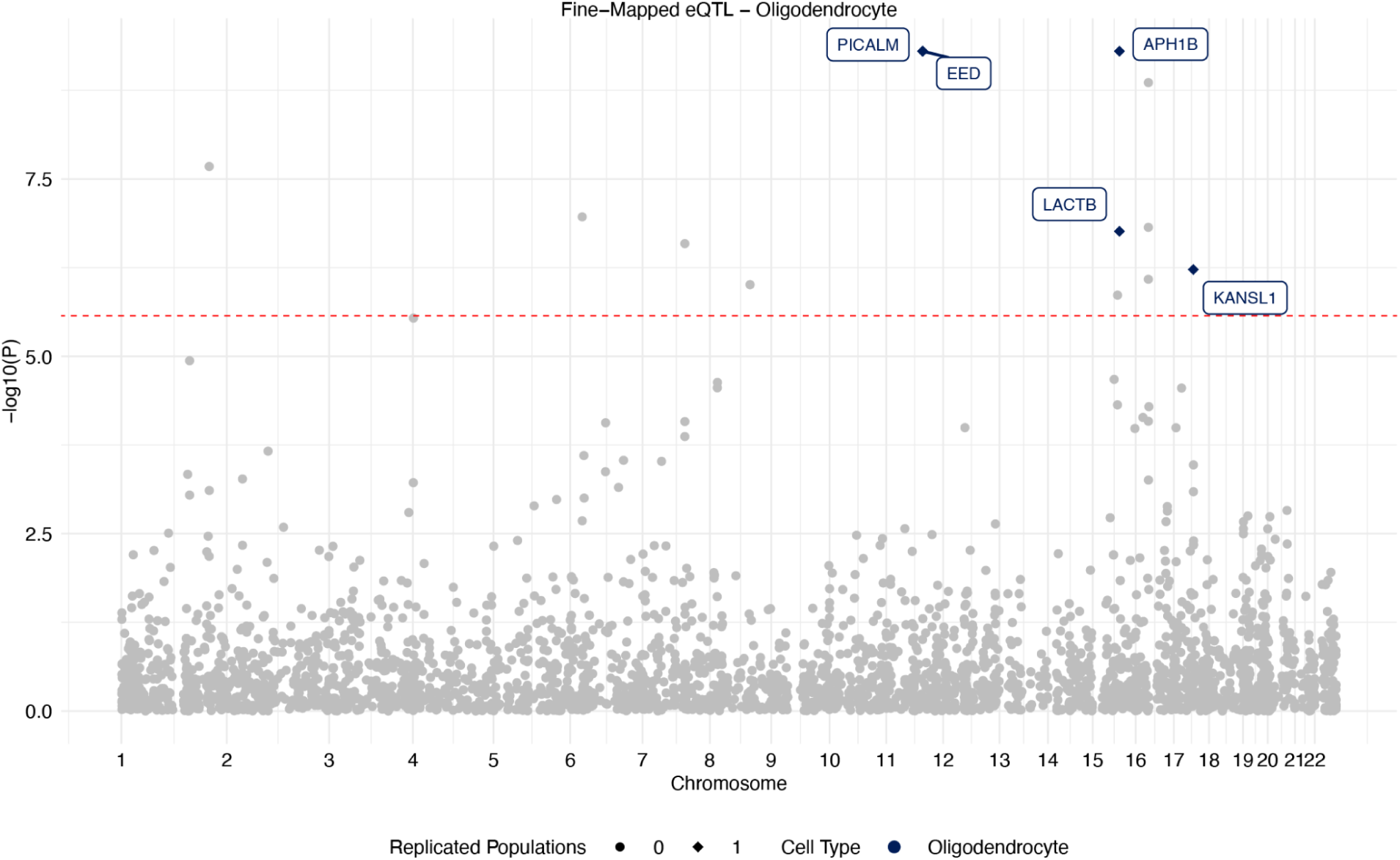
Manhattan plot of oligodendrocyte-specific finemapped eMAGMA genes after filtering out fine-mapped eMAGMA and MAGMA genes in the European AD GWAS. Gray: no replication in non-European populations; colored: replicated (P < 0.05) in non-European populations. Circles: zero replicated populations; diamonds: one replicated population; squares: two replicated populations. Color indicates cell type.

**Supplementary Figure 35:**
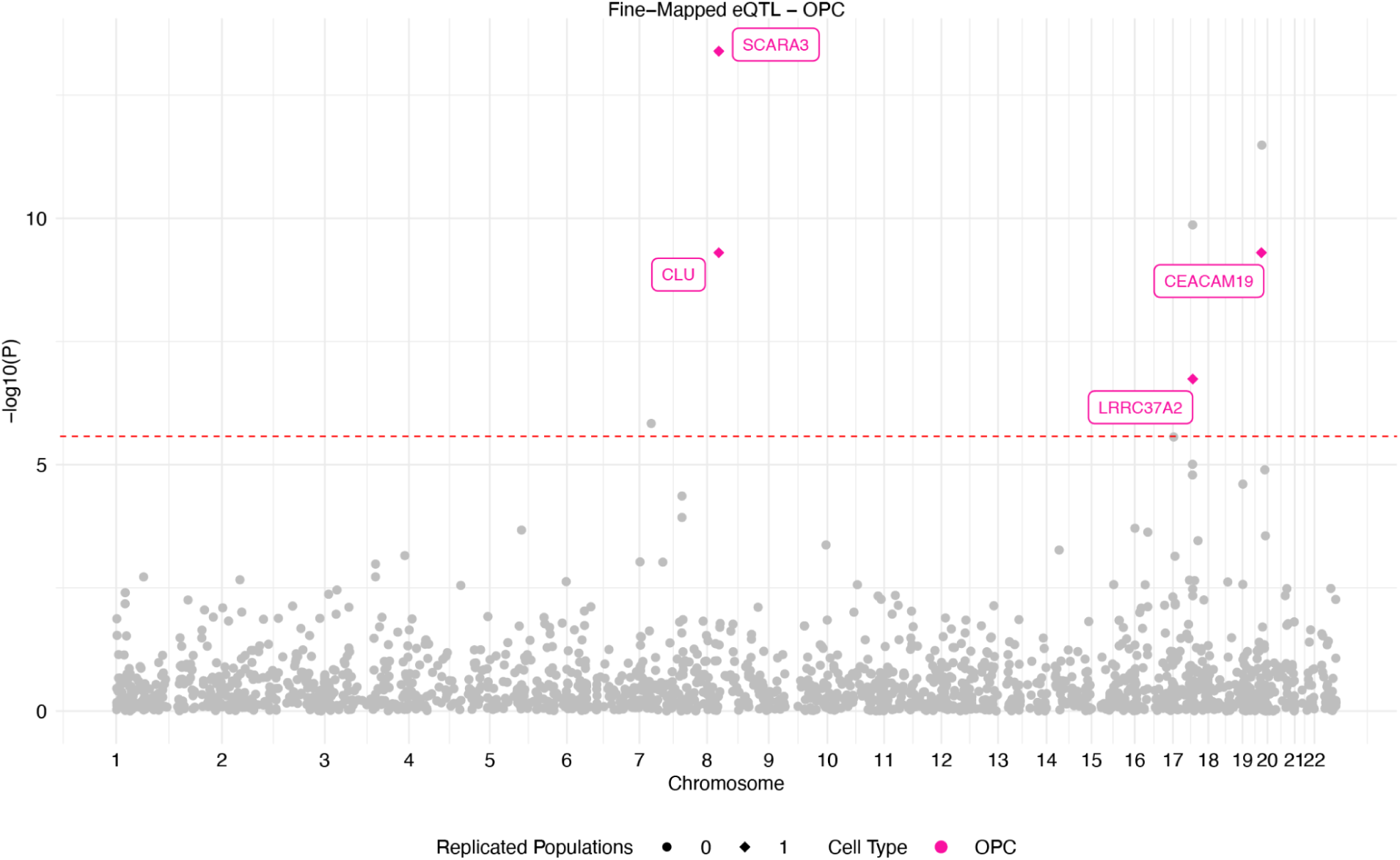
Manhattan plot of OPC-specific finemapped eMAGMA genes after filtering out fine-mapped eMAGMA and MAGMA genes in the European AD GWAS. Gray: no replication in non-European populations; colored: replicated (P < 0.05) in non-European populations. Circles: zero replicated populations; diamonds: one replicated population; squares: two replicated populations. Color indicates cell type.

**Supplementary Figure 36:**
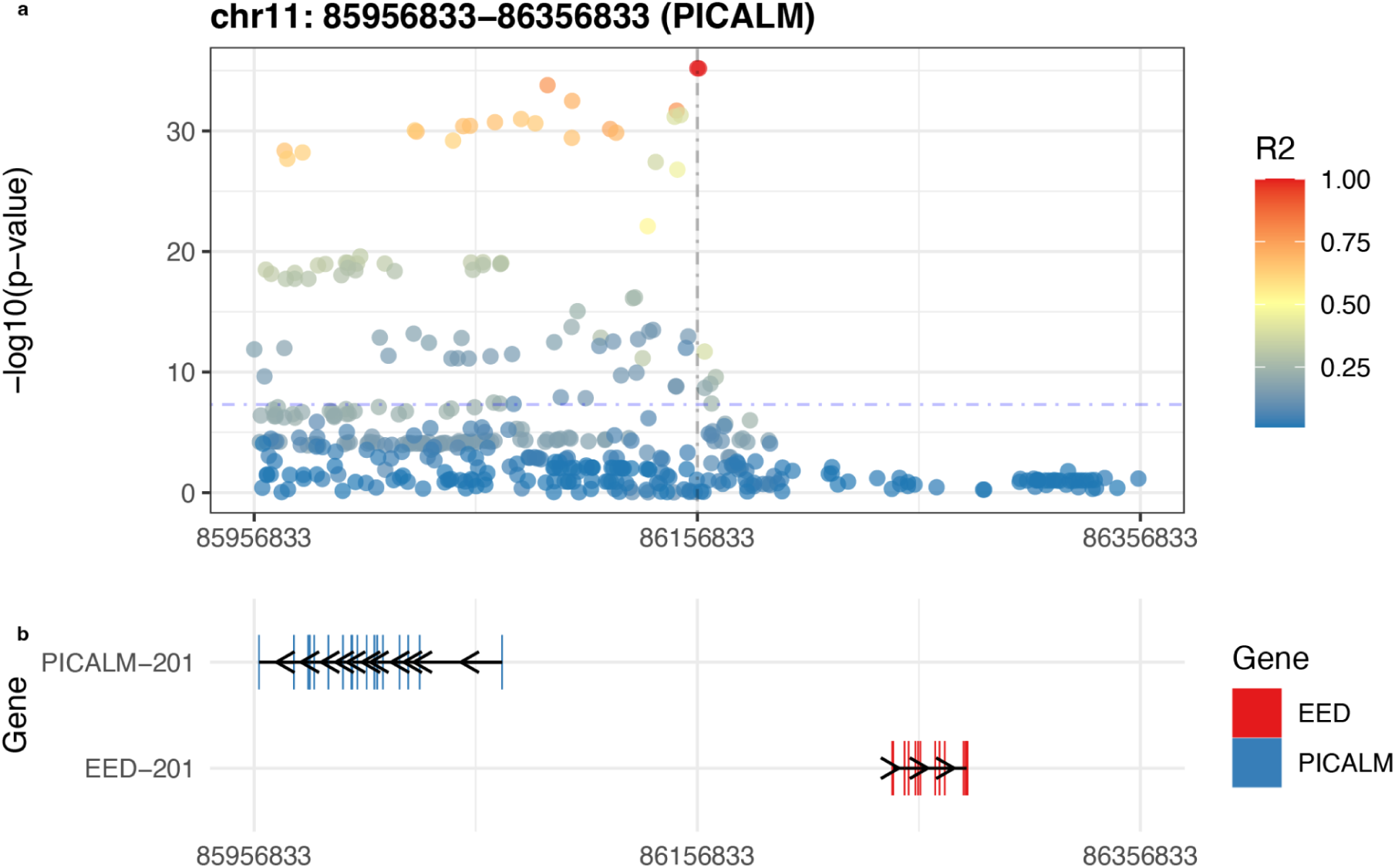
Locus zoom plot of AD GWAS risk variant in the *PICALM*/*EED* locus.

**Supplementary Figure 37:**
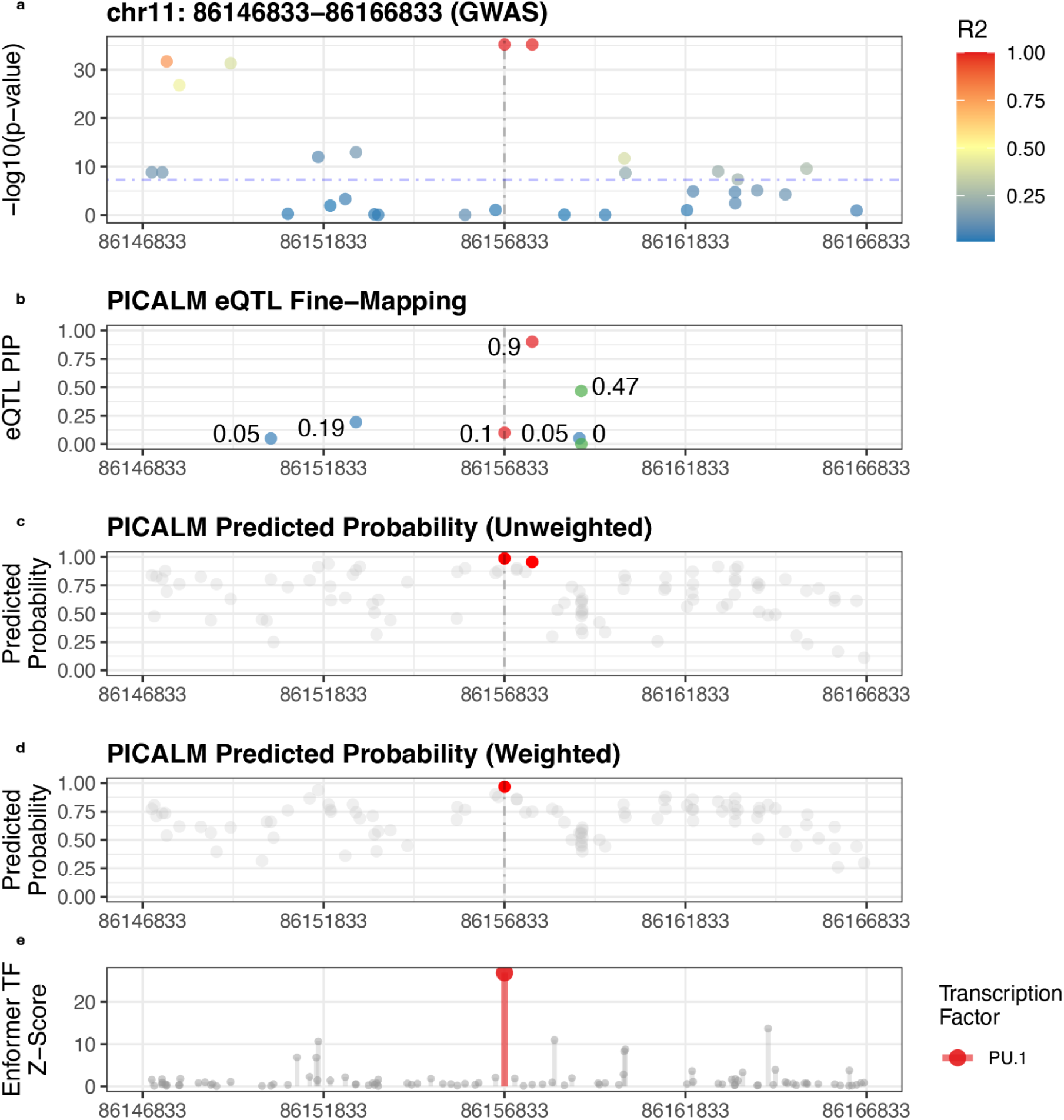
Zoomed in view of locus zoom plot of AD GWAS risk variant in the *PICALM*/*EED* locus. a) -log10(p-value) of the association statistics in this region centered at the variant *rs10792832* with variant *rs3851179* to the right in very high LD. b) PIP values of the *PICALM* eQTL fine-mapping for *rs10792832* and *rs3851179*. c) the predicted probabilities using the DL-VEP unweighted model for microglia for *rs10792832* and *rs3851179*. d) the predicted probabilities using the DL-VEP upweighted model for microglia for *rs10792832* and *rs3851179*. e) the Enformer DL-VEP score for *PU.1* TF binding, which overlaps with the *rs10792832* implying this variant decreases *PU.1* TF binding affinity.

**Supplementary Figure 38:**
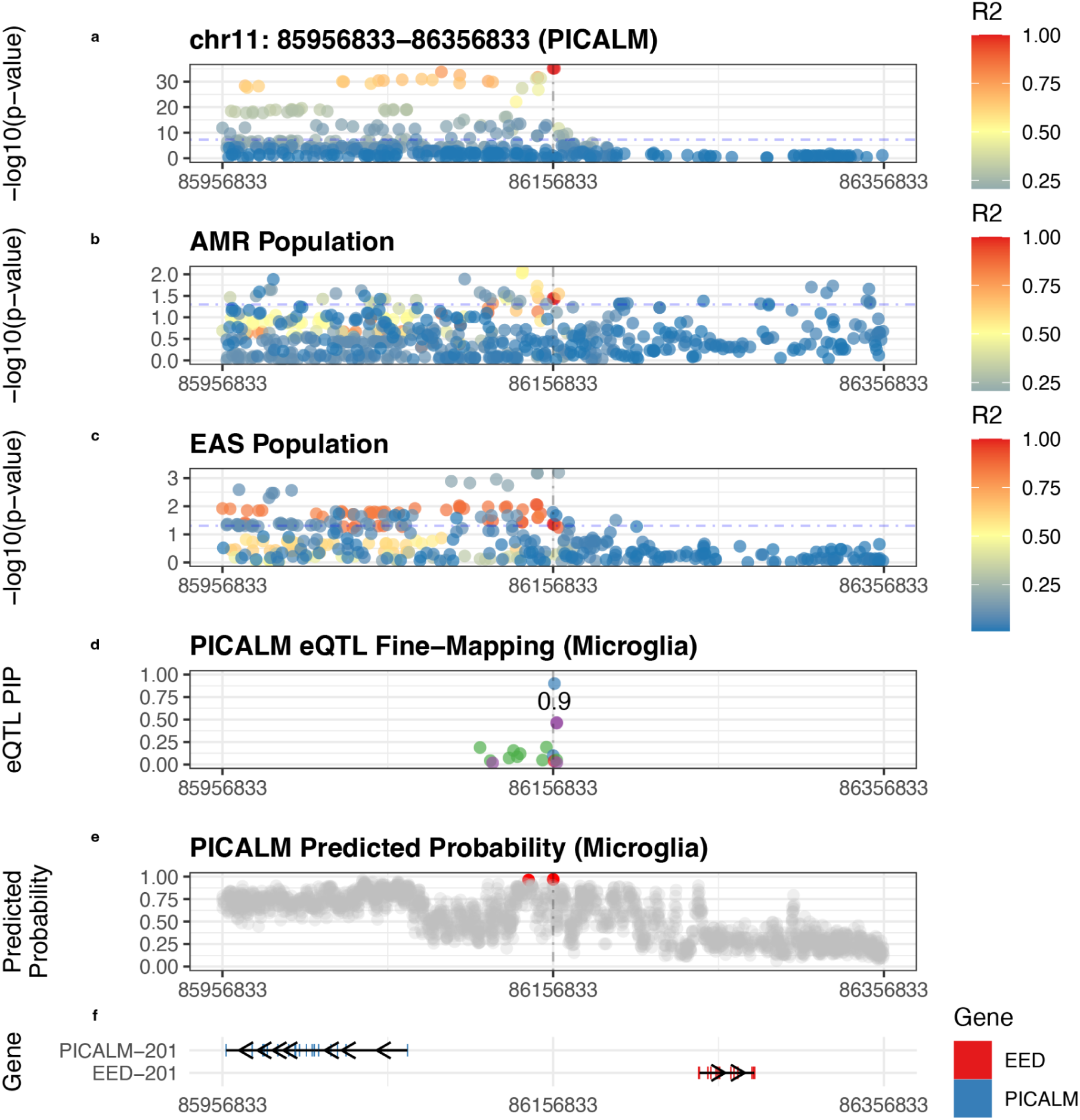
Zoomed out view of locus zoom plot of AD GWAS risk variant in the *PICALM*/*EED* locus. a) -log10(p-value) of the association statistics in this region centered at the variant *rs10792832* along with other variants in this region. b) -log10(p-value) of the association statistics in this region centered at the variant *rs10792832* along with other variants in this region using summary statistics from the AMR AD GWAS. c) -log10(p-value) of the association statistics in this region centered at the variant *rs10792832* along with other variants in this region using summary statistics from the EAS AD GWAS. d) PIP values for multiple credible sets found by *PICALM* eQTL fine-mapping, points in blue correspond to the credible set associated with *rs10792832* and other colors represent other credible sets. e) the predicted probabilities from the microglia scEEMS model for variants in this region. Points in red represent variants with predicted probability > 0.95. f) the position of these variants within the integenic region between the *PICALM* and EED genes.

**Supplementary Figure 39:**
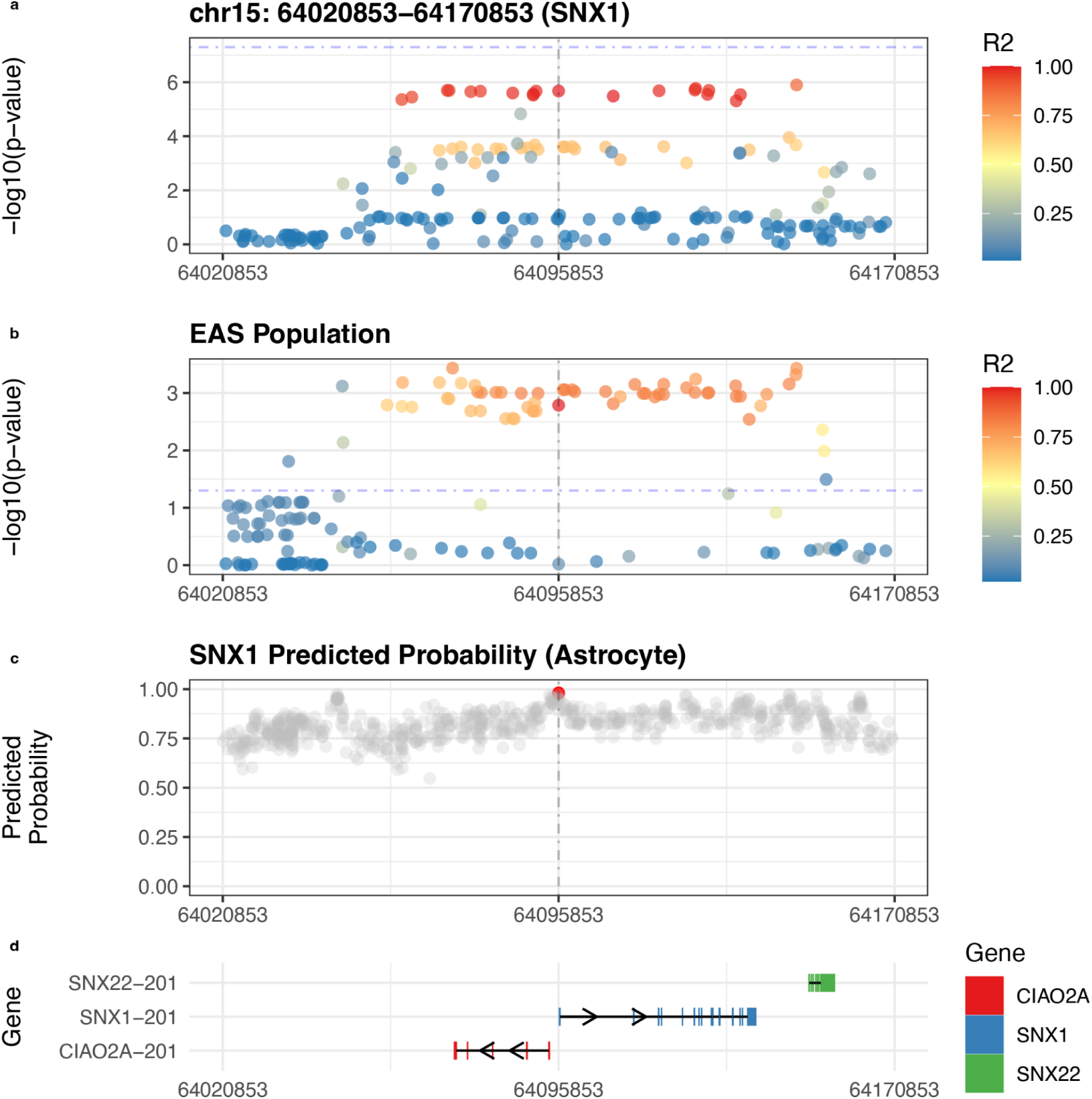
Zoomed in view of locus zoom plot of AD GWAS risk variant in the *SNX1* locus. a) -log10(p-value) of the association statistics in this region centered at the variant *rs332258* which is below the GWAS significant threshold denoted by the dashed line. b) -log10(p-value) of the association statistics in this region centered at the variant *rs332258* along with other variants in this region using summary statistics from the AMR AD GWAS. The variant *rs332258* is above the statistically significant threshold denoted by the dashed line. c) the predicted probabilities using the astrocyte scEEMS model for astrocyte where *rs332258* (red) is the only variant with predicted probability > 0.97. d) the position of these variants within the *SNX1* locus.

**Supplementary Figure 40:**
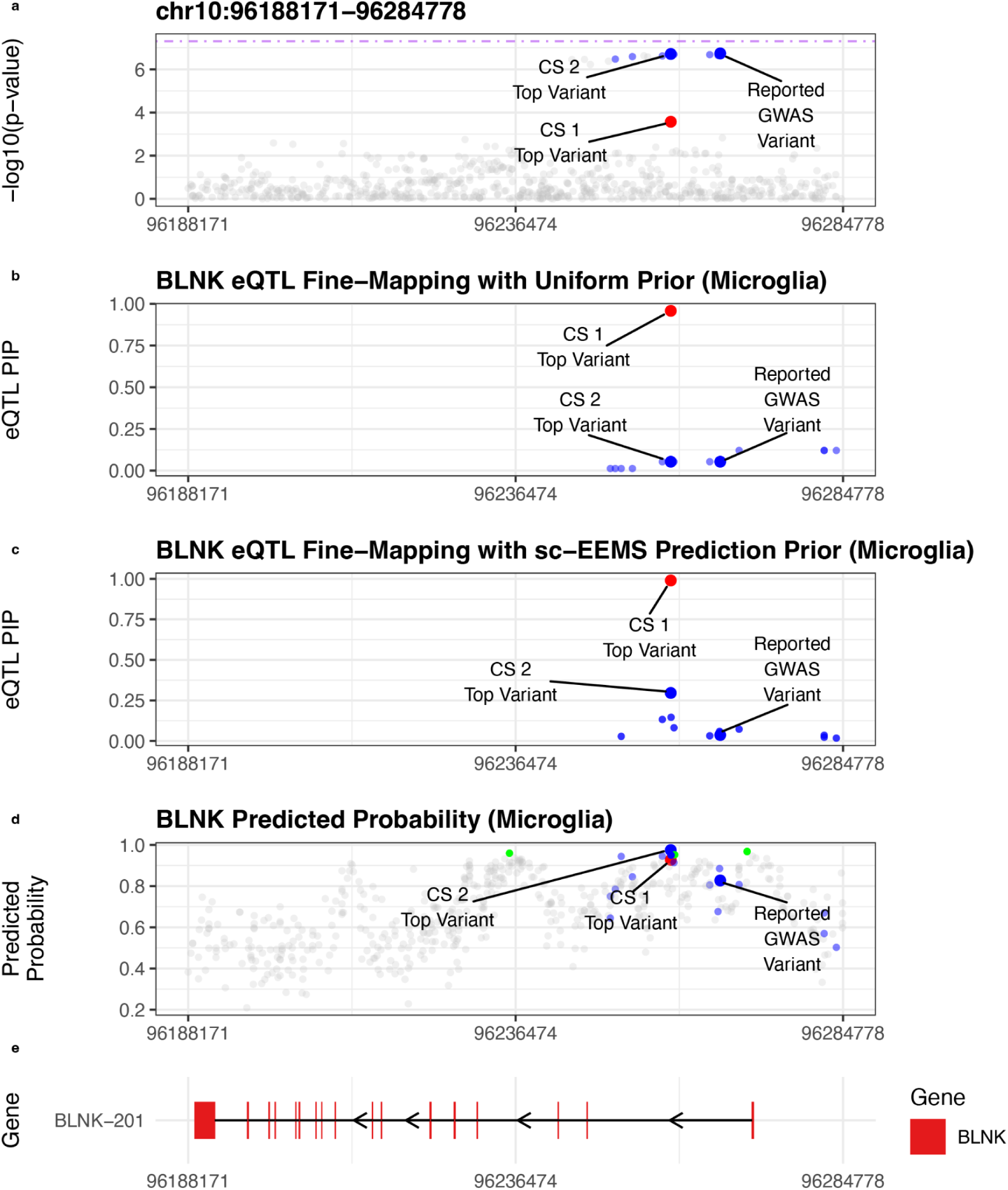
Zoomed in view of locus zoom plot of AD GWAS risk variant in the *BLNK* locus. a) -log10(p-value) of the association statistics in the *BLNK* region showing the reported AD GWAS variant^11^ as well as the top fine-mapped variants from two microglia *BLNK* eQTL credible sets. b) Microglia *BLNK* eQTL credible sets for two independent credible sets fine-mapped using a uniform prior. c) Microglia *BLNK* eQTL credible sets for two independent credible sets fine-mapped using the scEEMS prediction prior. d) the predicted probabilities using the microglia scEEMS model showing the variants mapped to the two eQTL credible sets as well as all other variants (green) with predicted probability above the microglia-specific prediction probability threshold. e) Locus plot showing the location of the *BLNK* gene.

**Supplementary Figure 41:**
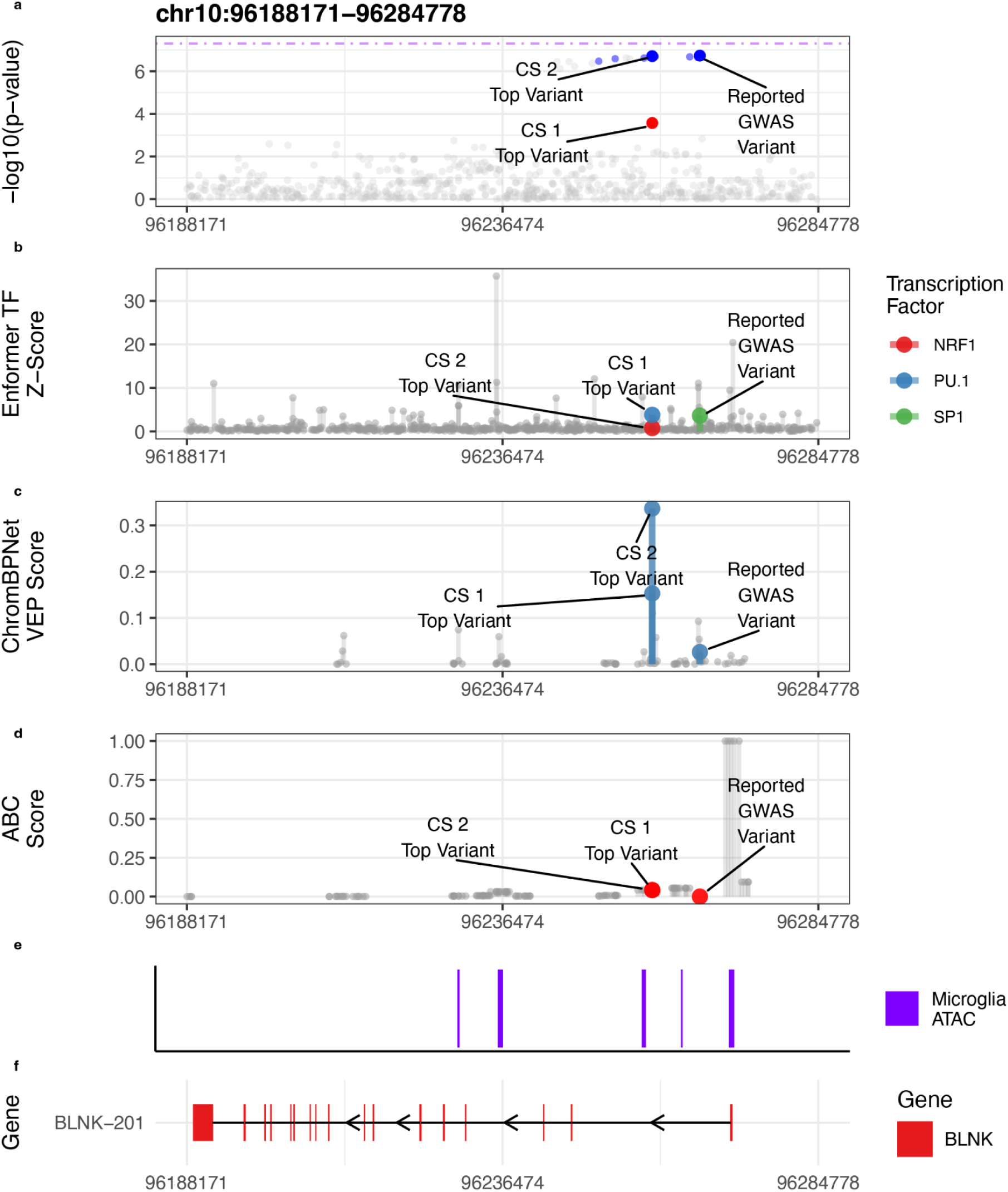
Zoomed in view of locus zoom plot of AD GWAS risk variant in the *BLNK* locus. a) -log10(p-value) of the association statistics in the *BLNK* region showing the reported AD GWAS variant^11^ as well as the top fine-mapped variants from two microglia *BLNK* eQTL credible sets. b) Highest Enformer DL-VEP scores for microglia-specific TFs Microglia *BLNK* eQTL credible sets for reported AD GWAS variant^11^ as well as the top fine-mapped variants from two microglia *BLNK* eQTL credible sets. c) ChromBPNet DL-VEP scores (trained on microglia ATAC-seq data) for reported AD GWAS variant^11^ as well as the top fine-mapped variants from two microglia *BLNK* eQTL credible sets. d) Microglia ABC scores for reported AD GWAS variant^11^ as well as the top fine-mapped variants from two microglia *BLNK* eQTL credible sets. e) Locus plot showing the location of the *BLNK* gene.

**Supplementary Figure 41:**
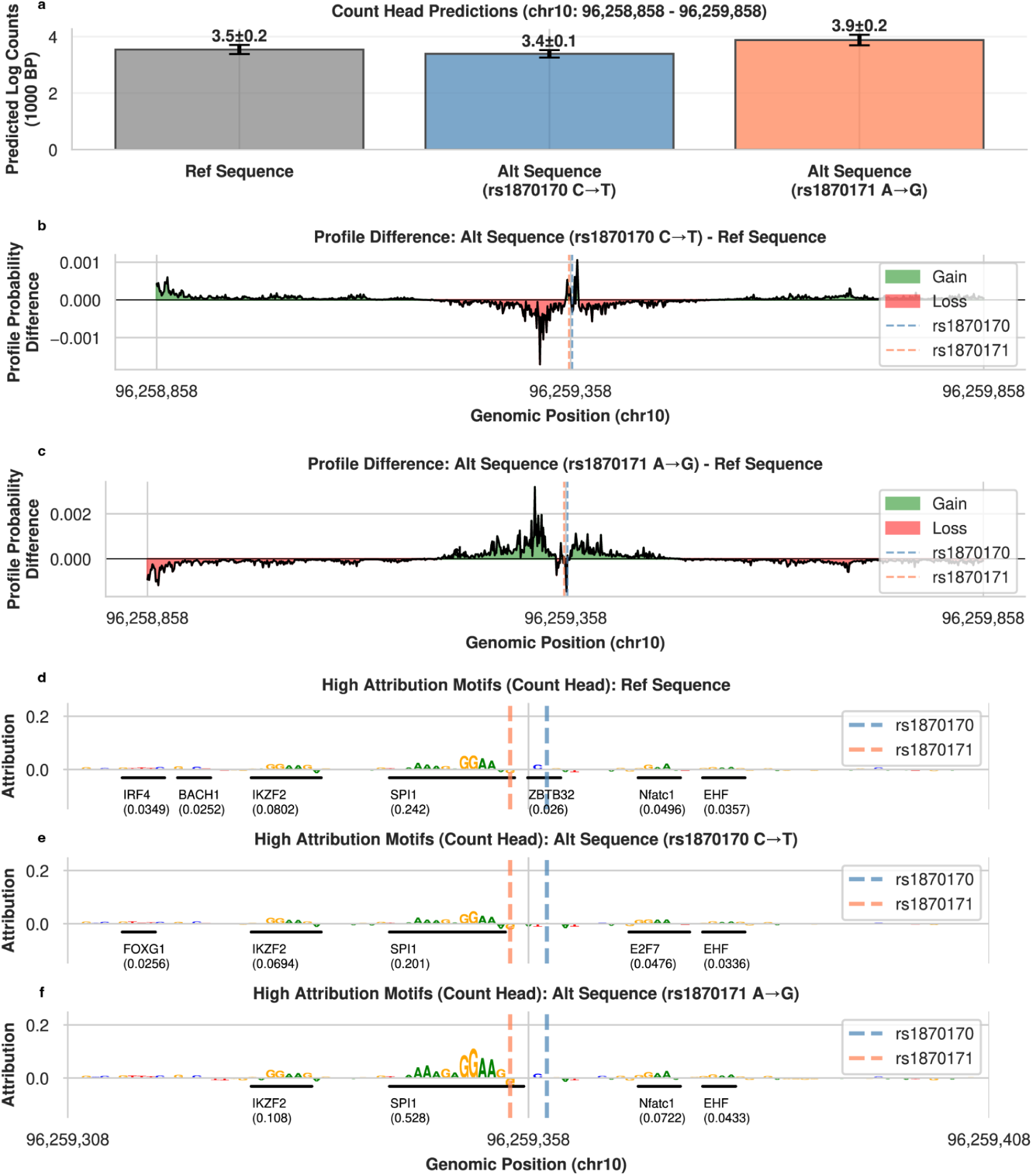
a) ChromBPNet Predictions of microglia ATAC-seq read counts (log transformed) for reference sequence, and alternate sequences corresponding to the variants *rs1870170* and *rs1870171* (referene allele is replaced with alternate allele). b) Difference in ChromBPNet profile predictions between alternate sequence for *rs1870170* and the reference sequence. c) Difference in ChromBPNet profile predictions between alternerate sequence for *rs1870171* and the reference sequence. d) Motif attribution plot for ChromBPNet microglia ATAC-seq read counts for the reference sequence. e) Motif attribution plot for ChromBPNet microglia ATAC-seq read counts of alternerate sequence for *rs1870170*. f) Motif attribution plot for ChromBPNet microglia ATAC-seq read counts of alternerate sequence for *rs1870171*.

## Supplementary Tables

Supplementary Table 1 - Cohort Statistics

Supplementary Table 2 - List of model features

Supplementary Table 3 - eMAGMA results - predicted eQTLs

Supplementary Table 4 - eMAGMA results - finemapped eQTLs

Supplementary Table 5 - MAGMA results

Supplementary Table 6 - Summary Statistics for *BLNK* Locus

## Code Availability

Instructions for model training are available at https://statfungen.github.io/xqtl-protocol/code/xqtl_modifier_score/ems_training.html

## Acknowledgements

We would like to thank Jonathan Pritchard and Luke O’Connor for helpful discussions about the analysis for this paper. We would also like to thank Katie Cho and Angelina Siagailo for their contributions to developing reusable training and prediction protocols for scEEMS.

## Author Contributions

D.A.K. and G.W. proposed the idea for the project. D.A.K., G.W., and C.M.L. developed the scEEMS model. C.M.L., G.C., A.L., R.N., and R.F. performed the analysis. T.R. and P.D.J. provided data for the project. D.A.K., G.W., and C.M.L. wrote the manuscript.

## References

1. GTEx Consortium. The GTEx Consortium atlas of genetic regulatory effects across human tissues. Science 369, 1318–1330 (2020).

2. Yao, D. W., O’Connor, L. J., Price, A. L. & Gusev, A. Quantifying genetic effects on disease mediated by assayed gene expression levels. Nat. Genet. 52, 626–633 (2020).

3. Connally, N. J. et al. The missing link between genetic association and regulatory function. Elife 11, (2022).

4. Mostafavi, H., Spence, J. P., Naqvi, S. & Pritchard, J. K. Systematic differences in discovery of genetic effects on gene expression and complex traits. Nat. Genet. 55, 1866–1875 (2023).

5. Umans, B. D., Battle, A. & Gilad, Y. Where Are the Disease-Associated eQTLs? Trends Genet. (2020) doi:10.1016/j.tig.2020.08.009.

6. Bryois, J. et al. Cell-type-specific cis-eQTLs in eight human brain cell types identify novel risk genes for psychiatric and neurological disorders. Nat. Neurosci. 25, 1104–1112 (2022).

7. Yazar, S. et al. Single-cell eQTL mapping identifies cell type-specific genetic control of autoimmune disease. Science 376, eabf3041 (2022).

8. Alasoo, K. et al. Shared genetic effects on chromatin and gene expression indicate a role for enhancer priming in immune response. Nat. Genet. 50, 424–431 (2018).

9. Fujita, M. et al. Cell subtype-specific effects of genetic variation in the Alzheimer’s disease brain. Nat. Genet. 56, 605–614 (2024).

10. Mathys, H. et al. Single-cell atlas reveals correlates of high cognitive function, dementia, and resilience to Alzheimer’s disease pathology. Cell 186, 4365–4385.e27 (2023).

11. Bellenguez, C. et al. New insights into the genetic etiology of Alzheimer’s disease and related dementias. Nat. Genet. 54, 412–436 (2022).

12. Jeong, R. & Bulyk, M. L. Chromatin accessibility variation provides insights into missing regulation underlying immune-mediated diseases. bioRxiv (2024) doi:10.1101/2024.04.12.589213.

13. Mu, Z. et al. Impact of disease-associated chromatin accessibility QTLs across immune cell types and contexts. medRxiv (2024) doi:10.1101/2024.12.05.24318552.

14. Zhou, J. & Troyanskaya, O. G. Predicting effects of noncoding variants with deep learning–based sequence model. Nat. Methods 12, 931–934 (2015).

15. Avsec, Ž., et al. Effective gene expression prediction from sequence by integrating long-range interactions. Nat. Methods 18, 1196–1203 (2021).

16. Linder, J., Srivastava, D., Yuan, H., Agarwal, V. & Kelley, D. R. Predicting RNA-seq coverage from DNA sequence as a unifying model of gene regulation. Nat. Genet. 57, 949–961 (2025).

17. Pampari, A. et al. ChromBPNet: bias factorized, base-resolution deep learning models of chromatin accessibility reveal cis-regulatory sequence syntax, transcription factor footprints and regulatory variants. bioRxivorg (2025) doi:10.1101/2024.12.25.630221.

18. Avsec, Ž., et al. Base-resolution models of transcription-factor binding reveal soft motif syntax. Nat. Genet. 53, 354–366 (2021).

19. Roadmap Epigenomics Consortium et al. Integrative analysis of 111 reference human epigenomes. Nature 518, 317–330 (2015).

20. ENCODE Project Consortium et al. Expanded encyclopaedias of DNA elements in the human and mouse genomes. Nature 583, 699–710 (2020).

21. Wang, Q. S. et al. Leveraging supervised learning for functionally informed fine-mapping of cis-eQTLs identifies an additional 20,913 putative causal eQTLs. Nat. Commun. 12, 3394 (2021).

22. Fulco, C. P. et al. Activity-by-contact model of enhancer-promoter regulation from thousands of CRISPR perturbations. Nat. Genet. 51, 1664–1669 (2019).

23. Nasser, J. et al. Genome-wide enhancer maps link risk variants to disease genes. Nature 593, 238–243 (2021).

24. Zeng, T., Spence, J. P., Mostafavi, H. & Pritchard, J. K. Bayesian estimation of gene constraint from an evolutionary model with gene features. Nat. Genet. 56, 1632–1643 (2024).

25. Prokhorenkova, L., Gusev, G., Vorobev, A., Dorogush, A. V. & Gulin, A. CatBoost: unbiased boosting with categorical features. arXiv [cs.LG] (2017).

26. Bennett, D. A. et al. Religious Orders Study and Rush Memory and Aging Project. J. Alzheimers. Dis. 64, S161–S189 (2018).

27. Leung, Y. Y. FunGen-xQTL atlas: a multi-omic atlas of quantitative trait loci in human brain and cerebrospinal fluid for target identification and prioritization in Alzheimer’s disease. Alzheimer’s & Dementia 20, e092045 (2024).

28. Cao, X. et al. Integrative multi-omics QTL colocalization maps regulatory architecture in aging human brain. medRxiv (2025) doi:10.1101/2025.04.17.25326042.

29. Beecham, G. W. et al. The Alzheimer’s Disease Sequencing Project: Study design and sample selection. Neurol. Genet. 3, e194 (2017).

30. Lundberg, S. M. & Lee, S.-I. A Unified Approach to Interpreting Model Predictions. in Advances in Neural Information Processing Systems (eds Guyon, I. et al.) vol. 30 (Curran Associates, Inc., 2017).

31. Lundberg, S. M. et al. From local explanations to global understanding with explainable AI for trees. *Nat*. Mach. Intell. 2, 56–67 (2020).

32. Gerring, Z. F., Mina-Vargas, A., Gamazon, E. R. & Derks, E. M. E-MAGMA: an eQTL-informed method to identify risk genes using genome-wide association study summary statistics. Bioinformatics 37, 2245–2249 (2021).

33. Rajabli, F. et al. Multi-ancestry genome-wide meta-analysis of 56,241 individuals identifies known and novel cross-population and ancestry-specific associations as novel risk loci for Alzheimer’s disease. Genome Biol. 26, 210 (2025).

34. Comandante-Lou, N., et al. PLXNB1 and other signaling drives a pathologic astrocyte state contributing to cognitive decline in Alzheimer’s Disease. bioRxivorg (2025) doi:10.1101/2025.02.24.639868.

35. Green, G. S. et al. Cellular communities reveal trajectories of brain ageing and Alzheimer’s disease. Nature 633, 634–645 (2024).

36. Wang, G., Sarkar, A., Carbonetto, P. & Stephens, M. A simple new approach to variable selection in regression, with application to genetic fine mapping. J. R. Stat. Soc. Series B Stat. Methodol. 82, 1273–1300 (2020).

37. Nott, A. et al. Brain cell type–specific enhancer–promoter interactome maps and disease-risk association. Science 366, 1134–1139 (2019).

38. Cuomo, A. S. E. et al. Single-cell RNA-sequencing of differentiating iPS cells reveals dynamic genetic effects on gene expression. Nat. Commun. 11, 810 (2020).

39. Chen, S. et al. A genomic mutational constraint map using variation in 76,156 human genomes. Nature 625, 92–100 (2024).

40. Lakhani, C. M. et al. Integration of Deep Learning Annotations with Functional Genomics Improves Identification of Causal Alzheimer’s Disease Variants. Genetic and Genomic Medicine (2025).

41. Lopes, K. de P., et al. Genetic analysis of the human microglial transcriptome across brain regions, aging and disease pathologies. Nat. Genet. 54, 4–17 (2022).

42. Finucane, H. K. et al. Partitioning heritability by functional annotation using genome-wide association summary statistics. Nat. Genet. 47, 1228–1235 (2015).

43. Gazal, S. et al. Linkage disequilibrium-dependent architecture of human complex traits shows action of negative selection. Nat. Genet. 49, 1421–1427 (2017).

44. Romero-Molina, C., Garretti, F., Andrews, S. J., Marcora, E. & Goate, A. M. Microglial efferocytosis: Diving into the Alzheimer’s disease gene pool. Neuron 110, 3513–3533 (2022).

45. Kozlova, A. et al. PICALM Alzheimer’s risk allele causes aberrant lipid droplets in microglia. Nature (2025) doi:10.1038/s41586-025-09486-x.

46. Duchateau, L., Wawrzyniak, N. & Sleegers, K. The ABC’s of Alzheimer risk gene ABCA7. Alzheimers. Dement. 20, 3629–3648 (2024).

47. Belbasis, L., Morris, S., van Duijn, C., Bennett, D. & Walters, R. Mendelian randomization identifies proteins involved in neurodegenerative diseases. Brain 148, 2412–2428 (2025).

48. Zhan, H., Cammann, D., Cummings, J. L., Dong, X. & Chen, J. Biomarker identification for Alzheimer’s disease through integration of comprehensive Mendelian randomization and proteomics data. J. Transl. Med. 23, 278 (2025).

49. Vardarajan, B. N. et al. Identification of Alzheimer disease-associated variants in genes that regulate retromer function. Neurobiol. Aging 33, 2231.e15–2231.e30 (2012).

50. Schreiber, J. tangermeme: A toolkit for understanding cis-regulatory logic using deep learning models. Bioinformatics (2025).

51. Shrikumar, A., Greenside, P. & Kundaje, A. Learning important features through propagating activation differences. in *Proceedings of the 34th International Conference on Machine Learning - Volume 70* 3145–3153 (JMLR.org, Sydney, NSW, Australia, 2017).

52. Novikova, G. et al. Integration of Alzheimer’s disease genetics and myeloid genomics identifies disease risk regulatory elements and genes. Nat. Commun. 12, 1610 (2021).

53. Sakaue, S. et al. Tissue-specific enhancer-gene maps from multimodal single-cell data identify causal disease alleles. Nat. Genet. 56, 615–626 (2024).

54. Ota, M. et al. Causal modeling of gene effects from regulators to programs to traits: integration of genetic associations and Perturb-seq. bioRxivorg (2025) doi:10.1101/2025.01.22.634424.

55. Kunes, R. Z., Walle, T., Land, M., Nawy, T. & Pe’er, D. Supervised discovery of interpretable gene programs from single-cell data. Nat. Biotechnol. 42, 1084–1095 (2024).

56. Spence, J. P. et al. Specificity, length, and luck: How genes are prioritized by rare and common variant association studies. Genomics (2024).

57. Weissbrod, O. et al. Functionally informed fine-mapping and polygenic localization of complex trait heritability. Nat. Genet. 52, 1355–1363 (2020).

