## Supplementary Methods for "Machine Learning-Based Prediction of Cell-type Resolved Brain eQTLs Enhances Discovery of Variants Explaining Alzheimer’s Disease Heritability"

### Supplementary Note

#### 1 Gradient Boosted Decision Trees

The sc-EEMS model predicts the probability of a variant being a cell-specific single cell eQTL using a gradient boosted classification model called CatBoost (1). Given training data $\mathcal{D} = \{(\mathbf{x}_i, y_i)\}_{i=1}^N$ where $y_i \in \{0, 1\}$ indicates whether variant $i$ is an eQTL and $\mathbf{x}_i \in \mathbb{R}^d$ is the feature vector, we learn an additive ensemble of regression trees. We follow the exposition of gradient boosted decision trees found in the XGBoost paper (2).

##### 1.1 Model Formulation

The model uses an ensemble of $T$ regression trees to make predictions:

$$F^{(T)}(\mathbf{x}) = \sum_{t=1}^{T} f_t(\mathbf{x}) \tag{1}$$

where each $f_t(\mathbf{x})$ is a regression tree that outputs a real-valued score. The model output $F^{(T)}(\mathbf{x})$ represents the log-odds (logit) of the positive class. We denote the logit for example $i$ as:

$$\gamma_i = F^{(T)}(\mathbf{x}_i) \tag{2}$$

We also define the term $\gamma_i^{(t)}$ as an approximation of $\gamma_i$ using the first $t$ regression trees.

$$\gamma_i^{(t)} = \sum_{k=1}^{t} f_k(\mathbf{x}_i) \tag{3}$$

**Logit to Probability:** The sigmoid function converts the logit to a probability:

$$p_i = P(y_i = 1|\mathbf{x}_i) = \sigma(\gamma_i) = \frac{1}{1 + \exp(-\gamma_i)} \tag{4}$$

##### 1.2 Loss Function for Binary Classification

For binary classification, we use the logistic loss (negative log-likelihood). For a single training example $(\mathbf{x}_i, y_i)$ with logit $\gamma_i$:

$$\ell(y_i, \gamma_i) = -\left[y_i \log p_i + (1 - y_i) \log(1 - p_i)\right] \tag{5}$$

where $p_i = \sigma(\gamma_i)$.

Substituting the sigmoid function and simplifying, this can be written directly in terms of the logit:

$$\ell(y_i, \gamma_i) = \log(1 + \exp(\gamma_i)) - y_i\gamma_i \tag{6}$$

The total loss over all training examples is:

$$L = \sum_{i=1}^{N} \ell(y_i, \gamma_i) = \sum_{i=1}^{N} \ell(y_i, F^{(T)}(\mathbf{x}_i)) \tag{7}$$

#### 1.3 Gradient and Hessian for Binary Classification

To perform gradient boosting, we need the first and second derivatives of the loss with respect to the logit $\gamma_i$.

**First Derivative (Gradient):**

$$g_i = \frac{\partial \ell(y_i, \gamma_i)}{\partial \gamma_i} = \frac{\exp(\gamma_i)}{1 + \exp(\gamma_i)} - y_i = p_i - y_i \tag{8}$$

**Second Derivative (Hessian):**

$$h_i = \frac{\partial^2 \ell(y_i, \gamma_i)}{\partial \gamma_i^2} = \frac{\exp(\gamma_i)}{(1 + \exp(\gamma_i))^2} = p_i(1 - p_i) \tag{9}$$

The gradient $g_i = p_i - y_i$ represents the residual between the predicted probability and the true label. The Hessian $h_i = p_i(1 - p_i)$ represents the curvature of the loss function and is always positive, making it suitable for second-order optimization.

#### 1.4 Second-Order Taylor Approximation

Since the loss function is non-linear, we approximate it using a second-order Taylor expansion around the current logit $\gamma_i^{(t-1)}$:

$$\ell(y_i, \gamma_i^{(t-1)} + f_t(\mathbf{x}_i)) \approx \ell(y_i, \gamma_i^{(t-1)}) + g_i^{(t)} f_t(\mathbf{x}_i) + \frac{1}{2} h_i^{(t)} f_t(\mathbf{x}_i)^2 \tag{10}$$

where $g_i^{(t)}$ and $h_i^{(t)}$ are the gradient and Hessian evaluated at iteration $t$ using the current model prediction $\gamma_i^{(t-1)}$.

Removing constant terms (which don't affect optimization), the approximate objective becomes:

$$\tilde{L}^{(t)} = \sum_{i=1}^{N} \left[ g_i^{(t)} f_t(\mathbf{x}_i) + \frac{1}{2} h_i^{(t)} f_t(\mathbf{x}_i)^2 \right] \tag{11}$$

This quadratic approximation motivates the use of both gradients and Hessians in tree construction, leading to the formulas for optimal leaf weights and split gains described below.

#### 1.5 Additive Training via Gradient Boosting

Gradient boosting builds the ensemble iteratively by adding one tree at a time. At each iteration $t$, we improve the current model by adding a new tree that corrects the errors of the existing ensemble.

##### 1.5.1 Step 1: Initialization

Initialize with a constant prediction representing the overall log-odds:

$$F^{(0)}(\mathbf{x}) = \log\left(\frac{\bar{p}}{1-\bar{p}}\right) \tag{12}$$

where $\bar{p} = \frac{1}{N}\sum_{i=1}^{N} y_i$ is the proportion of positive examples in the training set.

##### 1.5.2 Step 2: Compute Residuals

For iteration $t$, compute the current predictions and residuals for each training example:

1. Convert current log-odds to probabilities:

$$p_i^{(t-1)} = \frac{1}{1+\exp(-F^{(t-1)}(\mathbf{x}_i))} \tag{13}$$

2. Calculate the gradient (residual) and Hessian (weight) at iteration $t$ for each example:

$$g_i^{(t)} = p_i^{(t-1)} - y_i, \quad h_i^{(t)} = p_i^{(t-1)}(1-p_i^{(t-1)}) \tag{14}$$

##### 1.5.3 Step 3: Tree Construction Process

Build a regression tree $f_t(\mathbf{x})$ that minimizes the approximate loss.

*Initialization.* Begin with the root node containing all training examples $\{(\mathbf{x}_i, y_i)\}_{i=1}^{N}$ with their associated gradients $\{g_i^{(t)}\}$ and Hessians $\{h_i^{(t)}\}$.

*Greedy Splitting.* For each internal node containing samples $I$, evaluate all possible splits:

- For each feature $j$ and threshold $s$, partition samples into:
  - Left child: $I_L = \{i \in I | x_{ij} \leq s\}$
  - Right child: $I_R = \{i \in I | x_{ij} > s\}$
- Compute the gain from this split:

$$\text{Gain} = \frac{1}{2}\left[\frac{(\sum_{i\in I_L} g_i^{(t)})^2}{\sum_{i\in I_L} h_i^{(t)}} + \frac{(\sum_{i\in I_R} g_i^{(t)})^2}{\sum_{i\in I_R} h_i^{(t)}} - \frac{(\sum_{i\in I} g_i^{(t)})^2}{\sum_{i\in I} h_i^{(t)}}\right] \tag{15}$$

- Select the split with maximum gain

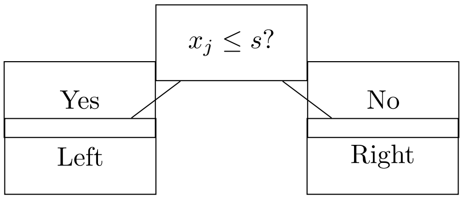


*Leaf Value Assignment.* For each terminal node (leaf) $j$ containing samples $I_j^{(t)}$, assign the optimal prediction value:

$$w_j^{(t)} = -\frac{\sum_{i \in I_j^{(t)}} g_i^{(t)}}{\sum_{i \in I_j^{(t)}} h_i^{(t)}} \tag{16}$$

This represents a Newton-Raphson step for minimizing the loss.

*Stopping Criteria.* Tree construction halts when:

1. Maximum depth is reached (depth = 6)
2. Minimum samples per leaf constraint is violated (samples $\geq 10$)
3. No split achieves sufficient gain improvement

###### 1.5.4 Step 4: Update Model

Add the new tree to the ensemble with a learning rate $\nu$:

$$F^{(t)}(\mathbf{x}) = F^{(t-1)}(\mathbf{x}) + \nu \cdot f_t(\mathbf{x}) \tag{17}$$

###### 1.5.5 Step 5: Repeat

Return to Step 2 and repeat for $T$ iterations.

###### 1.5.6 Step 6: Final Predictions

After training, the final probability prediction for a new example $\mathbf{x}$ is:

$$p(\mathbf{x}) = \frac{1}{1 + \exp(-F^{(T)}(\mathbf{x}))} \tag{18}$$

##### 1.6 Sample Weighting and Feature Weighting

###### 1.6.1 Sample Weighting

The sc-EEMS model applies sample weighting where each negative class variant has weight $s_i = 1$ and positive class variant has weight $s_i$ proportional to its PIP value. Sample weights are incorporated by scaling the contribution of each example to the loss function during each iteration of tree building $t$:

$$\tilde{L}^{(t)} = \sum_{i=1}^{N} s_i \cdot \left[ g_i^{(t)} f_t(\mathbf{x}_i) + \frac{1}{2} h_i^{(t)} f_t(\mathbf{x}_i)^2 \right] \tag{19}$$

This modification propagates through the gradient boosting framework:
**Weighted Gradients and Hessians:**

$$g_i^{(t)\text{weighted}} = s_i \cdot g_i^{(t)} = s_i(p_i^{(t-1)} - y_i) \tag{20}$$

$$h_i^{(t),\text{weighted}} = s_i \cdot h_i^{(t)} = s_i p_i^{(t-1)}(1 - p_i^{(t-1)}) \tag{21}$$

**Weighted Leaf Values:** The optimal leaf weight becomes:

$$w_j^{(t)} = -\frac{\sum_{i \in I_j^{(t)}} s_i g_i^{(t)}}{\sum_{i \in I_j^{(t)}} s_i h_i^{(t)}} \tag{22}$$

**Weighted Split Gain:** The gain formula for evaluating splits becomes:

$$\text{Gain} = \frac{1}{2} \left[ \frac{(\sum_{i \in I_L} s_i g_i^{(t)})^2}{\sum_{i \in I_L} s_i h_i^{(t)}} + \frac{(\sum_{i \in I_R} s_i g_i^{(t)})^2}{\sum_{i \in I_R} s_i h_i^{(t)}} - \frac{(\sum_{i \in I} s_i g_i^{(t)})^2}{\sum_{i \in I} s_i h_i^{(t)}} \right] \tag{23}$$

###### 1.6.2 Feature Weighting

The sc-EEMS model applies differential feature weighting to the feature vector $\mathbf{x_i} = (x_i^1, x_i^2, \ldots x_i^{d-1}, x_i^d)$ based on the feature category. The feature $x_i^k$ receives a feature weight $\lambda_k = 10$ if it is a deep learning feature and $\lambda_k = 1$ otherwise.

When evaluating a split on feature $k$, the gain is multiplied by the feature weight:

$$\text{Weighted Gain}_k = \lambda_k \cdot \text{Gain}_k \tag{24}$$

where $\text{Gain}_k$ is computed using the standard formula:

$$\text{Gain}_k = \frac{1}{2} \left[ \frac{(\sum_{i \in I_L} g_i^{(t)})^2}{\sum_{i \in I_L} h_i^{(t)}} + \frac{(\sum_{i \in I_R} g_i^{(t)})^2}{\sum_{i \in I_R} h_i^{(t)}} - \frac{(\sum_{i \in I} g_i^{(t)})^2}{\sum_{i \in I} h_i^{(t)}} \right] \tag{25}$$

At each split point, the algorithm compares the weighted gains across all features and selects the feature-threshold pair with the maximum weighted gain.

##### 1.7 CatBoost-Specific Implementation Details

Our implementation uses CatBoost with the following configuration:

##### 1.7.1 Model Parameters

```
model_params = {
    'depth': 6,
    'iterations': 1000,
    'learning_rate': 0.03,
    'l2_leaf_reg': 3.0,
    'min_data_in_leaf': 10
}
```

##### 1.7.2 Oblivious Decision Trees

CatBoost uses *oblivious* (also called symmetric) decision trees. In an oblivious tree, all nodes at the same depth level must use the same feature and threshold for splitting. This differs from standard decision trees where each node independently chooses its own splitting rule.

**Standard Decision Tree:** Each node at a level can split on different features.

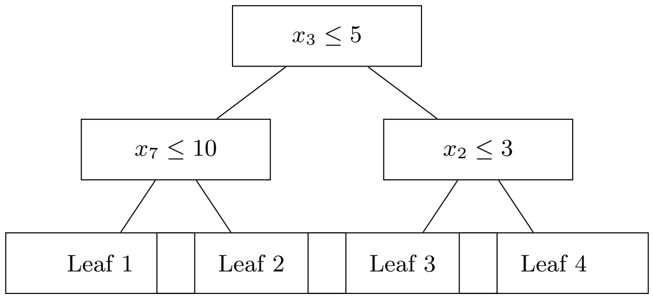


**Oblivious Decision Tree:** All nodes at the same level use the same split.

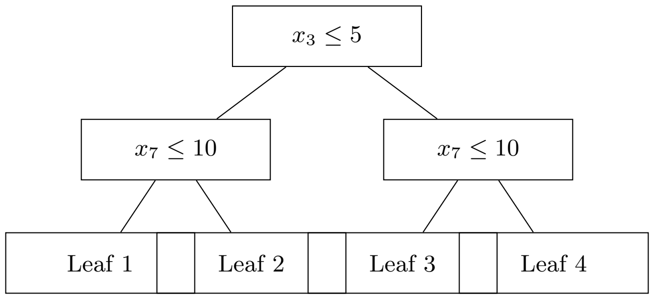


Notice that in the oblivious tree, both nodes at depth 1 use the identical split $x_7 \leq 10$, while in the standard tree they can differ.

For a tree of depth $d$, the oblivious structure:

- Uses only $d$ unique splitting rules (one per level)
- Creates exactly $2^d$ leaves

- Provides faster prediction time (simplified tree structure)
- Reduces overfitting (fewer parameters to fit)

###### 1.7.3 Ordered Boosting and Prediction Shift

Standard gradient boosting has a subtle overfitting problem. At each iteration $t$, gradients are computed using predictions from trees that were trained on all the data, including the current example $i$. This means when computing the gradient for $(\mathbf{x}_i, y_i)$, the model has already "seen" the true label $y_i$, which introduces a bias known as prediction shift (3).

CatBoost prevents this through *ordered boosting*:

1. Randomly order all training examples
2. For each example $i$, train a model using only examples that come *before* $i$ in this ordering
3. Use this model to compute the gradient $g_i^{(t)}$ for example $i$

This ensures gradients are computed from a model that has never seen the true label $y_i$, eliminating the bias.

###### 1.7.4 L2 Regularization

The `l2_leaf_reg` parameter adds L2 regularization to leaf values, modifying the optimal leaf weight formula:

$$w_j^{(t)} = -\frac{\sum_{i \in I_j^{(t)}} g_i^{(t)}}{\sum_{i \in I_j^{(t)}} h_i^{(t)} + \lambda} \tag{26}$$

where $\lambda = 3.0$ is the regularization coefficient. This shrinks leaf values toward zero, reducing overfitting by penalizing extreme predictions. Larger values of $\lambda$ result in more conservative leaf weights and smoother decision boundaries.

### 2 Construction of Predicted Prior Probabilities for Fine-Mapping

#### 2.1 Motivation for Recalibration

Due to the sample weighting scheme employed during model training, our classifier assumes an implicit prior probability of 0.5 for both the positive class (causal eQTL) and negative class (non-causal variant). However, the true prior probability of a variant being a causal eQTL is substantially lower in reality. To obtain realistic posterior probabilities that reflect the actual prevalence of causal eQTLs, we recalibrated the model's predicted probabilities using Bayes' rule.

#### 2.2 Derivation of Recalibration Formula

Let $\mathbf{x}$ represent the feature vector for a given variant-gene pair. The posterior probability of being a causal eQTL given features $\mathbf{x}$ under the training distribution is:

$$P_{\text{train}}(\text{eQTL} \mid \mathbf{x}) = \frac{P(\mathbf{x} \mid \text{eQTL}) \cdot P_{\text{train}}(\text{eQTL})}{P_{\text{train}}(\mathbf{x})} \tag{27}$$

where $P_{\text{train}}(\text{eQTL}) = 0.5$ due to our balanced training scheme.

Under the real-world distribution with prior $P_{\text{real}}(\text{eQTL}) = \pi$, the posterior probability is:

$$P_{\text{real}}(\text{eQTL} \mid \mathbf{x}) = \frac{P(\mathbf{x} \mid \text{eQTL}) \cdot P_{\text{real}}(\text{eQTL})}{P_{\text{real}}(\mathbf{x})} \tag{28}$$

We assume the likelihood $P(\mathbf{x} \mid \text{eQTL})$ is invariant to the prior distribution. Therefore, using Bayes' rule, we express the relationship between the training and real-world posterior probabilities:

$$P_{\text{real}}(\text{eQTL} \mid \mathbf{x}) = \frac{P_{\text{train}}(\text{eQTL} \mid \mathbf{x}) \cdot \frac{P_{\text{real}}(\text{eQTL})}{P_{\text{train}}(\text{eQTL})}}{P_{\text{train}}(\text{eQTL} \mid \mathbf{x}) \cdot \frac{P_{\text{real}}(\text{eQTL})}{P_{\text{train}}(\text{eQTL})} + P_{\text{train}}(\text{non-eQTL} \mid \mathbf{x}) \cdot \frac{P_{\text{real}}(\text{non-eQTL})}{P_{\text{train}}(\text{non-eQTL})}} \tag{29}$$

Substituting $P_{\text{train}}(\text{eQTL}) = 0.5$, $P_{\text{train}}(\text{non-eQTL}) = 0.5$, $P_{\text{real}}(\text{eQTL}) = \pi$, and $P_{\text{real}}(\text{non-eQTL}) = 1 - \pi$:

$$P_{\text{real}}(\text{eQTL} \mid \mathbf{x}) = \frac{P_{\text{train}}(\text{eQTL} \mid \mathbf{x}) \cdot \frac{\pi}{0.5}}{P_{\text{train}}(\text{eQTL} \mid \mathbf{x}) \cdot \frac{\pi}{0.5} + (1 - P_{\text{train}}(\text{eQTL} \mid \mathbf{x})) \cdot \frac{1-\pi}{0.5}} \tag{30}$$

#### 2.3 Logit Space Transformation

The recalibration can be expressed more simply in logit space. The logit transformation is defined as:

$$\text{logit}(p) = \log\left(\frac{p}{1-p}\right) \tag{31}$$

The logit of the training posterior is:

$$\text{logit}_{\text{train}}(\mathbf{x}) = \log\left(\frac{P_{\text{train}}(\text{eQTL} \mid \mathbf{x})}{P_{\text{train}}(\text{non-eQTL} \mid \mathbf{x})}\right) \tag{32}$$

Through algebraic manipulation of equation (32), we can show that the recalibrated logit is:

$$\text{logit}_{\text{real}}(\mathbf{x}) = \text{logit}_{\text{train}}(\mathbf{x}) + \log\left(\frac{P_{\text{real}}(\text{eQTL})/P_{\text{real}}(\text{non-eQTL})}{P_{\text{train}}(\text{eQTL})/P_{\text{train}}(\text{non-eQTL})}\right) \tag{33}$$

Substituting our priors:

$$\text{logit}_{\text{real}}(\mathbf{x}) = \text{logit}_{\text{train}}(\mathbf{x}) + \log\left(\frac{\pi/(1-\pi)}{0.5/0.5}\right) \tag{34}$$

This simplifies to:

$$\text{logit}_{\text{real}}(\mathbf{x}) = \text{logit}_{\text{train}}(\mathbf{x}) + \log\left(\frac{\pi}{1-\pi}\right) \tag{35}$$

#### 2.4 Application to sc-EEMS

For our analysis, we determined the realistic prior probability $\pi$ based on empirical observations from well-powered eQTL studies. The GTEx consortium found an average of 1.8 cis-eQTLs per gene in their best-powered tissues(4). Given that our analysis tested approximately 14,716 cis-variants per gene (variants within ±1MB of the transcription start site), this yields a prior probability of:

$$\pi = \frac{1.8}{14,716} \approx 0.00012 \tag{36}$$

The logit adjustment factor for this prior is:

$$\log\left(\frac{0.00012}{1-0.00012}\right) = \log\left(\frac{0.00012}{0.99988}\right) \approx -8.93 \tag{37}$$

Therefore, for each variant, we recalibrated the predicted probability by:

1. Computing the logit of the training probability: $\text{logit}_{\text{train}} = \log\left(\frac{p_{\text{train}}}{1-p_{\text{train}}}\right)$
2. Adjusting by the prior correction: $\text{logit}_{\text{real}} = \text{logit}_{\text{train}} - 8.93$
3. Converting back to probability scale: $p_{\text{real}} = \frac{1}{1+e^{-\text{logit}_{\text{real}}}}$

These recalibrated probabilities were then used as priors in the SuSiE fine-mapping framework to improve identification of causal eQTLs.

### References


[1] Dorogush, A. V., Ershov, V. & Gulin, A. Catboost: gradient boosting with categorical features support (2018). URL `https://arxiv.org/abs/1810.11363`. 1810.11363.

[2] Chen, T. & Guestrin, C. Xgboost: A scalable tree boosting system. In *Proceedings of the 22nd ACM SIGKDD International Conference on Knowledge Discovery and Data Mining*, KDD '16, 785–794 (Association for Computing Machinery, New York, NY, USA, 2016). URL `https://doi.org/10.1145/2939672.2939785`.

[3] Dorogush, A. V. *et al.* Fighting biases with dynamic boosting. *arXiv preprint arXiv:1706.09516* (2017).

[4] GTEx Consortium *et al.* The GTEx consortium atlas of genetic regulatory effects across human tissues. *Science* **369**, 1318–1330 (2020). URL `https://www.science.org/doi/abs/10.1126/science.aaz1776`.
